# Inferring Sexual Network Bridging Using Genomics: A Simulation Study

**DOI:** 10.64898/2026.05.24.26353967

**Authors:** Madeleine C. Kline, David Helekal, Kirstin Oliveira Roster, Yonatan Grad

## Abstract

The dynamics of sexually transmitted infections involve interconnected transmission networks, including men who have sex with men and heterosexual populations. Understanding the extent of bridging between these networks can inform surveillance, guide interventions, and aid in the interpretation of their impact, but methods for quantifying bridging have been lacking. Here, we addressed whether pathogen genomics tools, successfully used to reconstruct transmission in other contexts, could accurately infer sexual network bridging. Based on simulations of gonorrhea spread, we evaluated phylodynamic bridging metrics inferred by ancestral state reconstruction under a range of sampling schemes, from comprehensive to sparse. These metrics differentiated sexual network structures even with biased sampling schemes, but accuracy depended on the sampling scheme and density: phylodynamic bridging estimates using sequences from all detected infections for one network configuration were on average 6.9% above the true value, whereas estimates from 5% of infections in symptomatic men with many partners were on average >1000% above the true value. These results suggest routine overestimation of bridging from unadjusted inferences from genomics data and provide context for interpreting existing genomic surveillance data and targeted studies.

## Introduction

Modeling pathogen spread between populations is critical for understanding infectious disease epidemiology and for guiding and interpreting interventions. For diseases transmitted through respiratory routes, age groups play an important role in epidemic spread^1^ and can be modeled using age-structured contact matrices based on social interactions.^2^ For sexually transmitted infections (STIs), in contrast, spread is more complex to model because sexual partnerships have varying durations, rates of exposure, and can overlap, and links between heterosexual and single-sex networks, e.g., men who have sex with men (MSM), are unclear.

Current methods for estimating the dynamics that bridge heterosexual populations and populations of MSM include detailed behavioral surveys^3,4^ and pathogen phylogenetic analyses. However, these techniques do not provide the quantitative measures of bridging needed for targeted deployment of sexually transmitted infection (STI) interventions or the evaluation of their impact.

Understanding spread between networks is especially important in the study of gonorrhea, caused by the pathogen *Neisseria gonorrhoeae*, which is an urgent public health problem due to a high burden of disease and increasing antibiotic resistance.^5–14^ Critical public health questions center on anticipating in which populations highly drug resistant isolates are likely to appear and how long, if no more cases are detected, before one can be confident that there is no ongoing transmission.^15^ Further, as doxycycline post-exposure prophylaxis (doxy-PEP) to prevent bacterial sexually transmitted infections is recommended only for men who have sex with men (MSM) and transgender women (TGW),^16^ defining its indirect impact on heterosexual populations requires an estimate of the extent of bridging between these MSM and heterosexual networks.^17^

Pathogen genomic information is increasingly used to monitor disease epidemiology and respond to emerging threats.^18,19^ For *N. gonorrhoeae*, a common method to evaluate sexual network patterns is to generate genomic clusters based on sequence similarity and then assess cluster features, such as sexual behavior group representation.^20–22^ Some studies incorporate epidemiologic transmission pair data to connect clusters or comment on the duration of persistence of a cluster.^23,24^ While many gonorrhea surveillance efforts exist,^10,11,25–27^ the limited sampling in these programs makes it unclear whether common phylodynamic analyses could accurately estimate bridging of heterosexual and MSM populations, and, if the sampling is insufficient, what level of sampling would be necessary.

Here, we addressed these questions by implementing an individual-based gonorrhea transmission model with dynamic partnership formation, simulating genome sequences, and reconstructing bridging parameters using ancestral state reconstruction. We 1) defined a quantitative bridging metric that differentiated sexual network structures, 2) assessed whether phylodynamic estimates of the bridging metric obtained using ancestral state reconstruction differentiated simulations with varying bridging levels, and 3) determined whether these phylodynamic bridging estimates covaried with the “ground truth” bridging metrics using sequences from i) every infection and ii) infections meeting various sampling criteria and across a range of sampling densities.

## RESULTS

### Metrics to distinguish between frequent and infrequent bridging scenarios

We first established an agent-based network framework for simulating and analyzing spread within and between heterosexual and MSM sexual networks. Men who have sex with men and women (MSMW) formed the bridges between the two networks, and the modeled population consisted of MSMW, MSM, women who have sex with men (WSM), and men who have sex with women (MSW). As MSMW were the bridges between the networks, they could be classified either as part of the MSM network or the heterosexual network. We thus analyzed bridging both ways, using two bridging proportion terms which captured the proportion of all transmission events that counted as bridging. Bridging proportion 1 treated MSMW as part of the MSM network, such that a transmission event between MSMW and the heterosexual network would be considering bridging, and bridging proportion 2 treated MSMW as part of the heterosexual network, such that a transmission event between MSMW and the MSM network would be considered bridging (**Table 1**, **Figure 1, Methods**).

**Figure 1:**
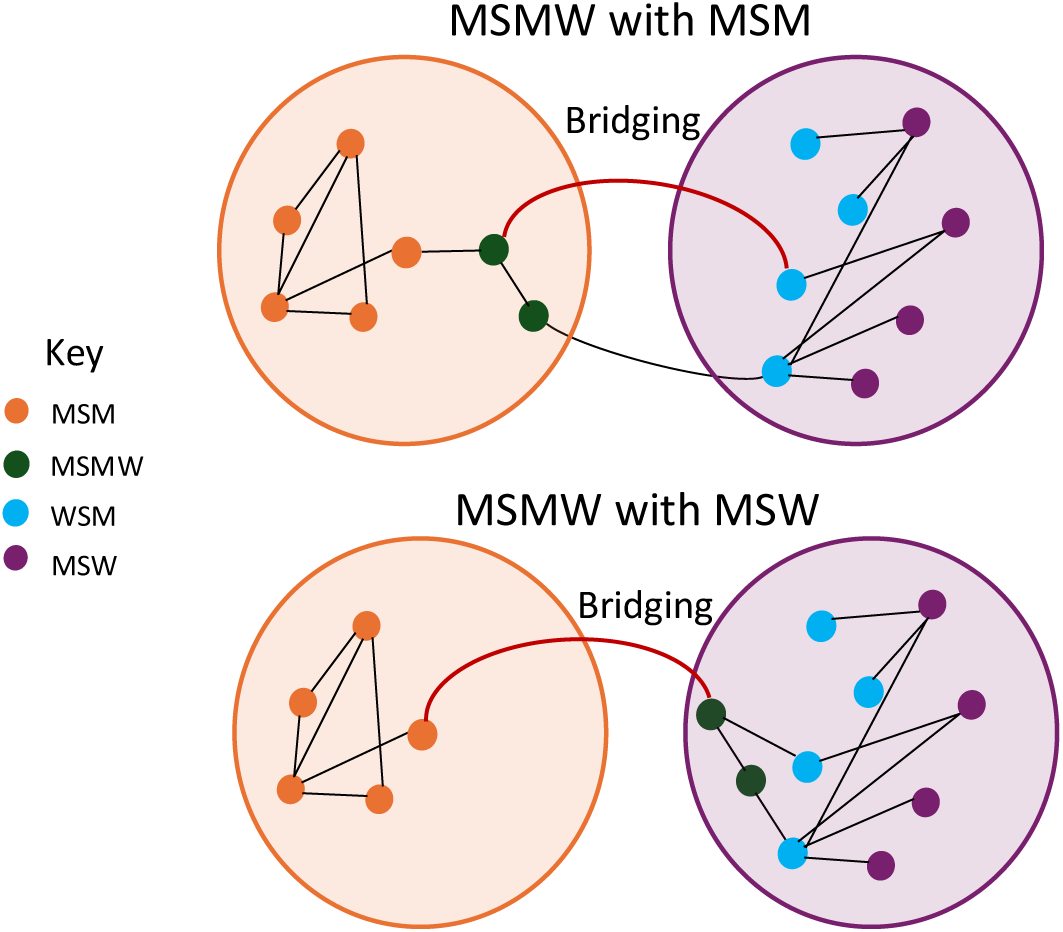
Bridging Definitions. The bridging proportion measures the proportion of all transmission events that bridge between network A and network B and depends on how network A and B are defined. When men who have sex with men and women (MSMW) are grouped with men who have sex with men (MSM), any transmissions that are between MSMW and women who have sex with men (WSM) count as between-network transmissions. When MSMW are grouped with MSW, any transmissions between MSM and MSMW count as between-network transmissions. Small circles represent agents in the agent-based transmission model. Larger circles represent groupings of networks A and B. Lines connecting agents are sexual partnerships comprising a sexual network configuration at a given point in time. The red lines highlight a transmission that would be designated a bridging transmission if it occurred between the two agents in that partnership. Colors represent sexual behavior group.

**Table 1:** Terminology.

| Short name | Definition |
| --- | --- |
| $\rho$ | Probability that, when an MSMW agent seeks a new partner, they will seek a female partner |
| Bridging proportion 1 | Proportion of transmissions that occur between networks, where MSMW are grouped with MSM, so the two networks are MSM + MSMW and MSW + WSM |
| Bridging proportion 2 | Proportion of transmissions that occur between networks, where MSMW are grouped with MSW, so the two networks are MSM and MSMW + MSW + WSM |
| Bridging metric | Bridging proportion 1 x bridging proportion 2<br>The product of bridging proportion 1, which is the proportion transmissions between network when MSMW are grouped with MSM, and bridging proportion 2, which is the proportion of transmission between network when MSMW are grouped with MSW. |
| Bridging values | Bridging proportion 1, bridging proportion 2, and the bridging metric |
| Ground truth | The bridging proportions or bridging metric calculated from the full transmission log documenting all simulation period transmission in the gonorrhea simulation model |
| Phylogenetic estimate | The bridging proportions or bridging metric estimated using our ancestral state reconstruction procedure |
| Sampling scheme | All infections from one of the following sub-populations: detected only, symptomatic only, symptomatic men, high activity symptomatic men, or a random subsample of high activity (**Supplementary Table 4**). The sampling scheme datasets themselves are comprehensive, meaning they include all infections that meet the definition. |
| Coverage level | The proportion of yearly infections in the sampling scheme that are included. A coverage level of 25% means that 25% of the annual complete sampling scheme infections are represented. |
| Dataset | A set of sequences that represents all sequences, a comprehensive sampling scheme, or a sampling scheme with incomplete coverage, on which the phylogenetic pipeline was run. Each one could represent a dataset that would be collected in the real world. |

We defined the parameter ρ as the probability that, when an agent classified as MSMW sought a new partner, the agent would seek a female partner. This parameter thus served as the primary input driving sexual network connectivity: when ρ was low, MSMW individuals were functionally MSM, and when high, they were functionally MSW.

To confirm that we could modulate bridging with simulation input parameters, we ran simulations for each of five values of ρ (0.05, 0.25, 0.5, 0.75 and 0.95; see **Methods** and **Supplementary Table 1**). Bridging proportion 1 peaked with ρ of 0.75 (median: 6.72 × 10^−2^, IQR: 5.92 × 10^−2^ – 7.50 × 10^−2^; **Figure 2, Supplementary Table 1**), and bridging proportion 2 peaked with ρ of 0.25 (median: 6.58 × 10^−2^, IQR: 5.86 × 10^−2^ – 7.37 × 10^−2^). Thus, these two measures together differentiated all five network configurations. Multiplying the two bridging proportion terms yielded the bridging metric, which distinguished frequent and infrequent bridging (0.05 median: 2.77 × 10^−4^, IQR: 2.04 × 10^−4^– 3.75 × 10^−4^; 0.5 median: 2.46 × 10^−3^, IQR: 1.99 × 10^−3^ – 3.10 × 10^−3^). These bridging proportion terms and metric defined the “ground truth”, which we then used as the standard for comparison in our genomics-based inferences.

**Figure 2:**
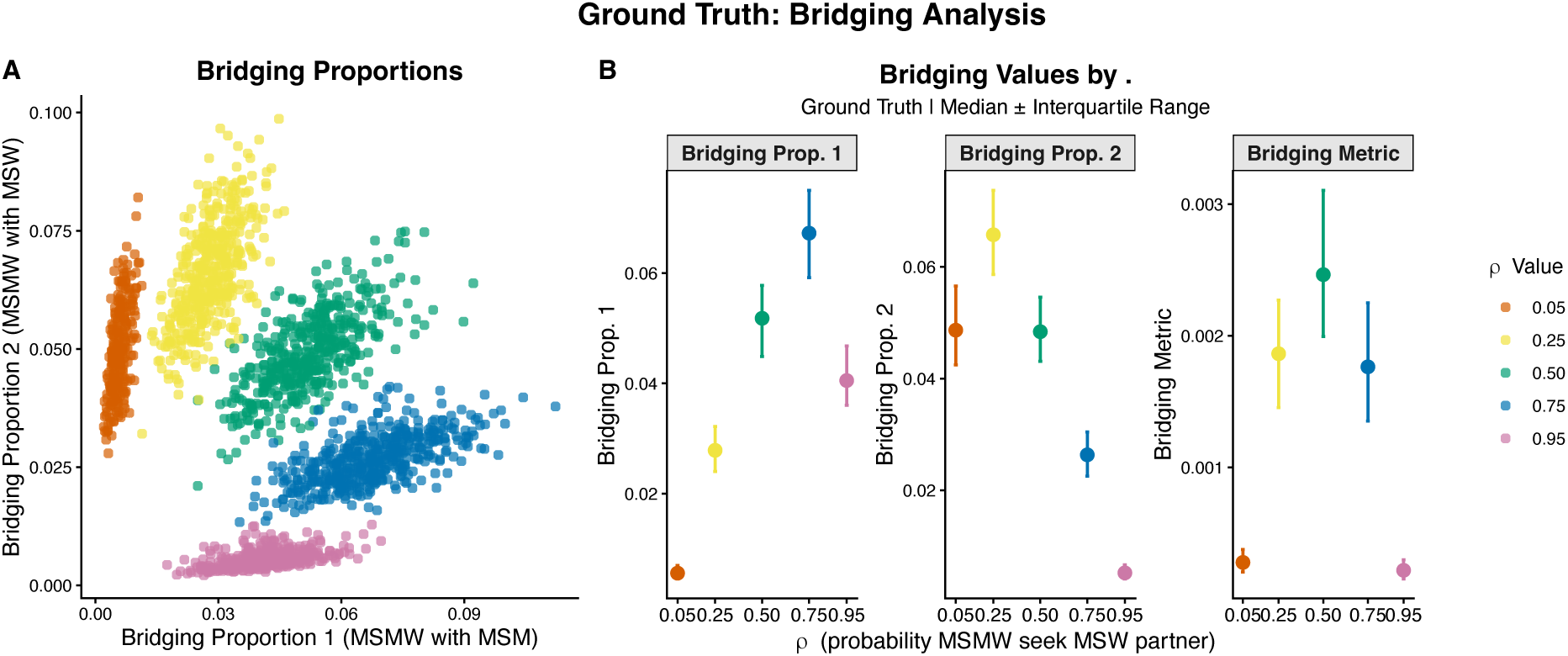
Ground Truth Bridging. The bridging proportions and bridging metric in simulations with different inputs of ρ, the probability that an MSMW agent will seek a female partner when seeking a new partner. Panel A: the bridging proportions for each of the two groupings of MSMW, either grouped with MSM (bridging proportion 1) or with MSW (bridging proportion 2). Panel B: the median bridging proportions for each grouping separately as a function of ρ, (left and middle panels) and their product, which is the bridging metric (right panel). Colors represent different ρ values, ranging from 0.05 where MSMW behave more like MSM to 0.95 where MSMW behave more like MSW; error bars represent interquartile ranges. Results are from 1000 simulations attempts for each value of ρ.

### Phylodynamic Bridging Estimates from Comprehensive Sampling of All Infections Differentiated Underlying Sexual Network Structures and Covaried with the Ground Truth

#### Differentiating Sexual Network Structures with Phylodynamic Bridging Value Estimates

We next sought to ask how well phylodynamic analysis of *N. gonorrhoeae* genome sequences with ancestral state reconstruction could recover ground truth bridging in a total information scenario where sequences were available for all infections (**Figure 3**, **Supplementary Figure 1**). Our phylodynamic bridging proportions were obtained by building phylogenetic trees, inferring ancestral states, and counting the proportion of parent-daughter relationships where a state change occurred (**Methods**). These phylodynamic estimates differentiated among simulations by input values of ρ. The phylodynamic estimate of bridging proportion 1 peaked at ρ of 0.5 (median: 4.66 × 10^−2^, IQR: 4.09 × 10^−2^ – 5.52 × 10^−2^; **Supplementary Table 2**). The phylodynamic estimate of bridging proportion 2 peaked at ρ of 0.25 (median: 5.05 × 10^−2^, IQR: 4.87 × 10^−2^ – 5.49 × 10^−2^). The bridging metric estimates differentiated frequent and infrequent bridging networks (0.05 median: 1.55 × 10^−4^, IQR: 1.53 × 10^−4^– 1.84 × 10^−4^; 0.5 median: 2.08 × 10^−3^, IQR: 1.62 × 10^−3^– 2.91 × 10^−3^).

**Figure 3:**
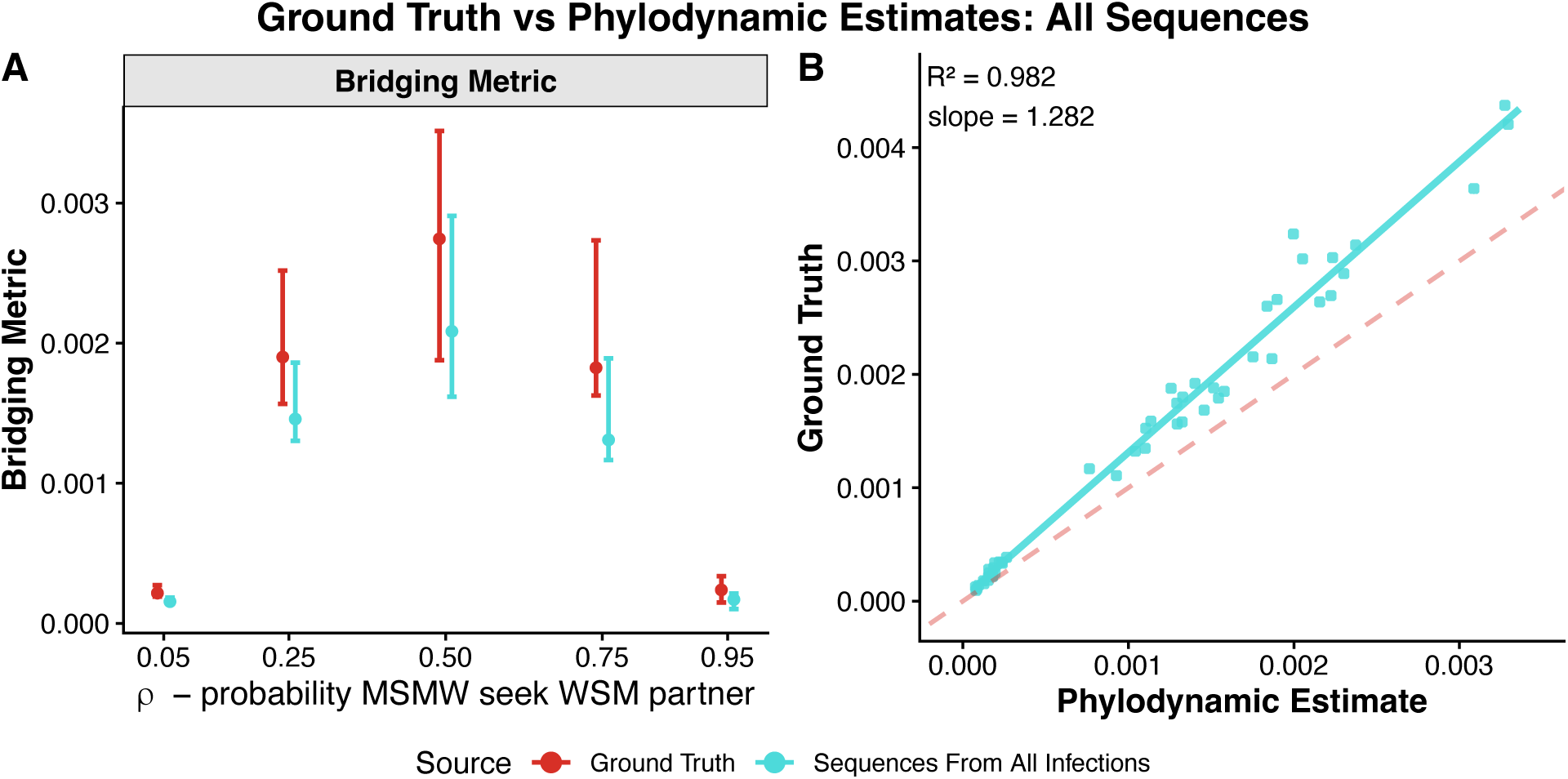
Phylodynamic Bridging Metric Estimates from All Sequences. Comparing phylodynamic estimates of the bridging metric to ground truth bridging metric by ρ value. ρ is the probability that, when seeking a new partner, an MSMW agent seeks a female partner. Panel A: The bridging metric (product of bridging proportion 1 and bridging proportion 2) by ρ value is on the X-axis. Colors and left to right orientation differentiate the ground truth (red) from the phylodynamic estimate using sequences from all infections (blue). Points are the median across 10 replicates and error bars are the interquartile ranges. Panel B: The bridging metric from the phylodynamic estimate is on the X-axis and the ground truth is on the Y axis, pooled across values of ρ. The dashed line indicates perfect agreement between the two measures. Results are from 10 simulations for each ρ value run with 10000 agents with a partnership burn-in of 2000 days, transmission burn-in of 10000 days, and simulation period of 3650 days.

#### Covariation of Phylodynamic Estimates of the Bridging Metric and Ground Truth

The phylodynamic bridging metric estimates using sequences for all infections covaried with but consistently underestimated the ground truth (R^2^ = 0.982, slope = 1.282, **Figure 3B**, **Supplementary Table 3**). For bridging proportion 1, the phylodynamic estimates more closely aligned with the ground truth in sexual networks where MSMW largely acted as MSM (low ρ) but diverged from the ground truth in sexual networks where MSMW largely acted as MSW (high ρ). Conversely, for bridging proportion 2, phylodynamic estimates more closely aligned with the ground truth in sexual networks where MSMW largely acted as MSW (high ρ) but diverged from the ground truth in sexual networks where MSMW largely acted as MSM (low ρ). The median bridging metric estimate was at most about 30% away from the ground truth across ρ values (**Supplementary Table 2**).

### Phylodynamic Bridging Metric Estimates Differentiated Sexual Network Structures Even in Biased Sampling Schemes

#### Differentiating Sexual Network Structures with Phylodynamic Estimates Across Sampling Schemes

We then evaluated how well phylodynamic inference recovered the ground truth bridging metric assuming only subsets of all infections were available through surveillance. To do so, we compared the ground truth bridging metric from 10 simulations for each ρ value to the estimated bridging metric calculated from our phylodynamic analysis for each sampling scheme. The sampling schemes were designed to represent more realistic surveillance systems (**Supplementary Table 4**, **Methods**). As with the phylodynamic bridging metric estimates using sequence data from all infections, each sampling scheme differentiated between underlying sexual network structures, as indicated by ρ (**Figure 4**). For example, the median bridging metric estimate in the scenario in which we sampled all infections from a random subset of high activity agents for ρ of 0.5 was 8.28 × 10^−3^ (IQR: 7.21 × 10^−3^ – 8.71 × 10^−3^) and when ρ was 0.95, it was 1.78 × 10^−3^ (IQR: 1.05 × 10^−3^ – 2.22 × 10^−3^) (**Supplementary Table 2**).

**Figure 4:**
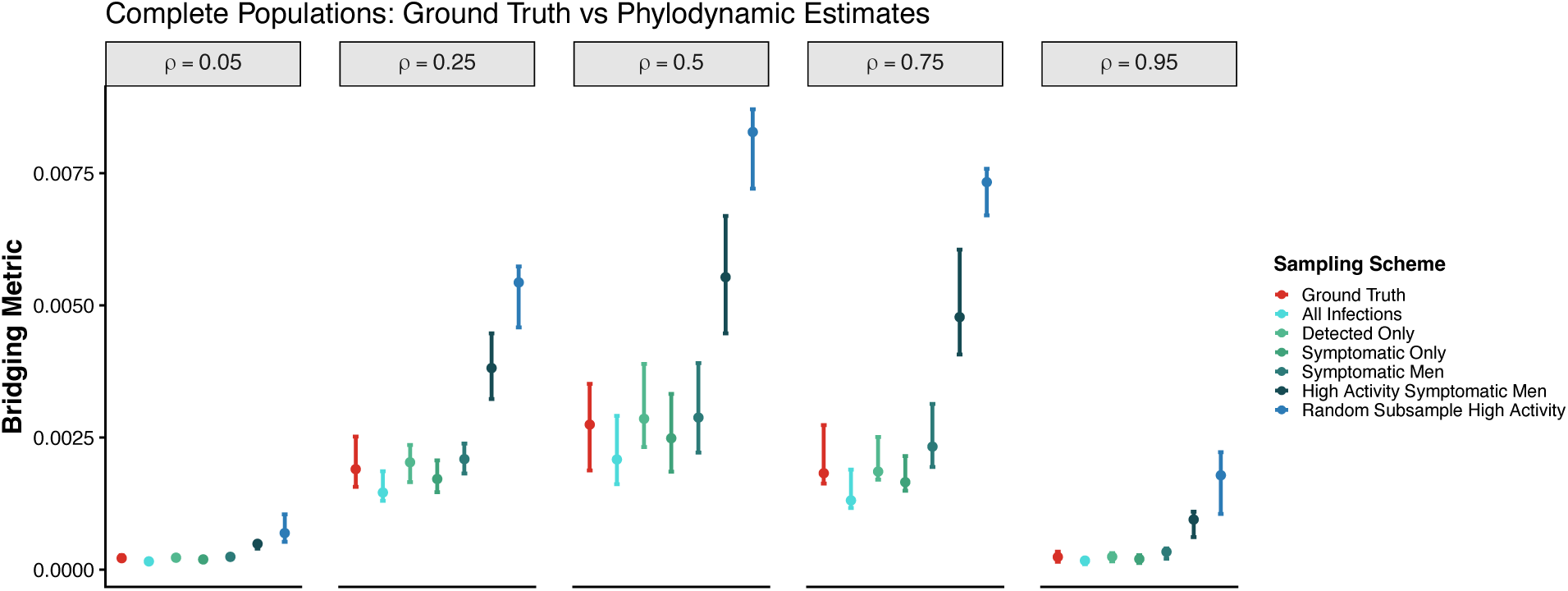
Phylodynamic Bridging Metric Estimates from Sampling Schemes. Comparing phylodynamic estimates of bridging metrics from different sampling schemes to ground truth bridging metrics by ρ value. The Y-axis shows the bridging metric (point = median, error bars = IQR). Colors and left to right orientation show the ground truth compared to each of the sampling schemes. The facet labels show the ρ value for that set of simulations, which represents the probability that MSMW seek female partners when they are seeking a new partner. Results are from 10 simulations for each ρ value run with 10000 agents with a partnership burn-in of 2000 days, transmission burn-in of 10000 days, and simulation period of 3650 days.

#### Covariation of Ground Truth and Phylodynamic Estimates Across Sampling Schemes

Phylodynamic bridging metric estimates from each sampling scheme covaried with the ground truth but did so less as the sampling became more biased (**Figure 5, Supplementary Table 3**). Estimating the bridging metric from all infections, only detected infections, only symptomatic infections, or infections in symptomatic men resulted in similarly high coefficients of determination (R^2^ = 0.982, 0.976, 0.975, and 0.960 respectively). However, the covariation decreased for the most biased sampling schemes, which captured only high activity individuals (‘high activity symptomatic men’ R^2^ = 0.895, ‘random subsample high activity’ R^2^ = 0.733). The difference from the ground truth also increased for more biased sampling schemes. For example, for ρ = 0.5, the bridging metric estimate from the ‘detected only’ population was 6.9% (IQR: 0.4% - 15%) above the ground truth (absolute difference 1.61 × 10^−4^, IQR: 1.01 × 10^−5^ – 2.96 × 10^−4^). For ρ = 0.5, the bridging metric estimate from the ‘high activity symptomatic men’ population was 137% (IQR: 89 – 142%) above the ground truth (**Figure 4, Supplementary Table 2**).

**Figure 5:**
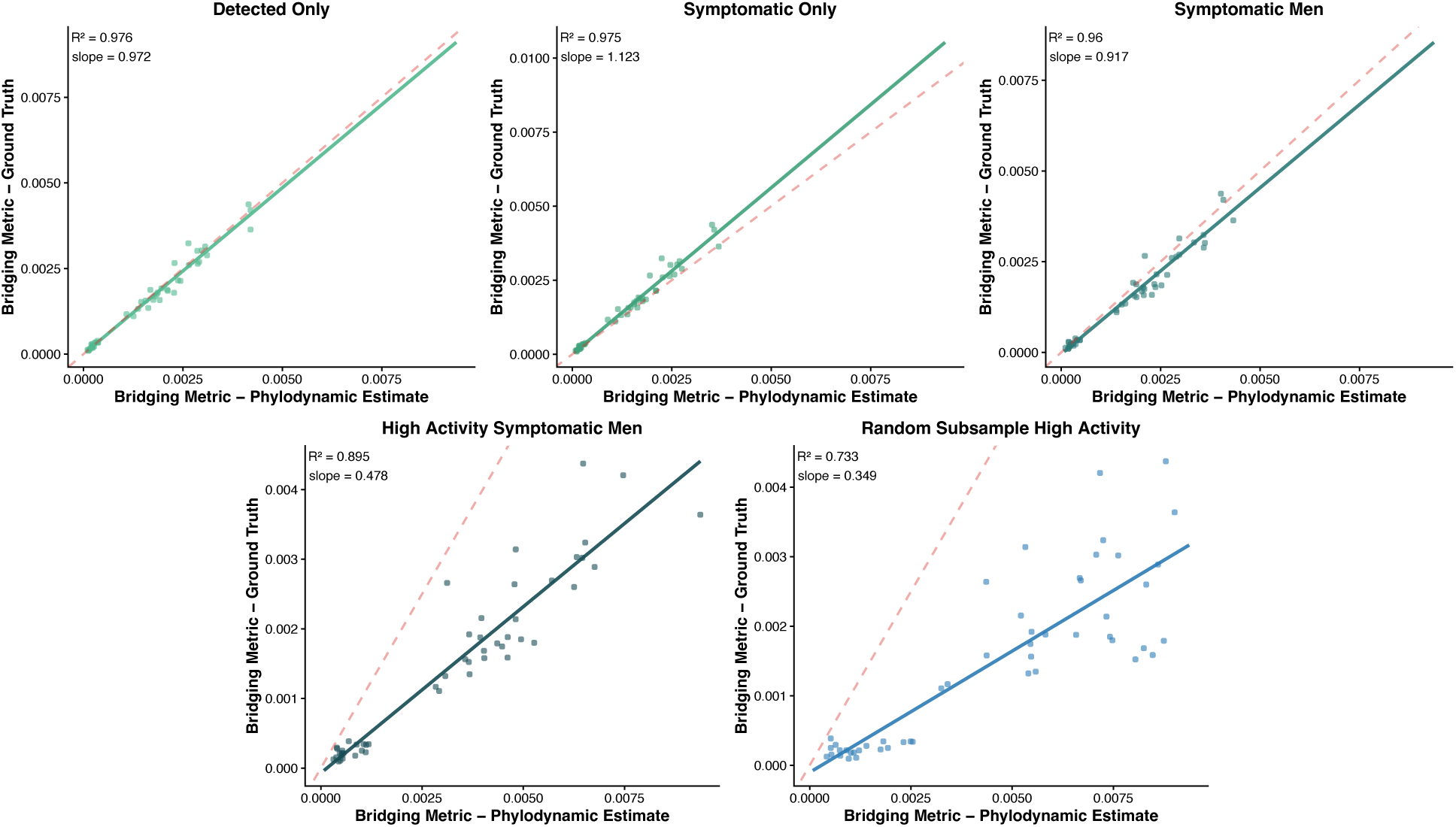
Comparing Sampling Scheme Bridging Metric Estimates to Ground Truth. Each panel shows the regression of the phylodynamic estimate of the bridging metric on the ground truth from that simulation for a specific sampling scheme, assuming all infections from that sampling scheme are captured. All ρ values, which represents the probability that MSMW seek female partners when they are seeking a new partner, are pooled. The top left shows the coefficient of determination (R^2^) and slope. The dashed lines show perfect one-to-one relationship with ground truth (slope of 1).

### Phylodynamic Bridging Metric Estimates Differentiated ρ Values in Sampling Schemes with Incomplete Coverage

#### Relationship Between Ground Truth and Phylodynamic Bridging Metric Estimates from Sampling Schemes with Incomplete Coverage

We next asked how sampling only a fraction of the infections impacted the accuracy of inferences about bridging. To do so, we evaluated the bridging metrics for each sampling scheme assuming we captured 50%, 25%, 10%, 5%, 4%, 3%, 2%, and 1% of annual infections. As coverage within each sampling scheme decreased, the phylodynamic bridging metric estimates increased (**Figure 6A**). For the more biased sampling schemes (‘high activity symptomatic men’ and ‘random subsample high activity’), and the more sparse coverage levels (e.g., 2% and 1% of yearly infections), the phylodynamic estimates were much greater than the ground truth. For example, the phylodynamic estimate of the bridging metric for the ρ value of 0.5, the sampling scheme ‘high activity symptomatic men,’ and coverage of 1%, was on average 1730% above the ground truth bridging metric (IQR: 1160% - 2190%, absolute difference: 4.37 × 10^−2^, IQR: 3.86 × 10^−2^ – 4.82 × 10^−2^).

**Figure 6:**
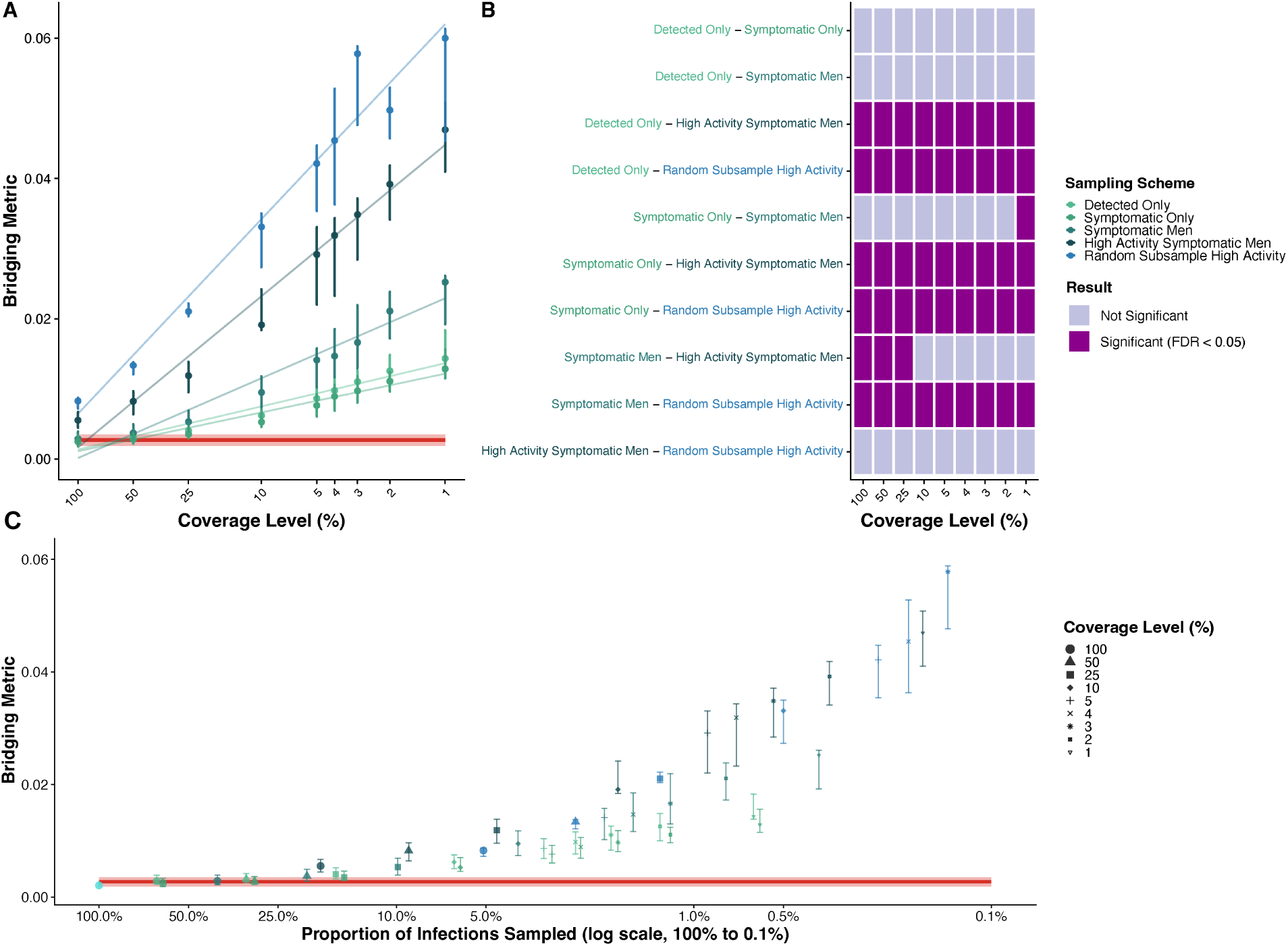
Phylodynamic Bridging Metric Estimates from Sampling Schemes with Incomplete Coverage, ρ = 0.5. Comparing phylodynamic estimates of bridging metrics from sampling schemes with incomplete coverage to ground truth bridging metrics when ρ = 0.5. All panels show results when ρ, which represents the probability that MSMW seek female partners when they are seeking a new partner, is 0.5. Panel A: The Y-axis represents the bridging metric (point = median of medians, error bars = IQR). The X-axis shows each coverage level for each of the datasets averaged across the simulations. Colors show the ground truth (red line) compared to each of the sampling scheme datasets representing sampling of specific subsets of the population. The horizontal line and shading show the median and IQR for the ground truth. Linear fits are shown to represent the rate of increase of overestimation with decreasing coverage for each sampling scheme. Panel B: Each box represents a pairwise Dunn test performed post-hoc following significant Kruskal-Wallis analysis showing that there were significant differences between sampling schemes at a given coverage level. Dunn test results show which of the sampling schemes within a coverage level are significantly different from one another. False discovery rate (FDR) was corrected for with the Benjamini-Hochberg correction. Panel C: The same datapoints from panel A are shown plotted on an X-axis that represents the proportion of total infections represented by downsampling from that sampling scheme. Point shape and size represent the coverage level. Results are from 10 simulations for each ρ value run with 10000 agents with a partnership burn-in of 2000 days, transmission burn-in of 10,000 days, and simulation period of 3650 days, with 10 replicates per simulation for each coverage level of each dataset.

To assess how decreasing coverage impacted accuracy across sampling schemes, we compared estimated bridging metrics between sampling schemes within a given coverage level and ρ value. We found significant differences between estimates among sampling schemes (all p values <1 × 10^−6^ by Kruskal-Wallis test, **Supplementary Table 5**), with the performance of each sampling scheme dependent on the coverage levels (**Figure 6B, Supplementary Figure 2, Supplementary Table 6**). Estimated bridging metrics from ‘detected only’ and ‘symptomatic only’ sampling schemes did not differ at any coverage level or ρ value. Estimated bridging metrics from ‘detected only’, ‘symptomatic only’, and ‘symptomatic men’ sampling schemes always differed from the ‘random subsample of high activity agents’, regardless of coverage level of ρ value. Estimated bridging metrics from ‘detected only’ or ‘symptomatic only’ and ‘high activity symptomatic men’ sampling schemes were also always significant, regardless of coverage level or ρ value. However, when coverage was sparse, estimated bridging metrics from the ‘symptomatic men’ sampling scheme did not differ significantly from those of ‘high activity symptomatic men’, (**Figure 6B, Supplementary Figure 2, Supplementary Table 6**).

To account for the fact that more selective sampling schemes captured fewer of the overall infections, we plotted each coverage level from all sampling schemes by the proportion of all infections they captured (**Figure 6C**). There were differences between sampling schemes even when they represented similar proportions of overall infections; for example, 4% coverage level of the ‘detected only’ sampling scheme represented about 2.5% of infections and had a bridging metric about 256% above ground truth for ρ = 0.5, whereas 50% coverage level of the ‘random subsample of high activity’ sampling scheme also represented about 2.5% of infections, but had a bridging metric 390% above ground truth for ρ = 0.5 (**Supplementary Tables 2 & 7**).

#### Differentiating Sexual Network Structures in Sampling Schemes with Incomplete Coverage

For all coverage levels, the phylodynamic estimate differentiated between underlying sexual network structures (**Supplementary Figure 3**). For example, the median bridging metric estimate in the dataset that captured 1% of infections from a random subsample of high activity agents for ρ = 0.5 was 0.060 (IQR: 0.045 – 0.061) and when ρ = 0.95, it was 0.024 (IQR: 0.012–0.036, **Supplementary Table 2**).

## DISCUSSION

Despite the importance of bridging across sexual networks for understanding the spread of sexually transmitted diseases, quantitative methods to measure bridging have been lacking. Comprehensive behavioral surveys to measure sexual network connectivity are possible, but they are often expensive, challenging to administer, and may not always be accurate.^28^ Here, we introduced a quantitative bridging metric and explored the genomic sampling necessary to recover it via phylodynamic reconstruction. Our bridging metric was obtained by calculating the proportion of transmissions that were classified as between-network, which we could modulate with the underlying model parameter ρ. The between-network proportion can be calculated by grouping those who bridge across networks, MSMW, with each of the two populations, MSM and MSW. Combining the two groupings into the product of the between-network proportions, the bridging metric, differentiated the five sexual network structures evaluated here. With an understanding of how the bridging metric relates to ρ and other model parameters, it could be mapped back to these parameters and used to test interventions in a transmission model.

The accuracy of phylodynamic estimation depended on the amount of bias in the sampling scheme and the degree of sampling. When we analyzed sequences from every infection – the “best case” scenario – estimates correlated with true bridging but consistently underestimated it. This could be due to multiple factors, including: i) many sequences are identical, given the rate of mutation and sampling density; and ii) some of the transmission events defined as between network may be mostly within network transmission. For example, if agent 1 is MSMW and has 1 MSM partner and 1 WSM partner in a year and agent 1 transmits back and forth twice to the WSM partner and only once to the MSM partner, that would count as 4 between-network transmissions when MSMW are grouped with MSM, and 1 between-network transmission when MSMW are grouped with MSW. However, the phylodynamic analysis will likely only detect one instance where a sequence similar to what is transmitting among MSW/WSM reaches the MSM population.

Given that recovering the *N. gonorrhoeae* genome sequence from every infection is unrealistic, we explored alternative sampling schemes to determine the impact on accuracy. The goal of this analysis was to assess whether an accurate, quantitative estimate of bridging could be recovered from surveillance datasets and, if not, how samples could be collected if the explicit goal is to recover bridging. The ‘detected only’, ‘symptomatic only’, and ‘symptomatic men only’ sampling schemes yielded phylodynamic estimates of the bridging metric that were similar to one another and to the ground truth. This indicates that the bias introduced by selectively filtering towards symptomatic men did not substantially impair accuracy, likely because the probability of a new infection being symptomatic was not substantially different between men and women. However, the high activity symptomatic men and the random subsample of high activity individuals both overestimated population mixing. These two sampling schemes are most like typical surveillance samples, which draw from sexual health clinics,^25^ suggesting that broader sampling in routine surveillance would help improve estimates of sexual network bridging. Biasing towards symptom status was less problematic than biasing towards high activity. This is likely because, in the model, MSM and MSMW had greater high activity proportions than MSW and WSM, and having higher activity increases the chances of having a bridging transmission event. Alternative model parameterizations could be explored in future work to determine under what scenarios this continues to hold true.

While lower levels of coverage yielded phylodynamic bridging metric estimates with similar rates of divergence from the ground truth in the ‘detected only’ and ‘symptomatic only’ datasets, the ‘symptomatic men’ sampling scheme estimates diverged from ground truth as the coverage dropped. Comparing sampling schemes and coverage levels that captured a similar proportion of all infections showed that more biased sampling schemes were still less accurate; the effect was not due solely to capturing fewer infections. Across all sampling schemes, sampling smaller proportions of infections led to increasing phylodynamic estimates of population mixing. This indicates that, if researchers seek to collect samples to measure bridging, the measurements will be more accurate with less coverage if activity status is more reflective of the total population in the sampled population.

Our simulations allowed us to quantify the phylodynamic bridging metric estimate relative to the ground truth. In the most extreme cases, for the ‘random subsample high activity’ sampling scheme, ρ of 0.5, and coverage level of 1%, the estimated bridging metric was on average 1850% above the ground truth. This indicates that estimates of bridging calculated via phylodynamic reconstruction are likely to overestimate bridging in the population. Even in the most biased scenarios (sampling a small proportion of infections in only a subset of the population), our phylodynamic procedure differentiated among sexual network structures. This indicates that comparing estimated bridging metrics from populations with similar surveillance systems that capture similar population groups at similar sampling intensities could yield a valid relative comparison. However, if two populations were sampled differently or have different transmission dynamics (e.g., more high activity WSM), comparing bridging estimates could be misleading.

Our study has several limitations. Our individual-based model made assumptions, including that each agent can only form one new partnership per day, all MSMW have the same probability of seeking a male vs. female partner, and there was no latent period between infection and becoming infectious, no partner treatment, no behavior changes with symptom status, and no test of cure. We also simplified our population to four behavior groups: MSW, WSM, MSMW, and MSM. Because most interventions (e.g., doxy-PEP) have been targeted at MSM in the U.S., we chose to focus this study on bridging involving MSMW.^29–31^ Additionally, while interventions like doxy-PEP are targeted towards MSM and TGW, we did not separately model TGW.^16^ While transgender individuals are an important and diverse group, it is difficult to obtain group-specific parameters and our transmission model does not explicitly model body site of infection. In our phylodynamic analysis, we simulated genome sequences with only one random seed and did not incorporate uncertainty or alternative parameterizations into our phylogenetic tree building, tree dating, polytomy resolution, or ancestral state reconstruction. In addition, unlike ancestral state reconstruction, the structured coalescent model is robust to sampling bias, but it is more difficult to implement and computationally expensive.^32^ Our work serves as a basis for future exploration and optimization, where each modular component (e.g., transmission model parameters, sequence simulation, sampling strategies, ancestral state reconstruction) could be implemented with alternative parameters, software, and tools.

In summary, we proposed a new measure that distinguished between sexual network structures with different levels of bridging. We described a phylodynamic analysis procedure which produced bridging estimates that covaried with ground truth bridging, even when only analyzing subsets of sequences, for example, only from symptomatic men. Across all sampling schemes, sampling smaller proportions of infections led to increasing phylodynamic estimates of population mixing. When the sampling schemes were more representative, they could withstand more downsampling before deviating from the ground truth. Thus, phylodynamic bridging analyses provided a quantitative estimate of bridging, when compared across populations with similar sampling schemes and densities, and otherwise similar sexual transmission dynamics. Applying this method to routine surveillance will likely overestimate population bridging and could therefore serve as an upper bound. To obtain more accurate bridging estimates, genome sequences would need to be more representative of overall sexual activity level distributions.

Using pathogen genomic data provides a complementary bridging estimation approach that will be increasingly useful as sequencing continues to expand and become more affordable. Our analyses provide guidance on how best to design and interpret surveillance data to infer rates of bridging. In addition to operationalizing a quantitative bridging measure and identifying key elements of STI genomic surveillance that influence bridging estimation, our modular framework serves as a basis for evaluating future questions that rely on different transmission dynamics, sampling strategies, and phylodynamic reconstruction methods.

## Methods

The four steps required to estimate bridging phylodynamically were: 1) establish a dynamic partner formation model, which creates the sexual network; 2) generate a susceptible-infected-susceptible transmission model upon the sexual network; 3) simulate sequence evolution along the transmission chains from the model; 4) infer bridging via phylogenetics and ancestral state reconstruction (**Figure 7**).

**Figure 7:**
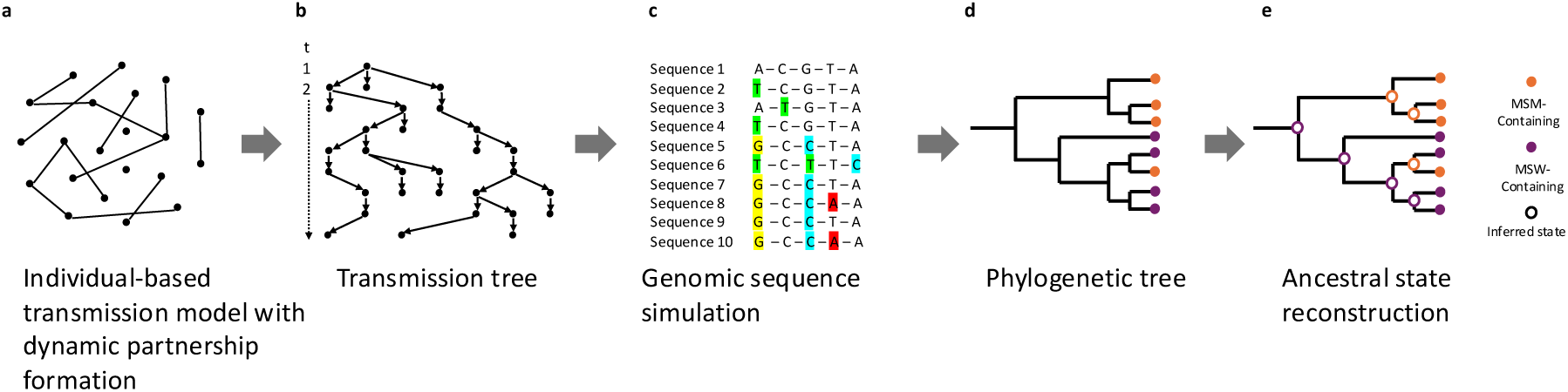
Summary of Core Methodological Procedure. The core analysis steps included running an individual-based gonorrhea transmission model, constructing a transmission tree from the transmission log, simulating genomic sequences from the transmission tree, building a phylogenetic tree from the sequence alignment, and inferring ancestral states from the phylogenetic tree. Panel A: A snapshot representing a sexual network at a single time point during the simulation. Dots represent agents in the agent-based model, lines represent network edges which are sexual partnerships between two agents. Transmission occurred along connected edges. Panel B: An example transmission tree generated with daily chains from simulated transmission log data. Each dot is an infected day of a simulation period infection. Panel C: A representation of sequences simulated from the transmission tree. Panel D: A representation of a phylogenetic tree generated from the simulated sequence data. Tips are annotated with the sexual behavior group of the agents infected with the infections the tip samples represent. Panel E: A representation of ancestral state reconstruction to infer the sexual behavior group states of internal nodes.

### Individual-Based Transmission and Pair Formation Model

Pair formation and transmission were simulated as a stochastic individual-based model. The simulated population consisted of four behavior groups, MSM, MSMW, WSM, and MSW, each of which was stratified into high and low activity subgroups based on the average annual number of partners (**Supplementary Table 8**).^4,33–35^ Steady and casual partnerships formed and dissolved daily to create a dynamic sexual network adapted from prior work.^36^ The primary parameter connecting sexual networks was ρ, the probability that a new partner for an MSMW agent was female, which was fixed for all agents in the population (**Table 1**). Transmission could occur within an active partnership with asymmetric infection status (**Supplementary Table 9**). The model accounted for symptomatic and asymptomatic infections, natural clearance, and treatment (**Supplementary Methods**, **Supplementary Table 9**).

### “Ground Truth” Bridging

Our primary bridging measure was the proportion of between-network transmissions, referred to here as the bridging proportion (**Table 1**). Because a bridging proportion requires two networks, we divided our simulated population in two ways and calculated the transmission proportion for each grouping: 1) grouping MSMW with MSM, so that network A was MSM+MSMW and network B was MSW+WSM (bridging proportion 1), 2) grouping MSMW with MSW so that network A was MSM only and network B was MSW+WSM+MSMW (bridging proportion 2) (**Figure 1**). This resulted in two bridging proportions for each transmission log: bridging proportion 1 and bridging proportion 2. Before grouping agents into network A or network B for these two sets of groupings, we extracted the dynamic behavior classification based on whether the agent’s sexual partners in the year prior to sampling were male, female, or both. This led to the reclassification of some MSMW as either MSM or MSW for specific infections. We then calculated the proportion of all transmission events in the simulation period that were between network A and network B for both definitions of network A and network B, thereby determining the “ground truth” bridging for each simulation. To summarize the extent of bridging using information from bridging proportion 1 and bridging proportion 2, we defined the “bridging metric” as the product of bridging proportion 1 and bridging proportion 2.

### Phylodynamic Bridging Estimation Procedure

#### Bioinformatics

We built hybrid trees from the transmission simulation with a tip for each infection and simulated genomic sequences using Seq-Gen version 1.3.5^37^ with parameters in **Supplementary Table 10** (**Supplementary Methods**). We generated a maximum-likelihood phylogenetic tree from the Seq-Gen sequences using IQ-TREE version 3.0.1^38^ with the GTR+G evolutionary model and fast option. We extracted sampling dates for each infection and dated and rooted the tree using LSD2.^39^ We then extracted tip states (sexual behavior groups) and used TreeTime version 0.11.4 with the “mugration” command to infer ancestral states.^40,41^

#### Sampling Schemes and Coverage Levels

To emulate more realistic specimen sampling, we filtered and downsampled the alignment of all sequences from the simulation to create a total of five sampling schemes: ‘detected only’, ‘symptomatic only’, ‘symptomatic men’, ‘high activity symptomatic men’, and all infections in a ‘random subsample of high activity’ agents (**Supplementary Table 4**). Each sampling scheme was then downsampled to include coverage of 50%, 25%, 10%, 5%, 4%, 3%, 2% and 1% of yearly infections. For each coverage level, 10 different samples were created. The full phylodynamic analysis then proceeded for each of the 401 (5 sampling schemes x 8 coverage levels × 10 replicates per coverage level = 400 + all sequences dataset = 401) datasets for each simulation.

#### Phylodynamic Bridging

To calculate phylodynamic bridging, we assigned tree tips a state based on their dynamic behavior classification and the two groupings of networks A and B, and ran TreeTime mugration twice, once for each grouping. We assumed that each tree tip had a known behavior designation. We then mapped tree tips and internal nodes to their inferred ancestral states from TreeTime and counted the bridging proportion by counting the proportion of parent nodes that were inferred to be in a different state compared to the daughter node out of all parent-daughter relationships. As with ground truth bridging, there were two measures of bridging proportions for each phylogenetic tree - bridging proportion 1 when MSMW were grouped with MSM and bridging proportion 2 when MSMW were grouped with MSW. The bridging metric is the product of bridging proportion 1 and bridging proportion 2 and was the main bridging value in this analysis.

### Simulated Data

#### Assessing The Bridging Values

Because our stochastic individual-based model produced slightly different results for each run, we conducted a parameter sweep to determine the average behavior of the system over repeated simulations. We ran 1,000 simulations with different seeds for each of five values of ρ, the primary bridging parameter in our simulation. Each simulation was run with 10,000 agents, a partnership burn-in of 2,000 days, a transmission burn-in of 10000 days, and a simulation period of 3,650 days (10 years, **Supplementary Table 11**). For each simulation, we determined whether all simulation period infections were of a single lineage descended from a single initial infector and, if so, extracted dynamic behavior classifications and calculated ground truth bridging for that simulation. We restricted to simulations where all simulation period infections descended from a single initial infector to mirror the process of estimating phylodynamic bridging (**Supplementary Methods**).

#### Comparing Ground Truth and Phylodynamic Bridging

We calculated both ground truth and phylodynamic bridging estimates for a total of 50 simulations: 10 simulations for each value of ρ, 0.05, 0.25, 0.5, 0.75, and 0.95, with a population size of 10,000, partnership burn-in of 2,000 days, transmission burn-in of 10,000 days, and simulation period of 3,650 days (10 years).

### Statistical Analysis

#### Assessing Covariation Between Ground Truth and Phylodynamic Estimates

As the phylodynamic-derived bridging estimates represent a different quantity than the ground truth bridging, we assessed covariation between the two rather than statistical difference. For each comprehensive sampling scheme, we pooled phylodynamic estimates across ρ values, regressed them on their paired ground truth bridging metrics, and calculated an R^2^ coefficient of determination. We also performed Kruskal-Wallis tests of all sampling schemes within each coverage level with post-hoc Dunn tests to assess the extent of error between sampling schemes as coverage decreased. We applied a global Benjamini-Hochberg FDR correction across Kruskal-Wallis tests and stratum-specific Benjamini-Hochberg FDR corrections within Dunn tests with a significance level of 0.05.

### Software

All pipelines were run with Snakemake version 9.13.4.^42^ Scripts were executed in python version 3.13.7 and R 4.5.1. Simulations were run on the FASRC Cannon high powered computing cluster supported by the FAS Division of Science Research Computing group at Harvard University. Harvard AI Sandbox GPT’s Claude 4 Sonnet and ChatGPT 5 were used for coding assistance. All code can be found at: https://github.com/gradlab/bridging_genomics

## Data Availability

All data produced in the present work are contained in the manuscript

https://github.com/gradlab/bridging_genomics

## Acknowledgements

This work was supported by award Number T32AG51108 from the National Institute of Aging (M.C.K), and by NIH R01 AI132606 (Y.H.G). The content is solely the responsibility of the authors and does not necessarily represent the official views of the National Institute of General Medical Sciences, the National Institute of Aging, or the National Institutes of Health. The authors acknowledge the contributions of the other members of the Grad Lab and the Center for Communicable Disease Dynamics.

## Competing Interests

Y.H.G is on the scientific advisory boards of Kanso Diagnostics and Claryx.

## Contributions

All authors meet authorship criteria. M.C.K. and Y.H.G conceptualized the project. M.C.K designed and implement the code for the transmission model, performed bioinformatics analyses, performed statistical analyses, created data visualizations and wrote the first draft of the manuscript. D.H. contributed to the code development, bioinformatic analyses, data interpretation, and statistical analyses. K.O.R contributed to the code development, data interpretation, data visualization, and supervised the project. Y.H.G contributed to the analysis plan, data interpretation, and supervised the project. All authors contributed to and revised the manuscript. Y.H.G and K.O.R supervised the project and contributed as co-senior authors.

## References

1. Loeb, M. et al. Effect of Influenza Vaccination of Children on Infection Rates in Hutterite Communities: A Randomized Trial. JAMA 303, 943–950 (2010).

2. Mossong, J. et al. Social Contacts and Mixing Patterns Relevant to the Spread of Infectious Diseases. PLOS Medicine 5, e74 (2008).

3. Erens, B., McManus, S., Prescott, A. & Field, J. National Survey of Sexual Attitudes and Lifestyles II.

4. Haderxhanaj, L. T., Leichliter, J. S., Aral, S. O. & Chesson, H. W. Sex in a Lifetime: Sexual Behaviors in the United States by Lifetime Number of Sex Partners, 2006–2010. Sex Transm Dis 41, 345–352 (2014).

5. WHO. Gonorrhoea (Neisseria gonorrhoeae infection). https://web.archive.org/web/20250107065052/ https://www.who.int/news-room/fact-sheets/detail/gonorrhoea-(neisseria-gonorrhoeae-infection) (2025).

6. CDC. Slides from STI Surveillance, 2023 | STI Statistics. https://web.archive.org/web/20250610071713/ https://www.cdc.gov/sti-statistics/annual/slides.html (2025).

7. Eyre, D. W. et al. Detection in the United Kingdom of the *Neisseria gonorrhoeae* FC428 clone, with ceftriaxone resistance and intermediate resistance to azithromycin, October to December 2018. Euro Surveill 24, 1900147 (2019).

8. Hal, S. J. van et al. Emergence of an extensively drug-resistant *Neisseria gonorrhoeae* clone. The Lancet Infectious Diseases 24, e547–e548 (2024).

9. Reimche, J. L. et al. Novel strain of multidrug non-susceptible *Neisseria gonorrhoeae* in the USA. The Lancet Infectious Diseases 24, e149–e151 (2024).

10. Unemo, M. et al. WHO global gonococcal antimicrobial surveillance programmes, 2019–22: a retrospective observational study. The Lancet Microbe 101181 (2025) doi:10.1016/j.lanmic.2025.101181.

11. Maatouk, I. et al. Antimicrobial resistance in *Neisseria gonorrhoeae* in nine sentinel countries within the World Health Organization Enhanced Gonococcal Antimicrobial Surveillance Programme (EGASP), 2023: a retrospective observational study. The Lancet Regional Health - Western Pacific 61, 101663 (2025).

12. Ye, M. et al. Emergence of Neisseria gonorrhoeae Clone with Reduced Susceptibility to Sitafloxacin in China: An In Vitro and Genomic Study. Antibiotics 13, 468 (2024).

13. CDC. Drug-Resistant Gonorrhea | Gonorrhea. https://web.archive.org/web/20250226154939/ https://www.cdc.gov/gonorrhea/hcp/drug-resistant/index.html (2025).

14. Cyr, S. S. Update to CDC’s Treatment Guidelines for Gonococcal Infection, 2020. MMWR Morb Mortal Wkly Rep 69, (2020).

15. Oliveira Roster, K. I., et al. Estimating the undetected burden and the likelihood of strain persistence of drug-resistant Neisseria gonorrhoeae. American Journal of Epidemiology kwae455 (2024) doi:10.1093/aje/kwae455.

16. Bachmann, L. H. CDC Clinical Guidelines on the Use of Doxycycline Postexposure Prophylaxis for Bacterial Sexually Transmitted Infection Prevention, United States, 2024. MMWR Recomm Rep 73, (2024).

17. Gopalkrishnan, K., Kyaw, T. & Klausner, J. D. Can doxycycline prophylaxis indirectly reduce congenital syphilis? Clin Infect Dis ciaf694 (2025) doi:10.1093/cid/ciaf694.

18. Ladner, J. T., Grubaugh, N. D., Pybus, O. G. & Andersen, K. G. Precision epidemiology for infectious disease control. Nat Med 25, 206–211 (2019).

19. Grad, Y. H. & Lipsitch, M. Epidemiologic data and pathogen genome sequences: a powerful synergy for public health. Genome Biology 15, 538 (2014).

20. Williamson, D. A. et al. Bridging of Neisseria gonorrhoeae lineages across sexual networks in the HIV pre-exposure prophylaxis era. Nat Commun 10, 3988 (2019).

21. Mortimer, T. D. et al. The Distribution and Spread of Susceptible and Resistant Neisseria gonorrhoeae Across Demographic Groups in a Major Metropolitan Center. Clinical Infectious Diseases 73, e3146–e3155 (2021).

22. Town, K. et al. Phylogenomic analysis of Neisseria gonorrhoeae transmission to assess sexual mixing and HIV transmission risk in England: a cross-sectional, observational, whole-genome sequencing study. Lancet Infect Dis 20, 478–486 (2020).

23. Town, K. et al. Exploring and Comparing the Structure of Sexual Networks Affected by Neisseria gonorrhoeae Using Sexual Partner Services Investigation and Genomic Data. Sexually Transmitted Diseases 48, S131 (2021).

24. Taouk, M. L. et al. Longitudinal genomic analysis of Neisseria gonorrhoeae transmission dynamics in Australia. Nat Commun 15, 8076 (2024).

25. Gonococcal Isolate Surveillance Project (GISP) Profiles | STI Statistics | CDC. https://web.archive.org/web/20250610071716/ https://www.cdc.gov/sti-statistics/gisp-profiles/index.html (2025).

26. Gonococcal Antimicrobial Surveillance Programme (GASP). https://web.archive.org/web/20250830051424/ https://www.who.int/initiatives/gonococcal-antimicrobial-surveillance-programme.

27. Lahra, M. M., van Hal, S. & Hogan, T. R. Australian Gonococcal Surveillance Programme Annual Report, 2023. Commun Dis Intell (2018) 49, (2025).

28. Graham, C. A., Catania, J. A., Brand, R., Duong, T. & Canchola, J. A. Recalling sexual behavior: A methodological analysis of memory recall bias via interview using the diary as the gold standard. The Journal of Sex Research 40, 325–332 (2003).

29. Rahman, N., Ghanem, K. G., Gilliams, E., Page, K. R. & Tuddenham, S. Factors associated with sexually transmitted infection diagnosis in women who have sex with women, women who have sex with men and women who have sex with both. Sex Transm Infect 97, 423–428 (2021).

30. Gorgos, L. M. & Marrazzo, J. M. Sexually Transmitted Infections Among Women Who Have Sex With Women. Clin Infect Dis 53, S84–S91 (2011).

31. Women Who Have Sex with Women (WSW) and Women Who Have Sex with Women and Men (WSWM). https://web.archive.org/web/20211031091024/ https://www.cdc.gov/std/treatment-guidelines/wsw.htm (2021).

32. De Maio, N., Wu, C.-H., O’Reilly, K. M. & Wilson, D. New Routes to Phylogeography: A Bayesian Structured Coalescent Approximation. PLoS Genet 11, e1005421 (2015).

33. Mercer, C. H. et al. Changes in sexual attitudes and lifestyles in Britain through the life course and over time: findings from the National Surveys of Sexual Attitudes and Lifestyles (Natsal). The Lancet 382, 1781–1794 (2013).

34. NSFG - Listing S - Key Statistics from the National Survey of Family Growth. https://web.archive.org/web/20250819093310/ https://www.cdc.gov/nchs/nsfg/key_statistics/s-keystat.htm#sexactbetwmen (2025).

35. Prah, P. et al. Men who have sex with men in Great Britain: comparing methods and estimates from probability and convenience sample surveys. Sex Transm Infect 92, 455–463 (2016).

36. Kretzschmar, M., Van Duynhoven, Y. T. H. P. & Severijnen, A. J. Modeling Prevention Strategies for Gonorrhea and Chlamydia Using Stochastic Network Simulations. American Journal of Epidemiology 144, 306–317 (1996).

37. Rambaut, A. & Grass, N. C. Seq-Gen: an application for the Monte Carlo simulation of DNA sequence evolution along phylogenetic trees. Bioinformatics 13, 235–238 (1997).

38. Wong, T. K. F. et al. IQ-TREE 3: Phylogenomic Inference Software using Complex Evolutionary Models. https://ecoevorxiv.org/repository/view/8916/ (2025).

39. To, T.-H., Jung, M., Lycett, S. & Gascuel, O. Fast Dating Using Least-Squares Criteria and Algorithms. Syst Biol 65, 82–97 (2016).

40. Sagulenko, P., Puller, V. & Neher, R. A. TreeTime: Maximum-likelihood phylodynamic analysis. Virus Evol 4, vex042 (2018).

41. Hadfield, J. et al. Nextstrain: real-time tracking of pathogen evolution. Bioinformatics 34, 4121–4123 (2018).

42. Mölder, F. et al. Sustainable data analysis with Snakemake. Preprint at 10.12688/f1000research.29032.1 (2021).

