## Supplementary Materials for "Inferring Sexual Network Bridging Using Genomics: A Simulation Study"

### **Supplementary Methods**

#### Partnership Formation Model

##### *Population initialization*

The simulated population was initialized by dividing the total population into behavior groups based on specified behavior proportions (**Supplementary Table 8**). Within behavior groups, each agent was assigned to be either high or low activity based on the high activity proportion of that behavior group. The men who have sex with men (MSM) and men who have sex with men and women (MSMW) behavior groups had a higher proportion high activity than women who have sex with men (WSM) and men who have sex with women (MSW)(**Supplementary Table 8**).^1–4^ Each agent was then assigned a new partner request frequency, which was drawn from a shifted gamma distribution corresponding to the agent’s behavior group and activity level. For example, if the value assigned to an agent was 0.5, they would seek a new partner every two years on average. Each agent was also assigned a steady partner capacity value, which determined how many steady partners that agent could have concurrently and could be 0, 1, or 2. The probability of having a given steady partner capacity depended on whether the agent was high or low activity.

##### *Partnership formation*

At each time step, the model drew a random number for each agent to determine whether that agent was seeking a new partner. For agents in the MSMW behavior group, an additional random draw determined if that agent was seeking a female partner (WSM) or a male partner (MSM or MSMW), based on the value of ρ, the probability that a new partner was female, which was fixed for all agents in the population (**Table 1**). The agents seeking new partners were matched with one another based on their allowed partner types. To form matches, the list of seekers was shuffled, and then each other seeker was iterated through until another seeker with a compatible behavior was identified. Once a pair was formed, both agents in the pair were removed from the seeker pool so that only one new partnership could be formed in each time step.

Once two agents were matched, the partnership was assigned to be steady or casual by comparing each agents’ current number of steady partners to their steady partner capacity. If either agent was already at their maximum number of steady partners, the new partnership would be casual. Otherwise, it would be steady. Once the steady vs. casual status of the partnership was determined, a duration was drawn from the appropriate steady or casual duration distribution sampler.^5^ In each time step, expired partnerships dissolved and then new partnerships formed. Whether an agent sought a new partnership was based only on their partner-seeking rate and not how many partners they had currently. There was a partnership formation burn-in during which the sexual network was established prior to transmission occurring in the system.

#### Gonorrhea Transmission Model

##### *Seeding*

Following partnership formation burn-in, initial infections were seeded randomly based on initial prevalences by behavior groups and activity levels (**Supplementary Table 9).** Initial infections were symptomatic or asymptomatic based on the initial probability of being symptomatic. For each initial infection, two infection durations were drawn from the respective duration samplers: 1) a natural clearance duration, 2) a symptomatic/asymptomatic duration depending on the symptom status of the infection. Whichever duration was shorter was selected as the duration of that infection. For the initial infections, a random day within the full duration was selected to be the time remaining infected.

##### *Transmission*

At each time step following seeding, active partnerships where one partner was infected, and the other partner was uninfected were eligible for transmission. Transmission occurred in these partnerships with the respective probability based on the partnership type (steady vs. casual) and the behavior groups of each partner. When a new infection occurred, it was symptomatic with the probability of symptoms for that agent’s behavior group. Once symptom status was determined, two durations were drawn from their respective duration distributions as above for the initial infections. We assumed that all symptomatic infections sought care and that asymptomatic infections could be detected by asymptomatic STI screening. The shorter of the two durations was selected. While an agent was infected, they were infectious, and the agent underwent testing, treatment, and recovery in the last timestep of the infection. If an agent was infected by two agents in the same timestep, one of the infections was randomly selected. Expired infections recovered at the beginning of the time step, and then transmission was simulated.

##### *Simulation run procedure*

The partnership formation simulation ran with no infections for the duration specified by partnership formation burn-in duration, after which point new infections were seeded (**Supplementary Table 10**). The simulation then ran for a transmission burn-in period, and subsequent simulation period. If all infections in the simulation period descended from one initial infector, that simulation would be used for subsequent analysis. This was to ensure that the transmission tree could be rooted at a most recent common ancestor for sequence simulation.

#### Simulated Sequence Evolution Along Transmission Chains

##### *Build a hybrid transmission tree*

All infections from the dominant lineage – which was all simulation period infections and the subset of transmission burn-in infections that descended from the initial infector – were built into a hybrid transmission tree. The goal of this step was to represent the relatedness of each infection in a format that a genome sequence simulator could recognize. This requires a) preserving the internal tree structure and b) creating a tree tip for each simulation period infection. The transmission tree therefore was structured as a typical transmission tree during the burn-in period, with a node for each transmission event. After the transmission burn-in period, the tree structure switched to daily chains. This meant that there was a node, either an internal node or a tip, for each infected day for each simulation period infection. This preserved the internal transmission structure and created a tip for each infection on the day it was sampled, which was the last day of the infection. This hybrid transmission tree was then processed into Newick format. Tree multifurcations were resolved by introducing 0 branch lengths, unary nodes were collapsed, internal node labels were removed, and burn-in period tips (dead end nodes) were removed.

##### *Simulating genome sequences*

We simulated sequences with Seq-Gen version 1.3.5^6^ using the parameters in **Supplementary Table 11**. Briefly, we simulated the number of substitutions that would occur across the *N. gonorrhea* genome in a year^7^ and adjusted that rate to the equivalent for a genome size of 10,000 bp for scalability. The resultant alignment represented what could be obtained from a recombination-masked whole-genome sequence alignment from a surveillance dataset.

### **Supplementary Figures**

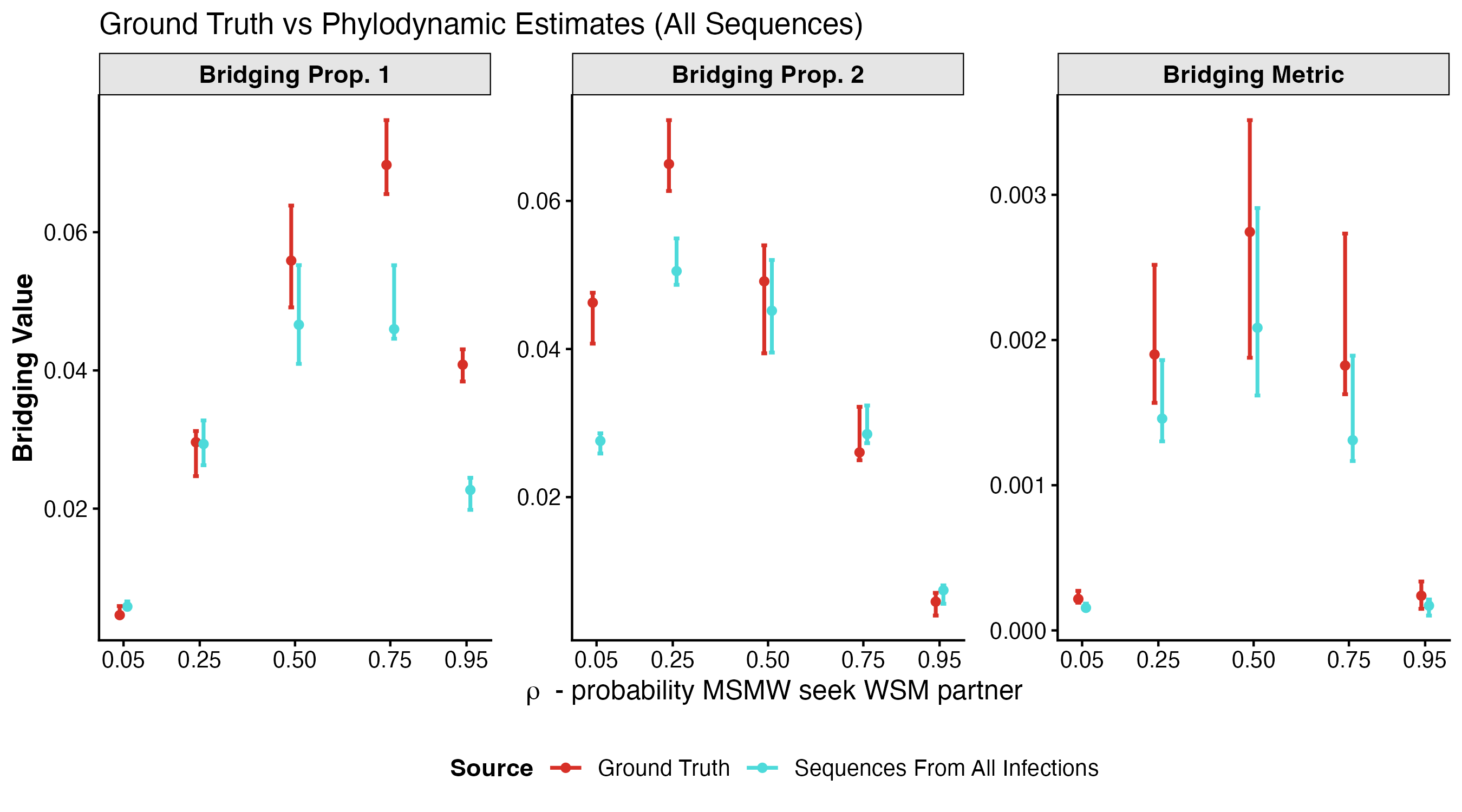

**Supplementary Figure 1:** **Phylodynamic Bridging Estimates from All Sequences.** Comparing bridging proportion 1 (left), bridging proportion 2 (middle) and the bridging metric (right) estimated phylodynamically using sequences from all infections to their respective ground truth values. Bridging proportion 1 is the proportion of between network transmissions when MSMW are grouped with MSM. Bridging proportion 2 is the proportion of between network transmissions when MSMW are grouped with MSW. The bridging metric is the product of bridging proportion 1 and bridging proportion 2. Points represent the median across 10 replicates for each value of ρ, the probability that, when seeking a new partner, and MSMW will pair with WSM. Error bars represent the interquartile range. Color and left right orientation differentiate ground truth from phylodynamic estimates using all sequences.

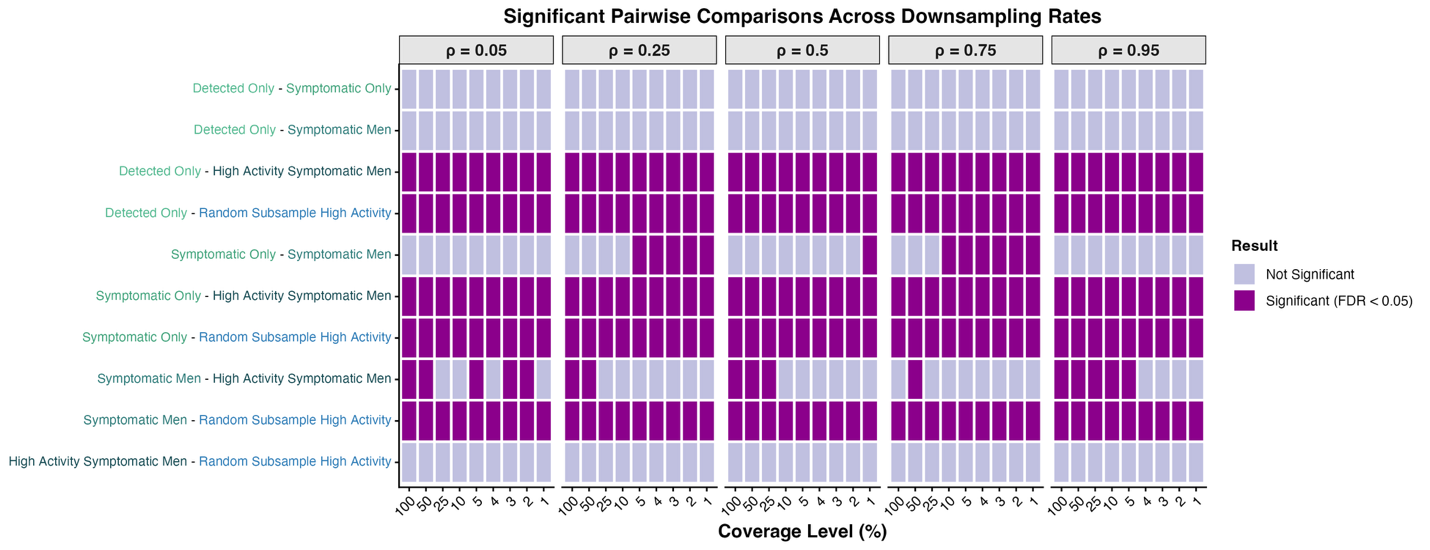

**Supplementary Figure 2: Significant difference between bridging metric estimates from sampling schemes at the same coverage level, by ρ value.** ρ represents the probability that MSMW seek female partners when they are seeking a new partner. Each box represents a pairwise Dunn test performed post-hoc following significant Kruskal-Wallis analysis showing that there were significant differences between sampling schemes at a given coverage level. Dunn test results show which of the specific sampling schemes within a coverage level are significantly different from one another. False discovery rate (FDR) was corrected for with the Benjamini-Hochberg correction.

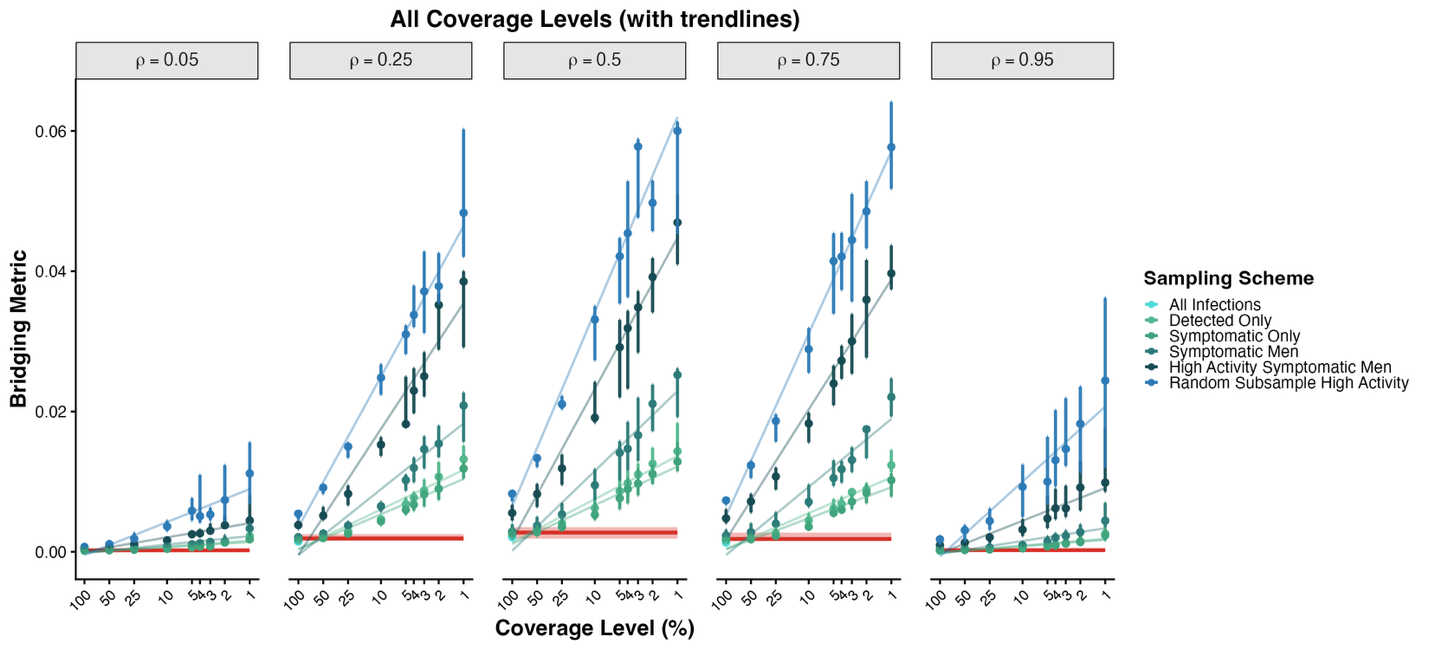

**Supplementary Figure 3**: **Phylodynamic Bridging Metric Estimates from Sampling Schemes with Incomplete Coverage, by ρ value.** ρ represents the probability that MSMW seek female partners when they are seeking a new partner. The Y-axis represents the bridging metric (point = median of medians, error bars = IQR). The X-axis shows each coverage level for each of the datasets averaged across the simulations. Colors show the ground truth compared to each of the filtered datasets representing sampling of specific subsets of the population. The horizontal line and shading show the median and IQR for the ground truth.

### **Supplementary Tables**

**Supplementary Table 1: Number of Successful Simulations and Bridging Values for Ground Truth Analysis**

| ρ | # Sims | Bridging proportion 1 - MSMW with MSM Median (IQR) | Bridging Proportion 2- MSMW with MSW Median (IQR) | Bridging Metric (product) - Median (IQR) |
| --- | --- | --- | --- | --- |
| 0.05 | 267 | 5.58E-03  (4.53E-03 - 6.97E-03) | 4.87E-02  (4.25E-02 - 5.66E-02) | 2.77E-04  (2.04E-04 - 3.75E-04) |
| 0.25 | 436 | 2.79E-02  (2.40E-02 - 3.22E-02) | 6.58E-02  (5.86E-02 - 7.37E-02) | 1.86E-03  (1.45E-03 -2.27E-03) |
| 0.5 | 464 | 5.18E-02  (4.49E-02 - 5.78E-02) | 4.84E-02  (4.31E-02 - 5.46E-02) | 2.46E-03  (1.99E-03 - 3.10E-03) |
| 0.75 | 467 | 6.72E-02  (5.92E-02 - 7.50E-02) | 2.64E-02  (2.26E-02 - 3.04E-02) | 1.76E-03  (1.35E-03 - 2.25E-03) |
| 0.95 | 401 | 4.05E-02  (3.60E-02 -4.68E-02) | 5.23E-03  (4.15E-03 - 6.64E-03) | 2.17E-04  (1.52E-04 - 2.98E-04) |

**Supplementary Table 2: Comparing all phylodynamic estimates to ground truth**

| Sampling Scheme | Coverage Level | ρ | Measure | Median Phylo (IQR) | Median Ground Truth (IQR) | Absolute Diff (IQR) | Relative to GT (IQR) | Relative to Estimate (IQR) |
| --- | --- | --- | --- | --- | --- | --- | --- | --- |
| All Sequences | 100 | 0.05 | Bridging Proportion 1 – MSMW with MSM | 5.83e-03 (5.43e-03 - 6.56e-03) | 4.59e-03 (4.59e-03 - 4.59e-03) | 1.14e-03 (1.33e-04 - 1.37e-03) | 2.16e-01 (2.19e-02 - 3.02e-01) | 1.77e-01 (1.43e-02 - 2.32e-01) |
| All Sequences | 100 | 0.25 | Bridging Proportion 1 – MSMW with MSM | 2.94e-02 (2.63e-02 - 3.28e-02) | 2.96e-02 (2.96e-02 - 2.96e-02) | 1.42e-03 (2.48e-04 - 2.09e-03) | 4.98e-02 (9.78e-03 - 7.39e-02) | 4.74e-02 (9.56e-03 - 6.88e-02) |
| All Sequences | 100 | 0.5 | Bridging Proportion 1 – MSMW with MSM | 4.66e-02 (4.09e-02 - 5.52e-02) | 5.59e-02 (5.59e-02 - 5.59e-02) | -8.53e-03 (-9.91e-03 - -7.01e-03) | -1.45e-01 (-1.65e-01 - -1.27e-01) | -1.69e-01 (-1.98e-01 - -1.46e-01) |
| All Sequences | 100 | 0.75 | Bridging Proportion 1 – MSMW with MSM | 4.60e-02 (4.46e-02 - 5.52e-02) | 6.97e-02 (6.97e-02 - 6.97e-02) | -2.21e-02 (-2.47e-02 - -2.00e-02) | -3.23e-01 (-3.35e-01 - -3.12e-01) | -4.78e-01 (-5.04e-01 - -4.53e-01) |
| All Sequences | 100 | 0.95 | Bridging Proportion 1 – MSMW with MSM | 2.27e-02 (1.98e-02 - 2.45e-02) | 4.08e-02 (4.08e-02 - 4.08e-02) | -1.83e-02 (-1.89e-02 - -1.75e-02) | -4.36e-01 (-4.54e-01 - -4.26e-01) | -7.73e-01 (-8.32e-01 - -7.43e-01) |
| All Sequences | 100 | 0.05 | Bridging Proportion 2 – MSMW with MSW | 2.76e-02 (2.59e-02 - 2.86e-02) | 4.63e-02 (4.63e-02 - 4.63e-02) | -1.78e-02 (-1.95e-02 - -1.48e-02) | -3.84e-01 (-4.10e-01 - -3.71e-01) | -6.23e-01 (-6.97e-01 - -5.89e-01) |
| All Sequences | 100 | 0.25 | Bridging Proportion 2 – MSMW with MSW | 5.05e-02 (4.87e-02 - 5.49e-02) | 6.50e-02 (6.50e-02 - 6.50e-02) | -1.49e-02 (-1.67e-02 - -1.19e-02) | -2.23e-01 (-2.38e-01 - -2.16e-01) | -2.88e-01 (-3.12e-01 - -2.75e-01) |
| All Sequences | 100 | 0.5 | Bridging Proportion 2 – MSMW with MSW | 4.52e-02 (3.95e-02 - 5.20e-02) | 4.91e-02 (4.91e-02 - 4.91e-02) | -3.46e-03 (-5.15e-03 - -8.29e-05) | -7.56e-02 (-1.01e-01 - -1.56e-03) | -8.20e-02 (-1.12e-01 - -1.57e-03) |
| All Sequences | 100 | 0.75 | Bridging Proportion 2 – MSMW with MSW | 2.85e-02 (2.73e-02 - 3.24e-02) | 2.60e-02 (2.60e-02 - 2.60e-02) | 1.06e-03 (-1.09e-04 - 1.83e-03) | 4.20e-02 (-2.42e-03 - 7.04e-02) | 4.02e-02 (-2.48e-03 - 6.57e-02) |
| All Sequences | 100 | 0.95 | Bridging Proportion 2 – MSMW with MSW | 7.42e-03 (5.60e-03 - 8.08e-03) | 5.88e-03 (5.88e-03 - 5.88e-03) | 1.32e-03 (1.14e-03 - 1.68e-03) | 2.72e-01 (2.04e-01 - 3.42e-01) | 2.14e-01 (1.69e-01 - 2.55e-01) |
| All Sequences | 100 | 0.05 | Bridging Metric (product) | 1.55e-04 (1.53e-04 - 1.84e-04) | 2.16e-04 (2.16e-04 - 2.16e-04) | -5.65e-05 (-9.62e-05 - -4.03e-05) | -2.77e-01 (-3.55e-01 - -1.85e-01) | -3.83e-01 (-5.51e-01 - -2.28e-01) |
| All Sequences | 100 | 0.25 | Bridging Metric (product) | 1.46e-03 (1.30e-03 - 1.86e-03) | 1.90e-03 (1.90e-03 - 1.90e-03) | -3.84e-04 (-4.80e-04 - -2.71e-04) | -1.85e-01 (-2.08e-01 - -1.72e-01) | -2.27e-01 (-2.62e-01 - -2.08e-01) |
| All Sequences | 100 | 0.5 | Bridging Metric (product) | 2.08e-03 (1.62e-03 - 2.91e-03) | 2.74e-03 (2.74e-03 - 2.74e-03) | -5.69e-04 (-7.67e-04 - -2.53e-04) | -1.93e-01 (-2.38e-01 - -1.41e-01) | -2.39e-01 (-3.12e-01 - -1.64e-01) |
| All Sequences | 100 | 0.75 | Bridging Metric (product) | 1.31e-03 (1.17e-03 - 1.89e-03) | 1.82e-03 (1.82e-03 - 1.82e-03) | -4.63e-04 (-7.52e-04 - -4.27e-04) | -2.80e-01 (-3.27e-01 - -2.62e-01) | -3.88e-01 (-4.87e-01 - -3.55e-01) |
| All Sequences | 100 | 0.95 | Bridging Metric (product) | 1.70e-04 (1.02e-04 - 2.13e-04) | 2.39e-04 (2.39e-04 - 2.39e-04) | -7.15e-05 (-1.07e-04 - -4.50e-05) | -3.09e-01 (-3.48e-01 - -2.43e-01) | -4.48e-01 (-5.34e-01 - -3.22e-01) |
| Detected Only | 100 | 0.05 | Bridging Metric (product) | 2.25e-04 (2.01e-04 - 2.31e-04) | 2.16e-04 (2.16e-04 - 2.16e-04) | -4.72e-06 (-3.21e-05 - 1.27e-05) | -1.63e-02 (-1.67e-01 - 6.43e-02) | -1.98e-02 (-2.04e-01 - 6.04e-02) |
| Detected Only | 100 | 0.25 | Bridging Metric (product) | 2.03e-03 (1.65e-03 - 2.36e-03) | 1.90e-03 (1.90e-03 - 1.90e-03) | 1.82e-04 (4.18e-05 - 2.23e-04) | 8.41e-02 (2.47e-02 - 1.14e-01) | 7.75e-02 (2.41e-02 - 1.02e-01) |
| Detected Only | 100 | 0.5 | Bridging Metric (product) | 2.85e-03 (2.32e-03 - 3.89e-03) | 2.74e-03 (2.74e-03 - 2.74e-03) | 1.61e-04 (1.01e-05 - 2.96e-04) | 6.86e-02 (4.19e-03 - 1.51e-01) | 6.42e-02 (4.11e-03 - 1.31e-01) |
| Detected Only | 100 | 0.75 | Bridging Metric (product) | 1.86e-03 (1.70e-03 - 2.51e-03) | 1.82e-03 (1.82e-03 - 1.82e-03) | -6.17e-05 (-1.39e-04 - 8.93e-05) | -3.21e-02 (-6.76e-02 - 5.09e-02) | -3.34e-02 (-7.26e-02 - 4.84e-02) |
| Detected Only | 100 | 0.95 | Bridging Metric (product) | 2.43e-04 (1.56e-04 - 3.12e-04) | 2.39e-04 (2.39e-04 - 2.39e-04) | 2.40e-05 (-1.78e-05 - 2.81e-05) | 8.69e-02 (-4.93e-02 - 1.71e-01) | 7.97e-02 (-5.34e-02 - 1.46e-01) |
| Detected Only | 50 | 0.05 | Bridging Metric (product) | 2.60e-04 (2.28e-04 - 2.72e-04) | 2.16e-04 (2.16e-04 - 2.16e-04) | 2.75e-05 (3.98e-06 - 4.48e-05) | 1.21e-01 (8.16e-03 - 2.07e-01) | 1.08e-01 (6.47e-03 - 1.71e-01) |
| Detected Only | 50 | 0.25 | Bridging Metric (product) | 2.29e-03 (1.98e-03 - 2.57e-03) | 1.90e-03 (1.90e-03 - 1.90e-03) | 3.96e-04 (2.84e-04 - 5.16e-04) | 2.07e-01 (1.69e-01 - 2.74e-01) | 1.71e-01 (1.44e-01 - 2.15e-01) |
| Detected Only | 50 | 0.5 | Bridging Metric (product) | 3.18e-03 (2.56e-03 - 4.23e-03) | 2.74e-03 (2.74e-03 - 2.74e-03) | 4.64e-04 (4.13e-04 - 5.83e-04) | 2.35e-01 (1.23e-01 - 3.00e-01) | 1.89e-01 (1.10e-01 - 2.31e-01) |
| Detected Only | 50 | 0.75 | Bridging Metric (product) | 2.10e-03 (1.94e-03 - 2.92e-03) | 1.82e-03 (1.82e-03 - 1.82e-03) | 2.22e-04 (7.30e-05 - 3.31e-04) | 1.12e-01 (3.30e-02 - 1.88e-01) | 9.90e-02 (3.19e-02 - 1.58e-01) |
| Detected Only | 50 | 0.95 | Bridging Metric (product) | 2.90e-04 (1.92e-04 - 3.83e-04) | 2.39e-04 (2.39e-04 - 2.39e-04) | 5.67e-05 (3.51e-05 - 7.11e-05) | 2.41e-01 (1.60e-01 - 3.49e-01) | 1.94e-01 (1.38e-01 - 2.58e-01) |
| Detected Only | 25 | 0.05 | Bridging Metric (product) | 3.22e-04 (2.66e-04 - 3.48e-04) | 2.16e-04 (2.16e-04 - 2.16e-04) | 8.34e-05 (6.37e-05 - 9.92e-05) | 3.60e-01 (2.09e-01 - 4.98e-01) | 2.64e-01 (1.73e-01 - 3.32e-01) |
| Detected Only | 25 | 0.25 | Bridging Metric (product) | 2.91e-03 (2.54e-03 - 3.27e-03) | 1.90e-03 (1.90e-03 - 1.90e-03) | 9.68e-04 (8.59e-04 - 1.14e-03) | 5.28e-01 (4.59e-01 - 6.03e-01) | 3.46e-01 (3.15e-01 - 3.76e-01) |
| Detected Only | 25 | 0.5 | Bridging Metric (product) | 4.06e-03 (3.27e-03 - 5.22e-03) | 2.74e-03 (2.74e-03 - 2.74e-03) | 1.26e-03 (1.10e-03 - 1.56e-03) | 5.56e-01 (4.08e-01 - 5.86e-01) | 3.57e-01 (2.90e-01 - 3.69e-01) |
| Detected Only | 25 | 0.75 | Bridging Metric (product) | 2.65e-03 (2.44e-03 - 3.71e-03) | 1.82e-03 (1.82e-03 - 1.82e-03) | 7.31e-04 (6.02e-04 - 1.02e-03) | 3.95e-01 (3.19e-01 - 4.65e-01) | 2.83e-01 (2.41e-01 - 3.17e-01) |
| Detected Only | 25 | 0.95 | Bridging Metric (product) | 3.81e-04 (2.53e-04 - 5.17e-04) | 2.39e-04 (2.39e-04 - 2.39e-04) | 1.16e-04 (1.02e-04 - 1.95e-04) | 7.17e-01 (6.09e-01 - 7.71e-01) | 4.18e-01 (3.78e-01 - 4.35e-01) |
| Detected Only | 10 | 0.05 | Bridging Metric (product) | 5.31e-04 (4.07e-04 - 6.14e-04) | 2.16e-04 (2.16e-04 - 2.16e-04) | 2.33e-04 (1.93e-04 - 3.61e-04) | 1.10e+00 (8.80e-01 - 1.49e+00) | 5.22e-01 (4.68e-01 - 5.97e-01) |
| Detected Only | 10 | 0.25 | Bridging Metric (product) | 4.62e-03 (3.78e-03 - 5.16e-03) | 1.90e-03 (1.90e-03 - 1.90e-03) | 2.55e-03 (2.18e-03 - 2.88e-03) | 1.43e+00 (1.38e+00 - 1.49e+00) | 5.88e-01 (5.80e-01 - 5.99e-01) |
| Detected Only | 10 | 0.5 | Bridging Metric (product) | 6.22e-03 (5.07e-03 - 7.48e-03) | 2.74e-03 (2.74e-03 - 2.74e-03) | 3.50e-03 (3.04e-03 - 4.02e-03) | 1.43e+00 (1.10e+00 - 1.44e+00) | 5.88e-01 (5.24e-01 - 5.91e-01) |
| Detected Only | 10 | 0.75 | Bridging Metric (product) | 4.46e-03 (4.08e-03 - 5.48e-03) | 1.82e-03 (1.82e-03 - 1.82e-03) | 2.74e-03 (2.19e-03 - 3.10e-03) | 1.43e+00 (1.07e+00 - 1.77e+00) | 5.87e-01 (5.16e-01 - 6.39e-01) |
| Detected Only | 10 | 0.95 | Bridging Metric (product) | 6.17e-04 (3.89e-04 - 8.06e-04) | 2.39e-04 (2.39e-04 - 2.39e-04) | 3.23e-04 (2.62e-04 - 4.99e-04) | 1.64e+00 (1.39e+00 - 1.97e+00) | 6.21e-01 (5.81e-01 - 6.63e-01) |
| Detected Only | 5 | 0.05 | Bridging Metric (product) | 6.37e-04 (5.43e-04 - 8.40e-04) | 2.16e-04 (2.16e-04 - 2.16e-04) | 4.18e-04 (2.84e-04 - 6.23e-04) | 1.88e+00 (1.17e+00 - 3.42e+00) | 6.48e-01 (5.37e-01 - 7.72e-01) |
| Detected Only | 5 | 0.25 | Bridging Metric (product) | 6.78e-03 (6.07e-03 - 7.70e-03) | 1.90e-03 (1.90e-03 - 1.90e-03) | 4.70e-03 (4.40e-03 - 5.53e-03) | 2.70e+00 (2.44e+00 - 2.77e+00) | 7.30e-01 (7.09e-01 - 7.35e-01) |
| Detected Only | 5 | 0.5 | Bridging Metric (product) | 8.64e-03 (6.84e-03 - 1.04e-02) | 2.74e-03 (2.74e-03 - 2.74e-03) | 6.00e-03 (4.93e-03 - 6.63e-03) | 2.30e+00 (1.97e+00 - 2.49e+00) | 6.97e-01 (6.64e-01 - 7.13e-01) |
| Detected Only | 5 | 0.75 | Bridging Metric (product) | 6.19e-03 (5.67e-03 - 7.51e-03) | 1.82e-03 (1.82e-03 - 1.82e-03) | 4.22e-03 (3.72e-03 - 5.17e-03) | 2.36e+00 (2.03e+00 - 2.51e+00) | 7.03e-01 (6.69e-01 - 7.15e-01) |
| Detected Only | 5 | 0.95 | Bridging Metric (product) | 9.25e-04 (6.50e-04 - 1.38e-03) | 2.39e-04 (2.39e-04 - 2.39e-04) | 6.99e-04 (4.95e-04 - 1.07e-03) | 3.39e+00 (2.88e+00 - 3.48e+00) | 7.72e-01 (7.41e-01 - 7.77e-01) |
| Detected Only | 4 | 0.05 | Bridging Metric (product) | 7.40e-04 (6.61e-04 - 9.49e-04) | 2.16e-04 (2.16e-04 - 2.16e-04) | 5.53e-04 (3.97e-04 - 7.00e-04) | 2.72e+00 (1.58e+00 - 3.53e+00) | 7.31e-01 (6.10e-01 - 7.79e-01) |
| Detected Only | 4 | 0.25 | Bridging Metric (product) | 7.73e-03 (6.61e-03 - 8.52e-03) | 1.90e-03 (1.90e-03 - 1.90e-03) | 5.60e-03 (4.97e-03 - 6.42e-03) | 3.07e+00 (2.70e+00 - 3.28e+00) | 7.54e-01 (7.29e-01 - 7.66e-01) |
| Detected Only | 4 | 0.5 | Bridging Metric (product) | 9.78e-03 (7.65e-03 - 1.16e-02) | 2.74e-03 (2.74e-03 - 2.74e-03) | 6.91e-03 (5.73e-03 - 7.91e-03) | 2.56e+00 (2.34e+00 - 2.88e+00) | 7.19e-01 (7.00e-01 - 7.42e-01) |
| Detected Only | 4 | 0.75 | Bridging Metric (product) | 6.82e-03 (6.36e-03 - 7.81e-03) | 1.82e-03 (1.82e-03 - 1.82e-03) | 5.06e-03 (4.41e-03 - 5.61e-03) | 2.86e+00 (2.53e+00 - 2.96e+00) | 7.41e-01 (7.17e-01 - 7.48e-01) |
| Detected Only | 4 | 0.95 | Bridging Metric (product) | 9.92e-04 (7.95e-04 - 1.33e-03) | 2.39e-04 (2.39e-04 - 2.39e-04) | 7.60e-04 (6.26e-04 - 1.02e-03) | 3.75e+00 (2.90e+00 - 5.65e+00) | 7.90e-01 (7.44e-01 - 8.45e-01) |
| Detected Only | 3 | 0.05 | Bridging Metric (product) | 1.07e-03 (7.84e-04 - 1.16e-03) | 2.16e-04 (2.16e-04 - 2.16e-04) | 8.23e-04 (5.39e-04 - 9.41e-04) | 3.62e+00 (2.32e+00 - 4.61e+00) | 7.82e-01 (6.99e-01 - 8.21e-01) |
| Detected Only | 3 | 0.25 | Bridging Metric (product) | 8.72e-03 (7.27e-03 - 1.03e-02) | 1.90e-03 (1.90e-03 - 1.90e-03) | 6.43e-03 (5.69e-03 - 8.16e-03) | 3.71e+00 (3.58e+00 - 3.85e+00) | 7.88e-01 (7.82e-01 - 7.94e-01) |
| Detected Only | 3 | 0.5 | Bridging Metric (product) | 1.10e-02 (8.35e-03 - 1.26e-02) | 2.74e-03 (2.74e-03 - 2.74e-03) | 7.78e-03 (6.64e-03 - 9.52e-03) | 3.14e+00 (2.66e+00 - 3.84e+00) | 7.58e-01 (7.26e-01 - 7.94e-01) |
| Detected Only | 3 | 0.75 | Bridging Metric (product) | 8.50e-03 (7.23e-03 - 8.93e-03) | 1.82e-03 (1.82e-03 - 1.82e-03) | 6.11e-03 (5.39e-03 - 7.24e-03) | 3.12e+00 (2.99e+00 - 3.77e+00) | 7.57e-01 (7.49e-01 - 7.90e-01) |
| Detected Only | 3 | 0.95 | Bridging Metric (product) | 1.23e-03 (9.21e-04 - 1.79e-03) | 2.39e-04 (2.39e-04 - 2.39e-04) | 9.45e-04 (7.94e-04 - 1.47e-03) | 4.85e+00 (4.10e+00 - 6.74e+00) | 8.29e-01 (8.04e-01 - 8.70e-01) |
| Detected Only | 2 | 0.05 | Bridging Metric (product) | 1.36e-03 (1.23e-03 - 1.50e-03) | 2.16e-04 (2.16e-04 - 2.16e-04) | 1.10e-03 (9.28e-04 - 1.27e-03) | 4.49e+00 (3.42e+00 - 6.11e+00) | 8.18e-01 (7.73e-01 - 8.59e-01) |
| Detected Only | 2 | 0.25 | Bridging Metric (product) | 1.07e-02 (7.62e-03 - 1.28e-02) | 1.90e-03 (1.90e-03 - 1.90e-03) | 8.81e-03 (6.05e-03 - 1.02e-02) | 4.35e+00 (3.92e+00 - 4.75e+00) | 8.13e-01 (7.97e-01 - 8.26e-01) |
| Detected Only | 2 | 0.5 | Bridging Metric (product) | 1.26e-02 (1.00e-02 - 1.48e-02) | 2.74e-03 (2.74e-03 - 2.74e-03) | 1.01e-02 (8.03e-03 - 1.07e-02) | 3.69e+00 (3.25e+00 - 4.07e+00) | 7.87e-01 (7.65e-01 - 8.03e-01) |
| Detected Only | 2 | 0.75 | Bridging Metric (product) | 8.86e-03 (8.67e-03 - 9.81e-03) | 1.82e-03 (1.82e-03 - 1.82e-03) | 7.44e-03 (6.66e-03 - 8.03e-03) | 4.02e+00 (3.06e+00 - 4.41e+00) | 8.01e-01 (7.53e-01 - 8.15e-01) |
| Detected Only | 2 | 0.95 | Bridging Metric (product) | 1.50e-03 (1.06e-03 - 2.15e-03) | 2.39e-04 (2.39e-04 - 2.39e-04) | 1.24e-03 (8.97e-04 - 1.88e-03) | 6.30e+00 (4.49e+00 - 7.61e+00) | 8.63e-01 (8.18e-01 - 8.84e-01) |
| Detected Only | 1 | 0.05 | Bridging Metric (product) | 2.25e-03 (1.86e-03 - 2.90e-03) | 2.16e-04 (2.16e-04 - 2.16e-04) | 2.05e-03 (1.55e-03 - 2.65e-03) | 9.86e+00 (6.02e+00 - 1.16e+01) | 9.08e-01 (8.51e-01 - 9.20e-01) |
| Detected Only | 1 | 0.25 | Bridging Metric (product) | 1.32e-02 (1.23e-02 - 1.51e-02) | 1.90e-03 (1.90e-03 - 1.90e-03) | 1.11e-02 (1.06e-02 - 1.31e-02) | 6.27e+00 (5.51e+00 - 6.84e+00) | 8.62e-01 (8.46e-01 - 8.72e-01) |
| Detected Only | 1 | 0.5 | Bridging Metric (product) | 1.43e-02 (1.38e-02 - 1.83e-02) | 2.74e-03 (2.74e-03 - 2.74e-03) | 1.23e-02 (1.04e-02 - 1.55e-02) | 5.08e+00 (3.90e+00 - 6.68e+00) | 8.34e-01 (7.96e-01 - 8.70e-01) |
| Detected Only | 1 | 0.75 | Bridging Metric (product) | 1.23e-02 (1.13e-02 - 1.45e-02) | 1.82e-03 (1.82e-03 - 1.82e-03) | 1.07e-02 (1.00e-02 - 1.19e-02) | 6.11e+00 (4.71e+00 - 6.92e+00) | 8.59e-01 (8.25e-01 - 8.74e-01) |
| Detected Only | 1 | 0.95 | Bridging Metric (product) | 2.55e-03 (1.82e-03 - 3.55e-03) | 2.39e-04 (2.39e-04 - 2.39e-04) | 2.38e-03 (1.61e-03 - 3.21e-03) | 9.95e+00 (8.97e+00 - 1.10e+01) | 9.09e-01 (9.00e-01 - 9.17e-01) |
| Symptomatic Only | 100 | 0.05 | Bridging Metric (product) | 1.92e-04 (1.70e-04 - 1.97e-04) | 2.16e-04 (2.16e-04 - 2.16e-04) | -3.12e-05 (-5.50e-05 - -1.55e-05) | -1.21e-01 (-2.94e-01 - -9.10e-02) | -1.38e-01 (-4.21e-01 - -1.00e-01) |
| Symptomatic Only | 100 | 0.25 | Bridging Metric (product) | 1.71e-03 (1.46e-03 - 2.06e-03) | 1.90e-03 (1.90e-03 - 1.90e-03) | -1.22e-04 (-1.85e-04 - -6.24e-05) | -6.77e-02 (-9.31e-02 - -2.95e-02) | -7.27e-02 (-1.03e-01 - -3.05e-02) |
| Symptomatic Only | 100 | 0.5 | Bridging Metric (product) | 2.49e-03 (1.85e-03 - 3.32e-03) | 2.74e-03 (2.74e-03 - 2.74e-03) | -1.31e-04 (-4.15e-04 - -2.67e-05) | -6.11e-02 (-1.37e-01 - -1.42e-02) | -6.54e-02 (-1.59e-01 - -1.44e-02) |
| Symptomatic Only | 100 | 0.75 | Bridging Metric (product) | 1.65e-03 (1.49e-03 - 2.15e-03) | 1.82e-03 (1.82e-03 - 1.82e-03) | -2.33e-04 (-3.88e-04 - -1.82e-04) | -1.19e-01 (-2.23e-01 - -9.83e-02) | -1.36e-01 (-2.88e-01 - -1.09e-01) |
| Symptomatic Only | 100 | 0.95 | Bridging Metric (product) | 2.01e-04 (1.24e-04 - 2.74e-04) | 2.39e-04 (2.39e-04 - 2.39e-04) | -3.12e-05 (-5.65e-05 - -7.95e-06) | -1.03e-01 (-2.07e-01 - -5.77e-02) | -1.15e-01 (-2.61e-01 - -6.18e-02) |
| Symptomatic Only | 50 | 0.05 | Bridging Metric (product) | 2.26e-04 (1.99e-04 - 2.35e-04) | 2.16e-04 (2.16e-04 - 2.16e-04) | -4.39e-06 (-1.71e-05 - 1.54e-05) | -1.15e-02 (-1.13e-01 - 7.14e-02) | -1.18e-02 (-1.28e-01 - 6.55e-02) |
| Symptomatic Only | 50 | 0.25 | Bridging Metric (product) | 1.96e-03 (1.68e-03 - 2.31e-03) | 1.90e-03 (1.90e-03 - 1.90e-03) | 7.92e-05 (3.93e-05 - 1.87e-04) | 3.50e-02 (2.45e-02 - 8.45e-02) | 3.38e-02 (2.39e-02 - 7.79e-02) |
| Symptomatic Only | 50 | 0.5 | Bridging Metric (product) | 2.82e-03 (2.22e-03 - 3.65e-03) | 2.74e-03 (2.74e-03 - 2.74e-03) | 1.45e-04 (-5.74e-05 - 2.69e-04) | 7.92e-02 (-2.16e-02 - 1.24e-01) | 7.33e-02 (-2.21e-02 - 1.10e-01) |
| Symptomatic Only | 50 | 0.75 | Bridging Metric (product) | 1.77e-03 (1.65e-03 - 2.45e-03) | 1.82e-03 (1.82e-03 - 1.82e-03) | -1.11e-04 (-1.77e-04 - 1.63e-05) | -4.98e-02 (-1.00e-01 - 7.64e-03) | -5.25e-02 (-1.12e-01 - 6.89e-03) |
| Symptomatic Only | 50 | 0.95 | Bridging Metric (product) | 2.52e-04 (1.57e-04 - 3.43e-04) | 2.39e-04 (2.39e-04 - 2.39e-04) | 2.11e-05 (3.83e-06 - 3.26e-05) | 6.83e-02 (2.32e-02 - 1.97e-01) | 6.38e-02 (2.27e-02 - 1.63e-01) |
| Symptomatic Only | 25 | 0.05 | Bridging Metric (product) | 2.89e-04 (2.36e-04 - 3.05e-04) | 2.16e-04 (2.16e-04 - 2.16e-04) | 6.10e-05 (1.42e-05 - 8.23e-05) | 3.02e-01 (8.47e-02 - 3.67e-01) | 2.32e-01 (7.68e-02 - 2.68e-01) |
| Symptomatic Only | 25 | 0.25 | Bridging Metric (product) | 2.59e-03 (2.33e-03 - 2.99e-03) | 1.90e-03 (1.90e-03 - 1.90e-03) | 7.76e-04 (5.51e-04 - 8.36e-04) | 3.86e-01 (3.10e-01 - 4.75e-01) | 2.78e-01 (2.37e-01 - 3.22e-01) |
| Symptomatic Only | 25 | 0.5 | Bridging Metric (product) | 3.55e-03 (2.96e-03 - 4.62e-03) | 2.74e-03 (2.74e-03 - 2.74e-03) | 9.08e-04 (7.17e-04 - 1.14e-03) | 4.16e-01 (2.56e-01 - 4.57e-01) | 2.94e-01 (2.04e-01 - 3.14e-01) |
| Symptomatic Only | 25 | 0.75 | Bridging Metric (product) | 2.28e-03 (2.15e-03 - 3.20e-03) | 1.82e-03 (1.82e-03 - 1.82e-03) | 4.65e-04 (2.94e-04 - 6.45e-04) | 2.45e-01 (1.35e-01 - 2.97e-01) | 1.97e-01 (1.19e-01 - 2.29e-01) |
| Symptomatic Only | 25 | 0.95 | Bridging Metric (product) | 3.27e-04 (2.06e-04 - 4.40e-04) | 2.39e-04 (2.39e-04 - 2.39e-04) | 9.50e-05 (5.53e-05 - 1.32e-04) | 3.72e-01 (2.59e-01 - 6.03e-01) | 2.71e-01 (2.05e-01 - 3.73e-01) |
| Symptomatic Only | 10 | 0.05 | Bridging Metric (product) | 4.20e-04 (3.71e-04 - 5.47e-04) | 2.16e-04 (2.16e-04 - 2.16e-04) | 2.28e-04 (1.31e-04 - 2.92e-04) | 8.11e-01 (5.49e-01 - 1.53e+00) | 4.47e-01 (3.53e-01 - 6.03e-01) |
| Symptomatic Only | 10 | 0.25 | Bridging Metric (product) | 4.37e-03 (3.78e-03 - 4.64e-03) | 1.90e-03 (1.90e-03 - 1.90e-03) | 2.38e-03 (2.04e-03 - 2.47e-03) | 1.26e+00 (1.15e+00 - 1.36e+00) | 5.57e-01 (5.35e-01 - 5.76e-01) |
| Symptomatic Only | 10 | 0.5 | Bridging Metric (product) | 5.28e-03 (4.56e-03 - 7.03e-03) | 2.74e-03 (2.74e-03 - 2.74e-03) | 2.76e-03 (2.34e-03 - 3.37e-03) | 1.07e+00 (9.00e-01 - 1.31e+00) | 5.17e-01 (4.74e-01 - 5.67e-01) |
| Symptomatic Only | 10 | 0.75 | Bridging Metric (product) | 3.58e-03 (3.15e-03 - 4.57e-03) | 1.82e-03 (1.82e-03 - 1.82e-03) | 1.60e-03 (1.27e-03 - 1.97e-03) | 8.57e-01 (6.86e-01 - 1.13e+00) | 4.62e-01 (4.06e-01 - 5.30e-01) |
| Symptomatic Only | 10 | 0.95 | Bridging Metric (product) | 4.91e-04 (3.59e-04 - 7.30e-04) | 2.39e-04 (2.39e-04 - 2.39e-04) | 2.63e-04 (2.31e-04 - 4.01e-04) | 1.19e+00 (1.02e+00 - 1.70e+00) | 5.44e-01 (5.06e-01 - 6.28e-01) |
| Symptomatic Only | 5 | 0.05 | Bridging Metric (product) | 7.31e-04 (6.57e-04 - 8.27e-04) | 2.16e-04 (2.16e-04 - 2.16e-04) | 4.90e-04 (3.91e-04 - 5.93e-04) | 2.49e+00 (1.46e+00 - 2.86e+00) | 7.14e-01 (5.88e-01 - 7.41e-01) |
| Symptomatic Only | 5 | 0.25 | Bridging Metric (product) | 5.99e-03 (5.29e-03 - 6.57e-03) | 1.90e-03 (1.90e-03 - 1.90e-03) | 4.04e-03 (3.71e-03 - 4.10e-03) | 2.25e+00 (1.94e+00 - 2.41e+00) | 6.92e-01 (6.60e-01 - 7.07e-01) |
| Symptomatic Only | 5 | 0.5 | Bridging Metric (product) | 7.64e-03 (6.08e-03 - 9.20e-03) | 2.74e-03 (2.74e-03 - 2.74e-03) | 4.80e-03 (4.15e-03 - 5.81e-03) | 1.97e+00 (1.64e+00 - 2.12e+00) | 6.63e-01 (6.21e-01 - 6.79e-01) |
| Symptomatic Only | 5 | 0.75 | Bridging Metric (product) | 5.54e-03 (5.07e-03 - 6.48e-03) | 1.82e-03 (1.82e-03 - 1.82e-03) | 3.50e-03 (3.15e-03 - 3.92e-03) | 1.95e+00 (1.59e+00 - 2.09e+00) | 6.60e-01 (6.13e-01 - 6.77e-01) |
| Symptomatic Only | 5 | 0.95 | Bridging Metric (product) | 7.65e-04 (5.87e-04 - 1.04e-03) | 2.39e-04 (2.39e-04 - 2.39e-04) | 5.42e-04 (4.47e-04 - 7.17e-04) | 2.50e+00 (2.17e+00 - 3.35e+00) | 7.14e-01 (6.84e-01 - 7.68e-01) |
| Symptomatic Only | 4 | 0.05 | Bridging Metric (product) | 7.91e-04 (5.93e-04 - 9.31e-04) | 2.16e-04 (2.16e-04 - 2.16e-04) | 4.57e-04 (3.82e-04 - 7.21e-04) | 2.46e+00 (1.53e+00 - 3.47e+00) | 7.09e-01 (6.05e-01 - 7.76e-01) |
| Symptomatic Only | 4 | 0.25 | Bridging Metric (product) | 6.73e-03 (5.86e-03 - 7.89e-03) | 1.90e-03 (1.90e-03 - 1.90e-03) | 4.73e-03 (3.95e-03 - 5.61e-03) | 2.69e+00 (2.66e+00 - 2.83e+00) | 7.29e-01 (7.27e-01 - 7.39e-01) |
| Symptomatic Only | 4 | 0.5 | Bridging Metric (product) | 8.93e-03 (6.91e-03 - 1.06e-02) | 2.74e-03 (2.74e-03 - 2.74e-03) | 5.61e-03 (4.88e-03 - 6.91e-03) | 2.32e+00 (1.99e+00 - 2.88e+00) | 6.98e-01 (6.65e-01 - 7.42e-01) |
| Symptomatic Only | 4 | 0.75 | Bridging Metric (product) | 6.00e-03 (5.57e-03 - 6.54e-03) | 1.82e-03 (1.82e-03 - 1.82e-03) | 3.85e-03 (3.48e-03 - 4.37e-03) | 2.32e+00 (1.98e+00 - 2.63e+00) | 6.99e-01 (6.64e-01 - 7.25e-01) |
| Symptomatic Only | 4 | 0.95 | Bridging Metric (product) | 9.05e-04 (6.38e-04 - 1.25e-03) | 2.39e-04 (2.39e-04 - 2.39e-04) | 6.47e-04 (4.85e-04 - 9.26e-04) | 3.38e+00 (2.29e+00 - 4.53e+00) | 7.70e-01 (6.93e-01 - 8.19e-01) |
| Symptomatic Only | 3 | 0.05 | Bridging Metric (product) | 7.80e-04 (7.31e-04 - 1.05e-03) | 2.16e-04 (2.16e-04 - 2.16e-04) | 5.15e-04 (4.83e-04 - 8.49e-04) | 3.04e+00 (1.76e+00 - 4.18e+00) | 7.46e-01 (6.34e-01 - 8.07e-01) |
| Symptomatic Only | 3 | 0.25 | Bridging Metric (product) | 8.19e-03 (6.48e-03 - 8.72e-03) | 1.90e-03 (1.90e-03 - 1.90e-03) | 5.96e-03 (5.09e-03 - 6.74e-03) | 3.35e+00 (2.82e+00 - 3.66e+00) | 7.70e-01 (7.38e-01 - 7.85e-01) |
| Symptomatic Only | 3 | 0.5 | Bridging Metric (product) | 9.73e-03 (8.07e-03 - 1.18e-02) | 2.74e-03 (2.74e-03 - 2.74e-03) | 6.84e-03 (5.65e-03 - 8.46e-03) | 2.62e+00 (2.27e+00 - 3.03e+00) | 7.23e-01 (6.95e-01 - 7.51e-01) |
| Symptomatic Only | 3 | 0.75 | Bridging Metric (product) | 7.15e-03 (6.11e-03 - 8.18e-03) | 1.82e-03 (1.82e-03 - 1.82e-03) | 5.18e-03 (4.04e-03 - 5.88e-03) | 2.79e+00 (2.19e+00 - 3.24e+00) | 7.36e-01 (6.86e-01 - 7.64e-01) |
| Symptomatic Only | 3 | 0.95 | Bridging Metric (product) | 1.17e-03 (9.99e-04 - 1.63e-03) | 2.39e-04 (2.39e-04 - 2.39e-04) | 9.50e-04 (8.45e-04 - 1.32e-03) | 4.43e+00 (3.56e+00 - 5.37e+00) | 8.16e-01 (7.79e-01 - 8.43e-01) |
| Symptomatic Only | 2 | 0.05 | Bridging Metric (product) | 1.32e-03 (9.89e-04 - 1.66e-03) | 2.16e-04 (2.16e-04 - 2.16e-04) | 1.03e-03 (7.08e-04 - 1.46e-03) | 4.79e+00 (2.67e+00 - 6.40e+00) | 8.27e-01 (7.25e-01 - 8.65e-01) |
| Symptomatic Only | 2 | 0.25 | Bridging Metric (product) | 8.98e-03 (7.41e-03 - 1.10e-02) | 1.90e-03 (1.90e-03 - 1.90e-03) | 7.11e-03 (5.76e-03 - 8.54e-03) | 3.86e+00 (3.53e+00 - 4.34e+00) | 7.94e-01 (7.79e-01 - 8.13e-01) |
| Symptomatic Only | 2 | 0.5 | Bridging Metric (product) | 1.11e-02 (9.68e-03 - 1.24e-02) | 2.74e-03 (2.74e-03 - 2.74e-03) | 7.97e-03 (6.98e-03 - 9.51e-03) | 3.04e+00 (2.92e+00 - 3.79e+00) | 7.52e-01 (7.45e-01 - 7.91e-01) |
| Symptomatic Only | 2 | 0.75 | Bridging Metric (product) | 8.41e-03 (6.92e-03 - 9.58e-03) | 1.82e-03 (1.82e-03 - 1.82e-03) | 5.96e-03 (5.49e-03 - 7.82e-03) | 3.37e+00 (2.61e+00 - 4.45e+00) | 7.71e-01 (7.23e-01 - 8.16e-01) |
| Symptomatic Only | 2 | 0.95 | Bridging Metric (product) | 1.39e-03 (1.03e-03 - 2.09e-03) | 2.39e-04 (2.39e-04 - 2.39e-04) | 1.18e-03 (8.53e-04 - 1.82e-03) | 6.11e+00 (3.61e+00 - 7.43e+00) | 8.59e-01 (7.79e-01 - 8.81e-01) |
| Symptomatic Only | 1 | 0.05 | Bridging Metric (product) | 1.77e-03 (1.54e-03 - 2.35e-03) | 2.16e-04 (2.16e-04 - 2.16e-04) | 1.57e-03 (1.23e-03 - 2.17e-03) | 8.78e+00 (4.60e+00 - 1.13e+01) | 8.97e-01 (8.21e-01 - 9.19e-01) |
| Symptomatic Only | 1 | 0.25 | Bridging Metric (product) | 1.19e-02 (1.05e-02 - 1.28e-02) | 1.90e-03 (1.90e-03 - 1.90e-03) | 9.86e-03 (9.09e-03 - 1.07e-02) | 4.94e+00 (4.50e+00 - 6.14e+00) | 8.32e-01 (8.18e-01 - 8.60e-01) |
| Symptomatic Only | 1 | 0.5 | Bridging Metric (product) | 1.29e-02 (1.15e-02 - 1.56e-02) | 2.74e-03 (2.74e-03 - 2.74e-03) | 1.03e-02 (9.35e-03 - 1.22e-02) | 4.06e+00 (3.79e+00 - 4.63e+00) | 8.02e-01 (7.91e-01 - 8.22e-01) |
| Symptomatic Only | 1 | 0.75 | Bridging Metric (product) | 1.02e-02 (7.88e-03 - 1.12e-02) | 1.82e-03 (1.82e-03 - 1.82e-03) | 8.29e-03 (6.01e-03 - 8.77e-03) | 4.21e+00 (3.04e+00 - 4.92e+00) | 8.06e-01 (7.52e-01 - 8.31e-01) |
| Symptomatic Only | 1 | 0.95 | Bridging Metric (product) | 2.37e-03 (1.62e-03 - 3.20e-03) | 2.39e-04 (2.39e-04 - 2.39e-04) | 2.19e-03 (1.46e-03 - 2.86e-03) | 9.53e+00 (8.05e+00 - 1.32e+01) | 9.04e-01 (8.89e-01 - 9.29e-01) |
| Symptomatic Men | 100 | 0.05 | Bridging Metric (product) | 2.41e-04 (1.95e-04 - 2.56e-04) | 2.16e-04 (2.16e-04 - 2.16e-04) | 1.62e-05 (-2.49e-05 - 3.69e-05) | 7.38e-02 (-1.23e-01 - 1.71e-01) | 6.66e-02 (-1.42e-01 - 1.45e-01) |
| Symptomatic Men | 100 | 0.25 | Bridging Metric (product) | 2.09e-03 (1.82e-03 - 2.39e-03) | 1.90e-03 (1.90e-03 - 1.90e-03) | 2.70e-04 (2.13e-04 - 2.86e-04) | 1.34e-01 (1.00e-01 - 2.30e-01) | 1.18e-01 (9.09e-02 - 1.86e-01) |
| Symptomatic Men | 100 | 0.5 | Bridging Metric (product) | 2.88e-03 (2.22e-03 - 3.91e-03) | 2.74e-03 (2.74e-03 - 2.74e-03) | 2.69e-04 (-4.94e-05 - 4.82e-04) | 1.71e-01 (-5.00e-03 - 2.03e-01) | 1.46e-01 (-6.87e-03 - 1.69e-01) |
| Symptomatic Men | 100 | 0.75 | Bridging Metric (product) | 2.33e-03 (1.94e-03 - 3.13e-03) | 1.82e-03 (1.82e-03 - 1.82e-03) | 3.57e-04 (3.17e-04 - 5.90e-04) | 1.97e-01 (1.26e-01 - 2.99e-01) | 1.65e-01 (1.11e-01 - 2.30e-01) |
| Symptomatic Men | 100 | 0.95 | Bridging Metric (product) | 3.37e-04 (2.05e-04 - 3.99e-04) | 2.39e-04 (2.39e-04 - 2.39e-04) | 6.67e-05 (5.72e-05 - 1.18e-04) | 3.90e-01 (2.45e-01 - 5.61e-01) | 2.81e-01 (1.97e-01 - 3.57e-01) |
| Symptomatic Men | 50 | 0.05 | Bridging Metric (product) | 3.13e-04 (2.33e-04 - 3.46e-04) | 2.16e-04 (2.16e-04 - 2.16e-04) | 8.49e-05 (4.11e-05 - 1.01e-04) | 4.21e-01 (2.09e-01 - 4.60e-01) | 2.96e-01 (1.73e-01 - 3.15e-01) |
| Symptomatic Men | 50 | 0.25 | Bridging Metric (product) | 2.59e-03 (2.36e-03 - 3.05e-03) | 1.90e-03 (1.90e-03 - 1.90e-03) | 8.10e-04 (5.35e-04 - 9.49e-04) | 4.39e-01 (2.93e-01 - 5.58e-01) | 3.05e-01 (2.26e-01 - 3.58e-01) |
| Symptomatic Men | 50 | 0.5 | Bridging Metric (product) | 3.73e-03 (2.82e-03 - 4.95e-03) | 2.74e-03 (2.74e-03 - 2.74e-03) | 8.86e-04 (8.45e-04 - 1.07e-03) | 4.49e-01 (2.92e-01 - 4.83e-01) | 3.10e-01 (2.26e-01 - 3.26e-01) |
| Symptomatic Men | 50 | 0.75 | Bridging Metric (product) | 2.80e-03 (2.52e-03 - 4.04e-03) | 1.82e-03 (1.82e-03 - 1.82e-03) | 1.09e-03 (9.28e-04 - 1.27e-03) | 4.96e-01 (4.25e-01 - 6.01e-01) | 3.31e-01 (2.98e-01 - 3.75e-01) |
| Symptomatic Men | 50 | 0.95 | Bridging Metric (product) | 4.45e-04 (2.88e-04 - 5.41e-04) | 2.39e-04 (2.39e-04 - 2.39e-04) | 1.64e-04 (1.41e-04 - 2.42e-04) | 8.59e-01 (6.62e-01 - 1.09e+00) | 4.62e-01 (3.98e-01 - 5.21e-01) |
| Symptomatic Men | 25 | 0.05 | Bridging Metric (product) | 4.11e-04 (3.42e-04 - 5.12e-04) | 2.16e-04 (2.16e-04 - 2.16e-04) | 1.95e-04 (9.93e-05 - 2.72e-04) | 9.01e-01 (5.63e-01 - 1.20e+00) | 4.74e-01 (3.60e-01 - 5.46e-01) |
| Symptomatic Men | 25 | 0.25 | Bridging Metric (product) | 3.75e-03 (3.24e-03 - 4.32e-03) | 1.90e-03 (1.90e-03 - 1.90e-03) | 1.86e-03 (1.53e-03 - 2.22e-03) | 9.74e-01 (8.11e-01 - 1.25e+00) | 4.93e-01 (4.48e-01 - 5.53e-01) |
| Symptomatic Men | 25 | 0.5 | Bridging Metric (product) | 5.33e-03 (3.93e-03 - 6.92e-03) | 2.74e-03 (2.74e-03 - 2.74e-03) | 2.47e-03 (2.05e-03 - 2.88e-03) | 1.05e+00 (8.50e-01 - 1.15e+00) | 5.11e-01 (4.60e-01 - 5.35e-01) |
| Symptomatic Men | 25 | 0.75 | Bridging Metric (product) | 4.02e-03 (3.62e-03 - 5.61e-03) | 1.82e-03 (1.82e-03 - 1.82e-03) | 2.25e-03 (2.08e-03 - 2.83e-03) | 1.23e+00 (9.72e-01 - 1.31e+00) | 5.51e-01 (4.93e-01 - 5.66e-01) |
| Symptomatic Men | 25 | 0.95 | Bridging Metric (product) | 6.10e-04 (3.85e-04 - 7.78e-04) | 2.39e-04 (2.39e-04 - 2.39e-04) | 3.53e-04 (2.64e-04 - 4.45e-04) | 1.57e+00 (1.41e+00 - 2.12e+00) | 6.10e-01 (5.85e-01 - 6.80e-01) |
| Symptomatic Men | 10 | 0.05 | Bridging Metric (product) | 7.83e-04 (5.44e-04 - 8.81e-04) | 2.16e-04 (2.16e-04 - 2.16e-04) | 4.70e-04 (3.63e-04 - 6.72e-04) | 2.21e+00 (1.58e+00 - 3.13e+00) | 6.89e-01 (6.10e-01 - 7.58e-01) |
| Symptomatic Men | 10 | 0.25 | Bridging Metric (product) | 6.49e-03 (5.72e-03 - 7.14e-03) | 1.90e-03 (1.90e-03 - 1.90e-03) | 4.67e-03 (3.95e-03 - 5.22e-03) | 2.65e+00 (2.39e+00 - 2.82e+00) | 7.26e-01 (7.05e-01 - 7.38e-01) |
| Symptomatic Men | 10 | 0.5 | Bridging Metric (product) | 9.50e-03 (7.39e-03 - 1.18e-02) | 2.74e-03 (2.74e-03 - 2.74e-03) | 6.63e-03 (5.59e-03 - 8.01e-03) | 2.42e+00 (2.29e+00 - 2.81e+00) | 7.08e-01 (6.96e-01 - 7.38e-01) |
| Symptomatic Men | 10 | 0.75 | Bridging Metric (product) | 7.11e-03 (6.39e-03 - 9.56e-03) | 1.82e-03 (1.82e-03 - 1.82e-03) | 5.42e-03 (4.68e-03 - 6.73e-03) | 2.70e+00 (2.39e+00 - 3.18e+00) | 7.30e-01 (7.05e-01 - 7.61e-01) |
| Symptomatic Men | 10 | 0.95 | Bridging Metric (product) | 9.45e-04 (6.42e-04 - 1.42e-03) | 2.39e-04 (2.39e-04 - 2.39e-04) | 7.06e-04 (5.15e-04 - 1.08e-03) | 3.36e+00 (2.85e+00 - 4.13e+00) | 7.70e-01 (7.40e-01 - 8.05e-01) |
| Symptomatic Men | 5 | 0.05 | Bridging Metric (product) | 1.07e-03 (8.09e-04 - 1.29e-03) | 2.16e-04 (2.16e-04 - 2.16e-04) | 7.66e-04 (6.17e-04 - 1.10e-03) | 3.82e+00 (2.42e+00 - 5.75e+00) | 7.85e-01 (7.07e-01 - 8.51e-01) |
| Symptomatic Men | 5 | 0.25 | Bridging Metric (product) | 1.02e-02 (8.72e-03 - 1.10e-02) | 1.90e-03 (1.90e-03 - 1.90e-03) | 8.39e-03 (6.92e-03 - 8.91e-03) | 4.42e+00 (4.17e+00 - 4.90e+00) | 8.15e-01 (8.07e-01 - 8.31e-01) |
| Symptomatic Men | 5 | 0.5 | Bridging Metric (product) | 1.41e-02 (1.02e-02 - 1.58e-02) | 2.74e-03 (2.74e-03 - 2.74e-03) | 1.09e-02 (8.18e-03 - 1.25e-02) | 3.92e+00 (3.74e+00 - 4.28e+00) | 7.97e-01 (7.89e-01 - 8.10e-01) |
| Symptomatic Men | 5 | 0.75 | Bridging Metric (product) | 1.05e-02 (9.41e-03 - 1.30e-02) | 1.82e-03 (1.82e-03 - 1.82e-03) | 8.72e-03 (7.78e-03 - 1.04e-02) | 4.45e+00 (3.99e+00 - 4.98e+00) | 8.16e-01 (8.00e-01 - 8.33e-01) |
| Symptomatic Men | 5 | 0.95 | Bridging Metric (product) | 1.48e-03 (1.12e-03 - 2.21e-03) | 2.39e-04 (2.39e-04 - 2.39e-04) | 1.24e-03 (9.17e-04 - 1.94e-03) | 6.57e+00 (4.21e+00 - 8.17e+00) | 8.68e-01 (8.07e-01 - 8.91e-01) |
| Symptomatic Men | 4 | 0.05 | Bridging Metric (product) | 1.23e-03 (9.37e-04 - 1.41e-03) | 2.16e-04 (2.16e-04 - 2.16e-04) | 9.30e-04 (6.60e-04 - 1.24e-03) | 4.86e+00 (2.52e+00 - 6.53e+00) | 8.28e-01 (7.15e-01 - 8.67e-01) |
| Symptomatic Men | 4 | 0.25 | Bridging Metric (product) | 1.20e-02 (9.95e-03 - 1.34e-02) | 1.90e-03 (1.90e-03 - 1.90e-03) | 9.79e-03 (8.38e-03 - 1.09e-02) | 5.31e+00 (4.21e+00 - 5.44e+00) | 8.42e-01 (8.08e-01 - 8.45e-01) |
| Symptomatic Men | 4 | 0.5 | Bridging Metric (product) | 1.47e-02 (1.17e-02 - 1.85e-02) | 2.74e-03 (2.74e-03 - 2.74e-03) | 1.20e-02 (9.33e-03 - 1.51e-02) | 4.31e+00 (3.78e+00 - 4.75e+00) | 8.12e-01 (7.91e-01 - 8.26e-01) |
| Symptomatic Men | 4 | 0.75 | Bridging Metric (product) | 1.18e-02 (9.88e-03 - 1.29e-02) | 1.82e-03 (1.82e-03 - 1.82e-03) | 9.90e-03 (8.22e-03 - 1.03e-02) | 4.80e+00 (4.09e+00 - 5.89e+00) | 8.27e-01 (8.04e-01 - 8.55e-01) |
| Symptomatic Men | 4 | 0.95 | Bridging Metric (product) | 2.00e-03 (1.44e-03 - 2.65e-03) | 2.39e-04 (2.39e-04 - 2.39e-04) | 1.74e-03 (1.29e-03 - 2.38e-03) | 7.70e+00 (6.22e+00 - 1.14e+01) | 8.85e-01 (8.61e-01 - 9.19e-01) |
| Symptomatic Men | 3 | 0.05 | Bridging Metric (product) | 1.42e-03 (1.15e-03 - 1.70e-03) | 2.16e-04 (2.16e-04 - 2.16e-04) | 1.15e-03 (9.53e-04 - 1.50e-03) | 7.01e+00 (3.38e+00 - 7.95e+00) | 8.75e-01 (7.69e-01 - 8.88e-01) |
| Symptomatic Men | 3 | 0.25 | Bridging Metric (product) | 1.46e-02 (1.19e-02 - 1.64e-02) | 1.90e-03 (1.90e-03 - 1.90e-03) | 1.28e-02 (1.02e-02 - 1.40e-02) | 6.35e+00 (5.39e+00 - 7.37e+00) | 8.64e-01 (8.43e-01 - 8.80e-01) |
| Symptomatic Men | 3 | 0.5 | Bridging Metric (product) | 1.66e-02 (1.30e-02 - 2.19e-02) | 2.74e-03 (2.74e-03 - 2.74e-03) | 1.39e-02 (1.11e-02 - 1.81e-02) | 5.47e+00 (5.02e+00 - 5.91e+00) | 8.45e-01 (8.34e-01 - 8.55e-01) |
| Symptomatic Men | 3 | 0.75 | Bridging Metric (product) | 1.31e-02 (1.14e-02 - 1.49e-02) | 1.82e-03 (1.82e-03 - 1.82e-03) | 1.07e-02 (9.81e-03 - 1.21e-02) | 6.08e+00 (4.78e+00 - 6.78e+00) | 8.59e-01 (8.27e-01 - 8.71e-01) |
| Symptomatic Men | 3 | 0.95 | Bridging Metric (product) | 2.28e-03 (1.58e-03 - 3.06e-03) | 2.39e-04 (2.39e-04 - 2.39e-04) | 2.05e-03 (1.36e-03 - 2.73e-03) | 8.53e+00 (6.80e+00 - 1.30e+01) | 8.95e-01 (8.72e-01 - 9.28e-01) |
| Symptomatic Men | 2 | 0.05 | Bridging Metric (product) | 1.47e-03 (1.21e-03 - 2.37e-03) | 2.16e-04 (2.16e-04 - 2.16e-04) | 1.18e-03 (9.68e-04 - 2.15e-03) | 6.13e+00 (4.17e+00 - 9.56e+00) | 8.52e-01 (8.06e-01 - 9.05e-01) |
| Symptomatic Men | 2 | 0.25 | Bridging Metric (product) | 1.54e-02 (1.35e-02 - 1.80e-02) | 1.90e-03 (1.90e-03 - 1.90e-03) | 1.33e-02 (1.18e-02 - 1.59e-02) | 7.64e+00 (6.69e+00 - 8.30e+00) | 8.84e-01 (8.69e-01 - 8.92e-01) |
| Symptomatic Men | 2 | 0.5 | Bridging Metric (product) | 2.11e-02 (1.72e-02 - 2.38e-02) | 2.74e-03 (2.74e-03 - 2.74e-03) | 1.87e-02 (1.47e-02 - 1.99e-02) | 6.83e+00 (5.68e+00 - 7.21e+00) | 8.72e-01 (8.50e-01 - 8.78e-01) |
| Symptomatic Men | 2 | 0.75 | Bridging Metric (product) | 1.75e-02 (1.33e-02 - 1.78e-02) | 1.82e-03 (1.82e-03 - 1.82e-03) | 1.45e-02 (1.13e-02 - 1.59e-02) | 7.42e+00 (5.17e+00 - 9.39e+00) | 8.79e-01 (8.36e-01 - 9.04e-01) |
| Symptomatic Men | 2 | 0.95 | Bridging Metric (product) | 2.81e-03 (2.07e-03 - 3.90e-03) | 2.39e-04 (2.39e-04 - 2.39e-04) | 2.63e-03 (1.86e-03 - 3.56e-03) | 1.15e+01 (9.57e+00 - 1.51e+01) | 9.20e-01 (9.05e-01 - 9.38e-01) |
| Symptomatic Men | 1 | 0.05 | Bridging Metric (product) | 3.33e-03 (2.35e-03 - 4.19e-03) | 2.16e-04 (2.16e-04 - 2.16e-04) | 3.07e-03 (2.09e-03 - 4.01e-03) | 1.42e+01 (9.18e+00 - 2.05e+01) | 9.34e-01 (9.02e-01 - 9.53e-01) |
| Symptomatic Men | 1 | 0.25 | Bridging Metric (product) | 2.09e-02 (1.57e-02 - 2.27e-02) | 1.90e-03 (1.90e-03 - 1.90e-03) | 1.86e-02 (1.43e-02 - 2.05e-02) | 9.43e+00 (8.15e+00 - 1.06e+01) | 9.04e-01 (8.90e-01 - 9.14e-01) |
| Symptomatic Men | 1 | 0.5 | Bridging Metric (product) | 2.52e-02 (1.92e-02 - 2.61e-02) | 2.74e-03 (2.74e-03 - 2.74e-03) | 2.15e-02 (1.78e-02 - 2.31e-02) | 8.32e+00 (6.75e+00 - 9.31e+00) | 8.93e-01 (8.71e-01 - 9.03e-01) |
| Symptomatic Men | 1 | 0.75 | Bridging Metric (product) | 2.21e-02 (1.93e-02 - 2.47e-02) | 1.82e-03 (1.82e-03 - 1.82e-03) | 1.89e-02 (1.80e-02 - 2.26e-02) | 1.10e+01 (7.90e+00 - 1.25e+01) | 9.17e-01 (8.86e-01 - 9.26e-01) |
| Symptomatic Men | 1 | 0.95 | Bridging Metric (product) | 4.42e-03 (3.52e-03 - 6.96e-03) | 2.39e-04 (2.39e-04 - 2.39e-04) | 4.20e-03 (3.40e-03 - 6.64e-03) | 2.07e+01 (1.49e+01 - 2.51e+01) | 9.54e-01 (9.35e-01 - 9.62e-01) |
| High Activity Symptomatic Men | 100 | 0.05 | Bridging Metric (product) | 4.87e-04 (3.95e-04 - 5.26e-04) | 2.16e-04 (2.16e-04 - 2.16e-04) | 2.63e-04 (1.95e-04 - 3.02e-04) | 1.27e+00 (8.69e-01 - 1.47e+00) | 5.57e-01 (4.62e-01 - 5.96e-01) |
| High Activity Symptomatic Men | 100 | 0.25 | Bridging Metric (product) | 3.81e-03 (3.23e-03 - 4.47e-03) | 1.90e-03 (1.90e-03 - 1.90e-03) | 1.91e-03 (1.77e-03 - 2.38e-03) | 1.20e+00 (8.57e-01 - 1.42e+00) | 5.45e-01 (4.61e-01 - 5.87e-01) |
| High Activity Symptomatic Men | 100 | 0.5 | Bridging Metric (product) | 5.53e-03 (4.47e-03 - 6.69e-03) | 2.74e-03 (2.74e-03 - 2.74e-03) | 2.62e-03 (2.33e-03 - 3.56e-03) | 1.37e+00 (8.94e-01 - 1.42e+00) | 5.77e-01 (4.67e-01 - 5.88e-01) |
| High Activity Symptomatic Men | 100 | 0.75 | Bridging Metric (product) | 4.78e-03 (4.07e-03 - 6.06e-03) | 1.82e-03 (1.82e-03 - 1.82e-03) | 3.06e-03 (2.28e-03 - 3.29e-03) | 1.41e+00 (1.11e+00 - 1.65e+00) | 5.86e-01 (5.26e-01 - 6.22e-01) |
| High Activity Symptomatic Men | 100 | 0.95 | Bridging Metric (product) | 9.47e-04 (6.16e-04 - 1.09e-03) | 2.39e-04 (2.39e-04 - 2.39e-04) | 6.91e-04 (4.36e-04 - 7.79e-04) | 2.97e+00 (2.37e+00 - 3.58e+00) | 7.48e-01 (7.03e-01 - 7.82e-01) |
| High Activity Symptomatic Men | 50 | 0.05 | Bridging Metric (product) | 6.79e-04 (5.55e-04 - 7.56e-04) | 2.16e-04 (2.16e-04 - 2.16e-04) | 3.97e-04 (2.87e-04 - 5.55e-04) | 2.04e+00 (1.28e+00 - 2.33e+00) | 6.71e-01 (5.56e-01 - 7.00e-01) |
| High Activity Symptomatic Men | 50 | 0.25 | Bridging Metric (product) | 5.15e-03 (4.53e-03 - 6.38e-03) | 1.90e-03 (1.90e-03 - 1.90e-03) | 3.34e-03 (3.27e-03 - 4.05e-03) | 2.01e+00 (1.63e+00 - 2.28e+00) | 6.67e-01 (6.20e-01 - 6.95e-01) |
| High Activity Symptomatic Men | 50 | 0.5 | Bridging Metric (product) | 8.24e-03 (6.42e-03 - 9.66e-03) | 2.74e-03 (2.74e-03 - 2.74e-03) | 5.12e-03 (4.22e-03 - 6.28e-03) | 2.30e+00 (1.59e+00 - 2.44e+00) | 6.97e-01 (6.06e-01 - 7.09e-01) |
| High Activity Symptomatic Men | 50 | 0.75 | Bridging Metric (product) | 7.18e-03 (5.62e-03 - 8.24e-03) | 1.82e-03 (1.82e-03 - 1.82e-03) | 5.35e-03 (3.77e-03 - 5.79e-03) | 2.32e+00 (1.99e+00 - 3.07e+00) | 6.98e-01 (6.66e-01 - 7.54e-01) |
| High Activity Symptomatic Men | 50 | 0.95 | Bridging Metric (product) | 1.32e-03 (8.12e-04 - 1.67e-03) | 2.39e-04 (2.39e-04 - 2.39e-04) | 1.02e-03 (6.85e-04 - 1.35e-03) | 4.54e+00 (4.18e+00 - 5.08e+00) | 8.19e-01 (8.07e-01 - 8.35e-01) |
| High Activity Symptomatic Men | 25 | 0.05 | Bridging Metric (product) | 1.03e-03 (8.59e-04 - 1.12e-03) | 2.16e-04 (2.16e-04 - 2.16e-04) | 7.91e-04 (5.51e-04 - 9.24e-04) | 3.62e+00 (2.68e+00 - 3.88e+00) | 7.84e-01 (7.29e-01 - 7.95e-01) |
| High Activity Symptomatic Men | 25 | 0.25 | Bridging Metric (product) | 8.27e-03 (6.66e-03 - 9.43e-03) | 1.90e-03 (1.90e-03 - 1.90e-03) | 6.53e-03 (5.27e-03 - 7.13e-03) | 3.57e+00 (3.08e+00 - 4.14e+00) | 7.81e-01 (7.55e-01 - 8.05e-01) |
| High Activity Symptomatic Men | 25 | 0.5 | Bridging Metric (product) | 1.19e-02 (9.56e-03 - 1.38e-02) | 2.74e-03 (2.74e-03 - 2.74e-03) | 8.91e-03 (7.46e-03 - 1.04e-02) | 3.70e+00 (2.73e+00 - 3.92e+00) | 7.87e-01 (7.28e-01 - 7.97e-01) |
| High Activity Symptomatic Men | 25 | 0.75 | Bridging Metric (product) | 1.07e-02 (8.90e-03 - 1.20e-02) | 1.82e-03 (1.82e-03 - 1.82e-03) | 8.78e-03 (7.29e-03 - 9.61e-03) | 4.38e+00 (3.59e+00 - 5.27e+00) | 8.13e-01 (7.82e-01 - 8.41e-01) |
| High Activity Symptomatic Men | 25 | 0.95 | Bridging Metric (product) | 2.07e-03 (1.25e-03 - 2.49e-03) | 2.39e-04 (2.39e-04 - 2.39e-04) | 1.79e-03 (1.13e-03 - 2.17e-03) | 7.23e+00 (6.33e+00 - 8.15e+00) | 8.78e-01 (8.64e-01 - 8.91e-01) |
| High Activity Symptomatic Men | 10 | 0.05 | Bridging Metric (product) | 1.62e-03 (1.33e-03 - 2.00e-03) | 2.16e-04 (2.16e-04 - 2.16e-04) | 1.41e-03 (1.02e-03 - 1.84e-03) | 6.57e+00 (5.07e+00 - 9.48e+00) | 8.68e-01 (8.33e-01 - 9.04e-01) |
| High Activity Symptomatic Men | 10 | 0.25 | Bridging Metric (product) | 1.53e-02 (1.37e-02 - 1.63e-02) | 1.90e-03 (1.90e-03 - 1.90e-03) | 1.33e-02 (1.18e-02 - 1.45e-02) | 7.41e+00 (5.97e+00 - 8.00e+00) | 8.81e-01 (8.56e-01 - 8.89e-01) |
| High Activity Symptomatic Men | 10 | 0.5 | Bridging Metric (product) | 1.91e-02 (1.84e-02 - 2.42e-02) | 2.74e-03 (2.74e-03 - 2.74e-03) | 1.68e-02 (1.57e-02 - 2.03e-02) | 6.70e+00 (5.21e+00 - 7.60e+00) | 8.70e-01 (8.38e-01 - 8.84e-01) |
| High Activity Symptomatic Men | 10 | 0.75 | Bridging Metric (product) | 1.83e-02 (1.56e-02 - 1.98e-02) | 1.82e-03 (1.82e-03 - 1.82e-03) | 1.64e-02 (1.39e-02 - 1.70e-02) | 8.04e+00 (5.78e+00 - 9.34e+00) | 8.89e-01 (8.52e-01 - 9.03e-01) |
| High Activity Symptomatic Men | 10 | 0.95 | Bridging Metric (product) | 3.16e-03 (2.27e-03 - 4.65e-03) | 2.39e-04 (2.39e-04 - 2.39e-04) | 2.88e-03 (2.11e-03 - 4.35e-03) | 1.36e+01 (1.15e+01 - 1.84e+01) | 9.31e-01 (9.20e-01 - 9.49e-01) |
| High Activity Symptomatic Men | 5 | 0.05 | Bridging Metric (product) | 2.50e-03 (2.29e-03 - 2.86e-03) | 2.16e-04 (2.16e-04 - 2.16e-04) | 2.28e-03 (2.07e-03 - 2.63e-03) | 9.75e+00 (9.28e+00 - 1.34e+01) | 9.07e-01 (9.03e-01 - 9.29e-01) |
| High Activity Symptomatic Men | 5 | 0.25 | Bridging Metric (product) | 1.82e-02 (1.77e-02 - 2.49e-02) | 1.90e-03 (1.90e-03 - 1.90e-03) | 1.66e-02 (1.55e-02 - 2.24e-02) | 9.76e+00 (8.67e+00 - 1.06e+01) | 9.07e-01 (8.97e-01 - 9.14e-01) |
| High Activity Symptomatic Men | 5 | 0.5 | Bridging Metric (product) | 2.92e-02 (2.20e-02 - 3.30e-02) | 2.74e-03 (2.74e-03 - 2.74e-03) | 2.55e-02 (2.01e-02 - 2.98e-02) | 1.03e+01 (8.23e+00 - 1.06e+01) | 9.12e-01 (8.91e-01 - 9.14e-01) |
| High Activity Symptomatic Men | 5 | 0.75 | Bridging Metric (product) | 2.40e-02 (2.09e-02 - 2.65e-02) | 1.82e-03 (1.82e-03 - 1.82e-03) | 2.23e-02 (1.90e-02 - 2.39e-02) | 1.04e+01 (8.92e+00 - 1.40e+01) | 9.12e-01 (8.99e-01 - 9.33e-01) |
| High Activity Symptomatic Men | 5 | 0.95 | Bridging Metric (product) | 4.75e-03 (3.39e-03 - 7.98e-03) | 2.39e-04 (2.39e-04 - 2.39e-04) | 4.46e-03 (3.22e-03 - 7.67e-03) | 2.39e+01 (1.69e+01 - 2.90e+01) | 9.60e-01 (9.44e-01 - 9.67e-01) |
| High Activity Symptomatic Men | 4 | 0.05 | Bridging Metric (product) | 2.63e-03 (2.17e-03 - 3.17e-03) | 2.16e-04 (2.16e-04 - 2.16e-04) | 2.43e-03 (2.01e-03 - 2.98e-03) | 1.23e+01 (8.16e+00 - 1.61e+01) | 9.23e-01 (8.90e-01 - 9.42e-01) |
| High Activity Symptomatic Men | 4 | 0.25 | Bridging Metric (product) | 2.30e-02 (1.98e-02 - 2.62e-02) | 1.90e-03 (1.90e-03 - 1.90e-03) | 2.11e-02 (1.80e-02 - 2.38e-02) | 1.22e+01 (9.48e+00 - 1.38e+01) | 9.24e-01 (9.05e-01 - 9.32e-01) |
| High Activity Symptomatic Men | 4 | 0.5 | Bridging Metric (product) | 3.19e-02 (2.33e-02 - 3.43e-02) | 2.74e-03 (2.74e-03 - 2.74e-03) | 2.90e-02 (2.15e-02 - 3.08e-02) | 1.04e+01 (9.13e+00 - 1.15e+01) | 9.12e-01 (9.01e-01 - 9.20e-01) |
| High Activity Symptomatic Men | 4 | 0.75 | Bridging Metric (product) | 2.73e-02 (2.47e-02 - 2.94e-02) | 1.82e-03 (1.82e-03 - 1.82e-03) | 2.54e-02 (2.29e-02 - 2.64e-02) | 1.31e+01 (9.30e+00 - 1.60e+01) | 9.29e-01 (9.03e-01 - 9.41e-01) |
| High Activity Symptomatic Men | 4 | 0.95 | Bridging Metric (product) | 6.20e-03 (4.67e-03 - 8.95e-03) | 2.39e-04 (2.39e-04 - 2.39e-04) | 5.92e-03 (4.51e-03 - 8.63e-03) | 2.48e+01 (2.04e+01 - 2.93e+01) | 9.61e-01 (9.53e-01 - 9.67e-01) |
| High Activity Symptomatic Men | 3 | 0.05 | Bridging Metric (product) | 3.00e-03 (2.72e-03 - 4.01e-03) | 2.16e-04 (2.16e-04 - 2.16e-04) | 2.80e-03 (2.55e-03 - 3.72e-03) | 1.41e+01 (1.28e+01 - 1.86e+01) | 9.33e-01 (9.28e-01 - 9.49e-01) |
| High Activity Symptomatic Men | 3 | 0.25 | Bridging Metric (product) | 2.50e-02 (2.22e-02 - 2.84e-02) | 1.90e-03 (1.90e-03 - 1.90e-03) | 2.29e-02 (2.07e-02 - 2.65e-02) | 1.25e+01 (1.11e+01 - 1.43e+01) | 9.26e-01 (9.17e-01 - 9.35e-01) |
| High Activity Symptomatic Men | 3 | 0.5 | Bridging Metric (product) | 3.48e-02 (2.84e-02 - 3.71e-02) | 2.74e-03 (2.74e-03 - 2.74e-03) | 3.16e-02 (2.66e-02 - 3.39e-02) | 1.14e+01 (9.91e+00 - 1.31e+01) | 9.19e-01 (9.08e-01 - 9.29e-01) |
| High Activity Symptomatic Men | 3 | 0.75 | Bridging Metric (product) | 3.00e-02 (2.54e-02 - 3.38e-02) | 1.82e-03 (1.82e-03 - 1.82e-03) | 2.83e-02 (2.37e-02 - 3.08e-02) | 1.43e+01 (1.05e+01 - 1.60e+01) | 9.34e-01 (9.13e-01 - 9.41e-01) |
| High Activity Symptomatic Men | 3 | 0.95 | Bridging Metric (product) | 6.19e-03 (4.98e-03 - 9.40e-03) | 2.39e-04 (2.39e-04 - 2.39e-04) | 5.91e-03 (4.82e-03 - 9.06e-03) | 2.74e+01 (2.23e+01 - 3.40e+01) | 9.65e-01 (9.57e-01 - 9.71e-01) |
| High Activity Symptomatic Men | 2 | 0.05 | Bridging Metric (product) | 3.78e-03 (3.48e-03 - 5.10e-03) | 2.16e-04 (2.16e-04 - 2.16e-04) | 3.56e-03 (3.23e-03 - 4.82e-03) | 1.79e+01 (1.24e+01 - 2.00e+01) | 9.47e-01 (9.24e-01 - 9.52e-01) |
| High Activity Symptomatic Men | 2 | 0.25 | Bridging Metric (product) | 3.52e-02 (2.88e-02 - 3.67e-02) | 1.90e-03 (1.90e-03 - 1.90e-03) | 3.25e-02 (2.69e-02 - 3.46e-02) | 1.35e+01 (1.27e+01 - 1.84e+01) | 9.31e-01 (9.27e-01 - 9.48e-01) |
| High Activity Symptomatic Men | 2 | 0.5 | Bridging Metric (product) | 3.92e-02 (3.41e-02 - 4.19e-02) | 2.74e-03 (2.74e-03 - 2.74e-03) | 3.64e-02 (3.25e-02 - 3.81e-02) | 1.39e+01 (1.11e+01 - 1.59e+01) | 9.33e-01 (9.17e-01 - 9.41e-01) |
| High Activity Symptomatic Men | 2 | 0.75 | Bridging Metric (product) | 3.59e-02 (2.77e-02 - 4.16e-02) | 1.82e-03 (1.82e-03 - 1.82e-03) | 3.34e-02 (2.64e-02 - 3.94e-02) | 1.60e+01 (1.34e+01 - 2.09e+01) | 9.41e-01 (9.30e-01 - 9.54e-01) |
| High Activity Symptomatic Men | 2 | 0.95 | Bridging Metric (product) | 9.17e-03 (5.89e-03 - 1.29e-02) | 2.39e-04 (2.39e-04 - 2.39e-04) | 8.99e-03 (5.67e-03 - 1.25e-02) | 3.96e+01 (2.72e+01 - 4.82e+01) | 9.75e-01 (9.64e-01 - 9.80e-01) |
| High Activity Symptomatic Men | 1 | 0.05 | Bridging Metric (product) | 4.46e-03 (3.18e-03 - 8.01e-03) | 2.16e-04 (2.16e-04 - 2.16e-04) | 4.25e-03 (2.89e-03 - 7.80e-03) | 1.96e+01 (1.26e+01 - 3.76e+01) | 9.51e-01 (9.24e-01 - 9.74e-01) |
| High Activity Symptomatic Men | 1 | 0.25 | Bridging Metric (product) | 3.85e-02 (2.92e-02 - 4.00e-02) | 1.90e-03 (1.90e-03 - 1.90e-03) | 3.62e-02 (2.74e-02 - 3.81e-02) | 1.82e+01 (1.42e+01 - 1.91e+01) | 9.48e-01 (9.34e-01 - 9.50e-01) |
| High Activity Symptomatic Men | 1 | 0.5 | Bridging Metric (product) | 4.69e-02 (4.10e-02 - 5.08e-02) | 2.74e-03 (2.74e-03 - 2.74e-03) | 4.37e-02 (3.86e-02 - 4.82e-02) | 1.73e+01 (1.16e+01 - 2.19e+01) | 9.44e-01 (9.20e-01 - 9.56e-01) |
| High Activity Symptomatic Men | 1 | 0.75 | Bridging Metric (product) | 3.97e-02 (3.74e-02 - 4.36e-02) | 1.82e-03 (1.82e-03 - 1.82e-03) | 3.80e-02 (3.46e-02 - 4.06e-02) | 2.20e+01 (1.48e+01 - 2.37e+01) | 9.56e-01 (9.36e-01 - 9.59e-01) |
| High Activity Symptomatic Men | 1 | 0.95 | Bridging Metric (product) | 9.86e-03 (8.58e-03 - 1.77e-02) | 2.39e-04 (2.39e-04 - 2.39e-04) | 9.71e-03 (8.34e-03 - 1.73e-02) | 4.82e+01 (3.99e+01 - 6.76e+01) | 9.79e-01 (9.76e-01 - 9.85e-01) |
| Random Subsample High Activity | 100 | 0.05 | Bridging Metric (product) | 6.91e-04 (5.25e-04 - 1.04e-03) | 2.16e-04 (2.16e-04 - 2.16e-04) | 4.54e-04 (3.07e-04 - 8.52e-04) | 2.46e+00 (1.47e+00 - 3.80e+00) | 7.11e-01 (5.81e-01 - 7.90e-01) |
| Random Subsample High Activity | 100 | 0.25 | Bridging Metric (product) | 5.43e-03 (4.58e-03 - 5.74e-03) | 1.90e-03 (1.90e-03 - 1.90e-03) | 3.73e-03 (2.86e-03 - 3.97e-03) | 1.81e+00 (1.49e+00 - 2.06e+00) | 6.44e-01 (5.98e-01 - 6.73e-01) |
| Random Subsample High Activity | 100 | 0.5 | Bridging Metric (product) | 8.28e-03 (7.21e-03 - 8.71e-03) | 2.74e-03 (2.74e-03 - 2.74e-03) | 5.28e-03 (4.28e-03 - 5.71e-03) | 2.09e+00 (1.13e+00 - 2.96e+00) | 6.76e-01 (5.26e-01 - 7.46e-01) |
| Random Subsample High Activity | 100 | 0.75 | Bridging Metric (product) | 7.33e-03 (6.70e-03 - 7.58e-03) | 1.82e-03 (1.82e-03 - 1.82e-03) | 4.65e-03 (4.02e-03 - 5.65e-03) | 2.31e+00 (1.62e+00 - 3.12e+00) | 6.97e-01 (6.17e-01 - 7.57e-01) |
| Random Subsample High Activity | 100 | 0.95 | Bridging Metric (product) | 1.78e-03 (1.05e-03 - 2.22e-03) | 2.39e-04 (2.39e-04 - 2.39e-04) | 1.50e-03 (9.11e-04 - 1.91e-03) | 6.39e+00 (5.02e+00 - 6.72e+00) | 8.65e-01 (8.33e-01 - 8.70e-01) |
| Random Subsample High Activity | 50 | 0.05 | Bridging Metric (product) | 1.10e-03 (1.00e-03 - 1.50e-03) | 2.16e-04 (2.16e-04 - 2.16e-04) | 8.85e-04 (8.45e-04 - 1.31e-03) | 5.01e+00 (3.62e+00 - 6.73e+00) | 8.33e-01 (7.83e-01 - 8.69e-01) |
| Random Subsample High Activity | 50 | 0.25 | Bridging Metric (product) | 9.14e-03 (8.27e-03 - 9.67e-03) | 1.90e-03 (1.90e-03 - 1.90e-03) | 6.86e-03 (5.63e-03 - 7.83e-03) | 3.70e+00 (2.68e+00 - 5.28e+00) | 7.86e-01 (7.26e-01 - 8.39e-01) |
| Random Subsample High Activity | 50 | 0.5 | Bridging Metric (product) | 1.34e-02 (1.21e-02 - 1.38e-02) | 2.74e-03 (2.74e-03 - 2.74e-03) | 1.05e-02 (9.65e-03 - 1.14e-02) | 3.90e+00 (2.66e+00 - 6.03e+00) | 7.95e-01 (7.23e-01 - 8.56e-01) |
| Random Subsample High Activity | 50 | 0.75 | Bridging Metric (product) | 1.23e-02 (1.06e-02 - 1.29e-02) | 1.82e-03 (1.82e-03 - 1.82e-03) | 9.31e-03 (8.68e-03 - 1.05e-02) | 4.44e+00 (3.55e+00 - 5.64e+00) | 8.16e-01 (7.79e-01 - 8.49e-01) |
| Random Subsample High Activity | 50 | 0.95 | Bridging Metric (product) | 3.08e-03 (1.50e-03 - 3.80e-03) | 2.39e-04 (2.39e-04 - 2.39e-04) | 2.80e-03 (1.40e-03 - 3.53e-03) | 1.09e+01 (9.13e+00 - 1.36e+01) | 9.16e-01 (9.01e-01 - 9.32e-01) |
| Random Subsample High Activity | 25 | 0.05 | Bridging Metric (product) | 1.86e-03 (1.59e-03 - 2.69e-03) | 2.16e-04 (2.16e-04 - 2.16e-04) | 1.64e-03 (1.31e-03 - 2.45e-03) | 7.99e+00 (5.95e+00 - 1.14e+01) | 8.89e-01 (8.54e-01 - 9.18e-01) |
| Random Subsample High Activity | 25 | 0.25 | Bridging Metric (product) | 1.50e-02 (1.34e-02 - 1.55e-02) | 1.90e-03 (1.90e-03 - 1.90e-03) | 1.24e-02 (1.16e-02 - 1.41e-02) | 6.82e+00 (4.90e+00 - 8.70e+00) | 8.71e-01 (8.30e-01 - 8.97e-01) |
| Random Subsample High Activity | 25 | 0.5 | Bridging Metric (product) | 2.10e-02 (2.03e-02 - 2.22e-02) | 2.74e-03 (2.74e-03 - 2.74e-03) | 1.85e-02 (1.79e-02 - 1.90e-02) | 6.69e+00 (4.80e+00 - 1.01e+01) | 8.70e-01 (8.26e-01 - 9.10e-01) |
| Random Subsample High Activity | 25 | 0.75 | Bridging Metric (product) | 1.86e-02 (1.57e-02 - 1.96e-02) | 1.82e-03 (1.82e-03 - 1.82e-03) | 1.62e-02 (1.38e-02 - 1.74e-02) | 7.28e+00 (5.98e+00 - 9.04e+00) | 8.79e-01 (8.57e-01 - 9.00e-01) |
| Random Subsample High Activity | 25 | 0.95 | Bridging Metric (product) | 4.40e-03 (3.12e-03 - 6.16e-03) | 2.39e-04 (2.39e-04 - 2.39e-04) | 4.11e-03 (2.99e-03 - 5.83e-03) | 1.85e+01 (1.60e+01 - 2.38e+01) | 9.49e-01 (9.41e-01 - 9.60e-01) |
| Random Subsample High Activity | 10 | 0.05 | Bridging Metric (product) | 3.62e-03 (2.98e-03 - 4.43e-03) | 2.16e-04 (2.16e-04 - 2.16e-04) | 3.35e-03 (2.65e-03 - 4.17e-03) | 1.50e+01 (1.24e+01 - 1.84e+01) | 9.37e-01 (9.25e-01 - 9.48e-01) |
| Random Subsample High Activity | 10 | 0.25 | Bridging Metric (product) | 2.48e-02 (2.24e-02 - 2.67e-02) | 1.90e-03 (1.90e-03 - 1.90e-03) | 2.31e-02 (2.05e-02 - 2.41e-02) | 1.09e+01 (9.40e+00 - 1.44e+01) | 9.15e-01 (9.04e-01 - 9.35e-01) |
| Random Subsample High Activity | 10 | 0.5 | Bridging Metric (product) | 3.31e-02 (2.73e-02 - 3.50e-02) | 2.74e-03 (2.74e-03 - 2.74e-03) | 2.94e-02 (2.50e-02 - 3.27e-02) | 1.08e+01 (9.48e+00 - 1.47e+01) | 9.15e-01 (9.05e-01 - 9.36e-01) |
| Random Subsample High Activity | 10 | 0.75 | Bridging Metric (product) | 2.89e-02 (2.56e-02 - 3.19e-02) | 1.82e-03 (1.82e-03 - 1.82e-03) | 2.71e-02 (2.40e-02 - 2.87e-02) | 1.37e+01 (1.06e+01 - 1.48e+01) | 9.32e-01 (9.14e-01 - 9.37e-01) |
| Random Subsample High Activity | 10 | 0.95 | Bridging Metric (product) | 9.27e-03 (4.99e-03 - 1.24e-02) | 2.39e-04 (2.39e-04 - 2.39e-04) | 9.04e-03 (4.80e-03 - 1.21e-02) | 3.41e+01 (2.77e+01 - 3.89e+01) | 9.71e-01 (9.65e-01 - 9.75e-01) |
| Random Subsample High Activity | 5 | 0.05 | Bridging Metric (product) | 5.85e-03 (4.49e-03 - 7.64e-03) | 2.16e-04 (2.16e-04 - 2.16e-04) | 5.57e-03 (4.21e-03 - 7.47e-03) | 2.48e+01 (1.54e+01 - 3.44e+01) | 9.61e-01 (9.39e-01 - 9.72e-01) |
| Random Subsample High Activity | 5 | 0.25 | Bridging Metric (product) | 3.10e-02 (2.82e-02 - 3.23e-02) | 1.90e-03 (1.90e-03 - 1.90e-03) | 2.86e-02 (2.58e-02 - 3.06e-02) | 1.61e+01 (1.28e+01 - 1.71e+01) | 9.41e-01 (9.27e-01 - 9.45e-01) |
| Random Subsample High Activity | 5 | 0.5 | Bridging Metric (product) | 4.21e-02 (3.54e-02 - 4.47e-02) | 2.74e-03 (2.74e-03 - 2.74e-03) | 3.92e-02 (3.37e-02 - 4.05e-02) | 1.46e+01 (1.16e+01 - 1.70e+01) | 9.36e-01 (9.20e-01 - 9.44e-01) |
| Random Subsample High Activity | 5 | 0.75 | Bridging Metric (product) | 4.14e-02 (3.40e-02 - 4.54e-02) | 1.82e-03 (1.82e-03 - 1.82e-03) | 3.90e-02 (3.24e-02 - 4.22e-02) | 1.95e+01 (1.42e+01 - 2.03e+01) | 9.51e-01 (9.34e-01 - 9.53e-01) |
| Random Subsample High Activity | 5 | 0.95 | Bridging Metric (product) | 1.00e-02 (6.81e-03 - 1.64e-02) | 2.39e-04 (2.39e-04 - 2.39e-04) | 9.79e-03 (6.67e-03 - 1.60e-02) | 4.73e+01 (4.06e+01 - 5.63e+01) | 9.79e-01 (9.76e-01 - 9.83e-01) |
| Random Subsample High Activity | 4 | 0.05 | Bridging Metric (product) | 5.09e-03 (4.18e-03 - 1.09e-02) | 2.16e-04 (2.16e-04 - 2.16e-04) | 4.88e-03 (3.88e-03 - 1.07e-02) | 2.34e+01 (1.51e+01 - 5.57e+01) | 9.59e-01 (9.36e-01 - 9.81e-01) |
| Random Subsample High Activity | 4 | 0.25 | Bridging Metric (product) | 3.38e-02 (3.20e-02 - 3.79e-02) | 1.90e-03 (1.90e-03 - 1.90e-03) | 3.20e-02 (3.07e-02 - 3.52e-02) | 1.83e+01 (1.50e+01 - 2.01e+01) | 9.48e-01 (9.37e-01 - 9.53e-01) |
| Random Subsample High Activity | 4 | 0.5 | Bridging Metric (product) | 4.54e-02 (3.63e-02 - 5.28e-02) | 2.74e-03 (2.74e-03 - 2.74e-03) | 4.19e-02 (3.43e-02 - 5.01e-02) | 1.72e+01 (1.44e+01 - 2.07e+01) | 9.45e-01 (9.35e-01 - 9.54e-01) |
| Random Subsample High Activity | 4 | 0.75 | Bridging Metric (product) | 4.21e-02 (3.73e-02 - 4.54e-02) | 1.82e-03 (1.82e-03 - 1.82e-03) | 4.03e-02 (3.52e-02 - 4.30e-02) | 2.06e+01 (1.54e+01 - 2.34e+01) | 9.53e-01 (9.38e-01 - 9.59e-01) |
| Random Subsample High Activity | 4 | 0.95 | Bridging Metric (product) | 1.31e-02 (9.35e-03 - 2.02e-02) | 2.39e-04 (2.39e-04 - 2.39e-04) | 1.29e-02 (9.21e-03 - 1.98e-02) | 5.74e+01 (5.14e+01 - 7.53e+01) | 9.83e-01 (9.81e-01 - 9.87e-01) |
| Random Subsample High Activity | 3 | 0.05 | Bridging Metric (product) | 5.31e-03 (4.48e-03 - 6.28e-03) | 2.16e-04 (2.16e-04 - 2.16e-04) | 5.14e-03 (4.20e-03 - 6.08e-03) | 2.28e+01 (1.70e+01 - 3.80e+01) | 9.58e-01 (9.44e-01 - 9.74e-01) |
| Random Subsample High Activity | 3 | 0.25 | Bridging Metric (product) | 3.71e-02 (3.12e-02 - 4.28e-02) | 1.90e-03 (1.90e-03 - 1.90e-03) | 3.55e-02 (2.89e-02 - 4.07e-02) | 2.00e+01 (1.70e+01 - 2.33e+01) | 9.52e-01 (9.44e-01 - 9.59e-01) |
| Random Subsample High Activity | 3 | 0.5 | Bridging Metric (product) | 5.78e-02 (4.77e-02 - 5.88e-02) | 2.74e-03 (2.74e-03 - 2.74e-03) | 5.37e-02 (4.60e-02 - 5.62e-02) | 1.94e+01 (1.70e+01 - 2.18e+01) | 9.51e-01 (9.44e-01 - 9.56e-01) |
| Random Subsample High Activity | 3 | 0.75 | Bridging Metric (product) | 4.45e-02 (3.57e-02 - 5.10e-02) | 1.82e-03 (1.82e-03 - 1.82e-03) | 4.24e-02 (3.25e-02 - 4.91e-02) | 2.17e+01 (1.47e+01 - 2.87e+01) | 9.54e-01 (9.36e-01 - 9.66e-01) |
| Random Subsample High Activity | 3 | 0.95 | Bridging Metric (product) | 1.47e-02 (1.23e-02 - 2.19e-02) | 2.39e-04 (2.39e-04 - 2.39e-04) | 1.44e-02 (1.21e-02 - 2.16e-02) | 6.38e+01 (6.15e+01 - 8.05e+01) | 9.85e-01 (9.84e-01 - 9.88e-01) |
| Random Subsample High Activity | 2 | 0.05 | Bridging Metric (product) | 7.38e-03 (4.39e-03 - 1.23e-02) | 2.16e-04 (2.16e-04 - 2.16e-04) | 7.16e-03 (4.18e-03 - 1.21e-02) | 3.41e+01 (1.86e+01 - 5.54e+01) | 9.72e-01 (9.43e-01 - 9.82e-01) |
| Random Subsample High Activity | 2 | 0.25 | Bridging Metric (product) | 3.79e-02 (3.54e-02 - 4.26e-02) | 1.90e-03 (1.90e-03 - 1.90e-03) | 3.59e-02 (3.36e-02 - 4.09e-02) | 1.79e+01 (1.69e+01 - 2.23e+01) | 9.47e-01 (9.44e-01 - 9.57e-01) |
| Random Subsample High Activity | 2 | 0.5 | Bridging Metric (product) | 4.97e-02 (4.57e-02 - 5.29e-02) | 2.74e-03 (2.74e-03 - 2.74e-03) | 4.70e-02 (4.36e-02 - 5.04e-02) | 1.95e+01 (1.26e+01 - 2.39e+01) | 9.51e-01 (9.24e-01 - 9.60e-01) |
| Random Subsample High Activity | 2 | 0.75 | Bridging Metric (product) | 4.85e-02 (4.33e-02 - 5.28e-02) | 1.82e-03 (1.82e-03 - 1.82e-03) | 4.61e-02 (4.13e-02 - 5.04e-02) | 2.69e+01 (1.75e+01 - 2.73e+01) | 9.64e-01 (9.45e-01 - 9.65e-01) |
| Random Subsample High Activity | 2 | 0.95 | Bridging Metric (product) | 1.82e-02 (1.13e-02 - 2.36e-02) | 2.39e-04 (2.39e-04 - 2.39e-04) | 1.81e-02 (1.11e-02 - 2.32e-02) | 7.41e+01 (6.03e+01 - 9.42e+01) | 9.87e-01 (9.84e-01 - 9.89e-01) |
| Random Subsample High Activity | 1 | 0.05 | Bridging Metric (product) | 1.12e-02 (6.84e-03 - 1.56e-02) | 2.16e-04 (2.16e-04 - 2.16e-04) | 1.10e-02 (6.50e-03 - 1.54e-02) | 5.64e+01 (2.42e+01 - 7.84e+01) | 9.83e-01 (9.52e-01 - 9.87e-01) |
| Random Subsample High Activity | 1 | 0.25 | Bridging Metric (product) | 4.83e-02 (4.21e-02 - 6.02e-02) | 1.90e-03 (1.90e-03 - 1.90e-03) | 4.62e-02 (4.04e-02 - 5.79e-02) | 2.53e+01 (1.93e+01 - 3.60e+01) | 9.61e-01 (9.51e-01 - 9.73e-01) |
| Random Subsample High Activity | 1 | 0.5 | Bridging Metric (product) | 6.00e-02 (4.54e-02 - 6.13e-02) | 2.74e-03 (2.74e-03 - 2.74e-03) | 5.64e-02 (4.32e-02 - 5.79e-02) | 1.85e+01 (1.66e+01 - 2.15e+01) | 9.49e-01 (9.43e-01 - 9.55e-01) |
| Random Subsample High Activity | 1 | 0.75 | Bridging Metric (product) | 5.77e-02 (5.18e-02 - 6.41e-02) | 1.82e-03 (1.82e-03 - 1.82e-03) | 5.53e-02 (4.96e-02 - 6.24e-02) | 3.32e+01 (1.99e+01 - 3.65e+01) | 9.71e-01 (9.51e-01 - 9.73e-01) |
| Random Subsample High Activity | 1 | 0.95 | Bridging Metric (product) | 2.44e-02 (1.21e-02 - 3.62e-02) | 2.39e-04 (2.39e-04 - 2.39e-04) | 2.41e-02 (1.19e-02 - 3.59e-02) | 1.04e+02 (8.11e+01 - 1.29e+02) | 9.91e-01 (9.88e-01 - 9.92e-01) |

**Supplementary Table 3: Regression Coefficients and Correlations between Sampling Schemes and Ground Truth**

| Sampling Scheme | Slope | Intercept | Slope SE | Slope p-value (Wald) | R^2^ |
| --- | --- | --- | --- | --- | --- |
| All Infections | 1.28 | 2.92E-05 | 0.018 | 3.55e-87 | 0.982 |
| Detected Only | 0.97 | -1.54E-06 | 0.022 | 2.10e-40 | 0.976 |
| Symptomatic Only | 1.12 | 4.79E-06 | 0.026 | 5.03e-40 | 0.975 |
| Symptomatic Men | 0.92 | -4.67E-05 | 0.027 | 4.18e-35 | 0.960 |
| High Activity Symptomatic Men | 0.48 | -7.12E-05 | 0.024 | 3.92e-25 | 0.895 |
| Random Subsample High Activity | 0.35 | -1.02E-04 | 0.030 | 2.19e-15 | 0.733 |

**Supplementary Table 4: Description of sampling schemes**

| Sampling Scheme Name | Description |
| --- | --- |
| Detected only | All infections that were detected, either because they were symptomatic and sought care (all symptomatic infections) or were asymptomatic and detected with screening. This excludes infections which recovered naturally. |
| Symptomatic only | Subset of detected infections that were symptomatic. |
| Symptomatic men | Subset of symptomatic infections that occurred in men (MSM, MSMW, MSW). |
| High activity symptomatic men | Subset of symptomatic infections that occurred in male agents who were designated high activity. |
| Random subsample of high activity agents | A random subsample of 25% of all high activity male agents were selected and this includes all of the infections that occurred in those agents. |

**Supplementary Table 5: Kruskal-Wallis Tests by ρ and coverage level**

| ρ | Coverage Level (%) | Statistic | P-adj (BH) | Significant (FDR 0.05) |
| --- | --- | --- | --- | --- |
| 0.05 | 100 | 43.95 | <1e-6 | TRUE |
| 0.05 | 1 | 26.49 | 2.5e-5 | TRUE |
| 0.05 | 2 | 36.14 | <1e-6 | TRUE |
| 0.05 | 3 | 38.84 | <1e-6 | TRUE |
| 0.05 | 4 | 39.22 | <1e-6 | TRUE |
| 0.05 | 5 | 38.93 | <1e-6 | TRUE |
| 0.05 | 10 | 39.92 | <1e-6 | TRUE |
| 0.05 | 25 | 39.12 | <1e-6 | TRUE |
| 0.05 | 50 | 38.92 | <1e-6 | TRUE |
| 0.25 | 100 | 42.92 | <1e-6 | TRUE |
| 0.25 | 1 | 41.80 | <1e-6 | TRUE |
| 0.25 | 2 | 40.90 | <1e-6 | TRUE |
| 0.25 | 3 | 42.29 | <1e-6 | TRUE |
| 0.25 | 4 | 42.85 | <1e-6 | TRUE |
| 0.25 | 5 | 42.40 | <1e-6 | TRUE |
| 0.25 | 10 | 43.07 | <1e-6 | TRUE |
| 0.25 | 25 | 41.03 | <1e-6 | TRUE |
| 0.25 | 50 | 38.91 | <1e-6 | TRUE |
| 0.5 | 100 | 40.14 | <1e-6 | TRUE |
| 0.5 | 1 | 40.18 | <1e-6 | TRUE |
| 0.5 | 2 | 42.23 | <1e-6 | TRUE |
| 0.5 | 3 | 40.89 | <1e-6 | TRUE |
| 0.5 | 4 | 39.74 | <1e-6 | TRUE |
| 0.5 | 5 | 40.05 | <1e-6 | TRUE |
| 0.5 | 10 | 40.24 | <1e-6 | TRUE |
| 0.5 | 25 | 39.18 | <1e-6 | TRUE |
| 0.5 | 50 | 37.63 | <1e-6 | TRUE |
| 0.75 | 100 | 44.21 | <1e-6 | TRUE |
| 0.75 | 1 | 45.69 | <1e-6 | TRUE |
| 0.75 | 2 | 43.74 | <1e-6 | TRUE |
| 0.75 | 3 | 43.12 | <1e-6 | TRUE |
| 0.75 | 4 | 43.83 | <1e-6 | TRUE |
| 0.75 | 5 | 43.58 | <1e-6 | TRUE |
| 0.75 | 10 | 43.48 | <1e-6 | TRUE |
| 0.75 | 25 | 41.37 | <1e-6 | TRUE |
| 0.75 | 50 | 39.89 | <1e-6 | TRUE |
| 0.95 | 100 | 44.48 | <1e-6 | TRUE |
| 0.95 | 1 | 32.90 | 1e-6 | TRUE |
| 0.95 | 2 | 32.46 | 2e-6 | TRUE |
| 0.95 | 3 | 32.87 | 1e-6 | TRUE |
| 0.95 | 4 | 36.30 | <1e-6 | TRUE |
| 0.95 | 5 | 36.92 | <1e-6 | TRUE |
| 0.95 | 10 | 38.54 | <1e-6 | TRUE |
| 0.95 | 25 | 39.29 | <1e-6 | TRUE |
| 0.95 | 50 | 38.87 | <1e-6 | TRUE |

**Supplementary Table 6: Post-hoc Dunn Tests by ρ and coverage level**

| ρ | Coverage Level (%) | Population Comparison | Statistic | Adjusted P Value (BH) | Significant (FDR 0.05) |
| --- | --- | --- | --- | --- | --- |
| 0.05 | 100 | all - detected only | -1.59 | 1.69E-01 | FALSE |
| 0.05 | 100 | all - high activity symptomatic men | -4.38 | 6e-5 | TRUE |
| 0.05 | 100 | detected only - high activity symptomatic men | -2.79 | 1.13E-02 | TRUE |
| 0.05 | 100 | all - random subsample high activity | -5.33 | 2e-6 | TRUE |
| 0.05 | 100 | detected only - random subsample high activity | -3.74 | 6.94e-4 | TRUE |
| 0.05 | 100 | high activity symptomatic men - random subsample high activity | -0.95 | 4.29E-01 | FALSE |
| 0.05 | 100 | all - symptomatic men | -1.91 | 9.40E-02 | FALSE |
| 0.05 | 100 | detected only - symptomatic men | -0.32 | 7.49E-01 | FALSE |
| 0.05 | 100 | high activity symptomatic men - symptomatic men | 2.47 | 2.53E-02 | TRUE |
| 0.05 | 100 | random subsample high activity - symptomatic men | 3.42 | 1.57E-03 | TRUE |
| 0.05 | 100 | all - symptomatic only | -0.70 | 5.16E-01 | FALSE |
| 0.05 | 100 | detected only - symptomatic only | 0.88 | 4.35E-01 | FALSE |
| 0.05 | 100 | high activity symptomatic men - symptomatic only | 3.67 | 7.15e-4 | TRUE |
| 0.05 | 100 | random subsample high activity - symptomatic only | 4.62 | 2.8e-5 | TRUE |
| 0.05 | 100 | symptomatic men - symptomatic only | 1.20 | 3.12E-01 | FALSE |
| 0.05 | 1 | detected only - high activity symptomatic men | -2.59 | 2.38E-02 | TRUE |
| 0.05 | 1 | detected only - random subsample high activity | -3.71 | 1.03E-03 | TRUE |
| 0.05 | 1 | high activity symptomatic men - random subsample high activity | -1.12 | 2.92E-01 | FALSE |
| 0.05 | 1 | detected only - symptomatic men | -1.24 | 2.68E-01 | FALSE |
| 0.05 | 1 | high activity symptomatic men - symptomatic men | 1.35 | 2.53E-01 | FALSE |
| 0.05 | 1 | random subsample high activity - symptomatic men | 2.47 | 2.71E-02 | TRUE |
| 0.05 | 1 | detected only - symptomatic only | 0.72 | 4.71E-01 | FALSE |
| 0.05 | 1 | high activity symptomatic men - symptomatic only | 3.31 | 3.07E-03 | TRUE |
| 0.05 | 1 | random subsample high activity - symptomatic only | 4.43 | 9.3e-5 | TRUE |
| 0.05 | 1 | symptomatic men - symptomatic only | 1.96 | 8.27E-02 | FALSE |
| 0.05 | 2 | detected only - high activity symptomatic men | -3.41 | 1.32E-03 | TRUE |
| 0.05 | 2 | detected only - random subsample high activity | -4.45 | 4.3e-5 | TRUE |
| 0.05 | 2 | high activity symptomatic men - random subsample high activity | -1.04 | 4.24E-01 | FALSE |
| 0.05 | 2 | detected only - symptomatic men | -0.61 | 5.99E-01 | FALSE |
| 0.05 | 2 | high activity symptomatic men - symptomatic men | 2.79 | 8.74E-03 | TRUE |
| 0.05 | 2 | random subsample high activity - symptomatic men | 3.83 | 4.19e-4 | TRUE |
| 0.05 | 2 | detected only - symptomatic only | 0.18 | 8.54E-01 | FALSE |
| 0.05 | 2 | high activity symptomatic men - symptomatic only | 3.59 | 8.29e-4 | TRUE |
| 0.05 | 2 | random subsample high activity - symptomatic only | 4.63 | 3.6e-5 | TRUE |
| 0.05 | 2 | symptomatic men - symptomatic only | 0.80 | 5.31E-01 | FALSE |
| 0.05 | 3 | detected only - high activity symptomatic men | -3.64 | 6.94e-4 | TRUE |
| 0.05 | 3 | detected only - random subsample high activity | -4.60 | 2.1e-5 | TRUE |
| 0.05 | 3 | high activity symptomatic men - random subsample high activity | -0.97 | 3.71E-01 | FALSE |
| 0.05 | 3 | detected only - symptomatic men | -1.35 | 2.21E-01 | FALSE |
| 0.05 | 3 | high activity symptomatic men - symptomatic men | 2.29 | 3.71E-02 | TRUE |
| 0.05 | 3 | random subsample high activity - symptomatic men | 3.25 | 2.29E-03 | TRUE |
| 0.05 | 3 | detected only - symptomatic only | 0.38 | 7.01E-01 | FALSE |
| 0.05 | 3 | high activity symptomatic men - symptomatic only | 4.02 | 1.95e-4 | TRUE |
| 0.05 | 3 | random subsample high activity - symptomatic only | 4.99 | 6e-6 | TRUE |
| 0.05 | 3 | symptomatic men - symptomatic only | 1.73 | 1.19E-01 | FALSE |
| 0.05 | 4 | detected only - high activity symptomatic men | -3.64 | 6.94e-4 | TRUE |
| 0.05 | 4 | detected only - random subsample high activity | -4.88 | 5e-6 | TRUE |
| 0.05 | 4 | high activity symptomatic men - random subsample high activity | -1.24 | 2.38E-01 | FALSE |
| 0.05 | 4 | detected only - symptomatic men | -1.47 | 1.76E-01 | FALSE |
| 0.05 | 4 | high activity symptomatic men - symptomatic men | 2.16 | 5.09E-02 | FALSE |
| 0.05 | 4 | random subsample high activity - symptomatic men | 3.41 | 1.32E-03 | TRUE |
| 0.05 | 4 | detected only - symptomatic only | 0.09 | 9.27E-01 | FALSE |
| 0.05 | 4 | high activity symptomatic men - symptomatic only | 3.73 | 6.45e-4 | TRUE |
| 0.05 | 4 | random subsample high activity - symptomatic only | 4.97 | 5e-6 | TRUE |
| 0.05 | 4 | symptomatic men - symptomatic only | 1.56 | 1.68E-01 | FALSE |
| 0.05 | 5 | detected only - high activity symptomatic men | -3.80 | 4.74e-4 | TRUE |
| 0.05 | 5 | detected only - random subsample high activity | -5.09 | 4e-6 | TRUE |
| 0.05 | 5 | high activity symptomatic men - random subsample high activity | -1.29 | 2.32E-01 | FALSE |
| 0.05 | 5 | detected only - symptomatic men | -1.63 | 1.49E-01 | FALSE |
| 0.05 | 5 | high activity symptomatic men - symptomatic men | 2.18 | 4.90E-02 | TRUE |
| 0.05 | 5 | random subsample high activity - symptomatic men | 3.47 | 1.18E-03 | TRUE |
| 0.05 | 5 | detected only - symptomatic only | -0.37 | 7.13E-01 | FALSE |
| 0.05 | 5 | high activity symptomatic men - symptomatic only | 3.44 | 1.18E-03 | TRUE |
| 0.05 | 5 | random subsample high activity - symptomatic only | 4.72 | 1.2e-5 | TRUE |
| 0.05 | 5 | symptomatic men - symptomatic only | 1.26 | 2.32E-01 | FALSE |
| 0.05 | 10 | detected only - high activity symptomatic men | -3.51 | 1.11E-03 | TRUE |
| 0.05 | 10 | detected only - random subsample high activity | -4.86 | 6e-6 | TRUE |
| 0.05 | 10 | high activity symptomatic men - random subsample high activity | -1.35 | 1.97E-01 | FALSE |
| 0.05 | 10 | detected only - symptomatic men | -1.41 | 1.97E-01 | FALSE |
| 0.05 | 10 | high activity symptomatic men - symptomatic men | 2.10 | 5.93E-02 | FALSE |
| 0.05 | 10 | random subsample high activity - symptomatic men | 3.45 | 1.12E-03 | TRUE |
| 0.05 | 10 | detected only - symptomatic only | 0.28 | 7.82E-01 | FALSE |
| 0.05 | 10 | high activity symptomatic men - symptomatic only | 3.79 | 5.05e-4 | TRUE |
| 0.05 | 10 | random subsample high activity - symptomatic only | 5.14 | 3e-6 | TRUE |
| 0.05 | 10 | symptomatic men - symptomatic only | 1.69 | 1.31E-01 | FALSE |
| 0.05 | 25 | detected only - high activity symptomatic men | -3.21 | 2.69E-03 | TRUE |
| 0.05 | 25 | detected only - random subsample high activity | -4.68 | 1.4e-5 | TRUE |
| 0.05 | 25 | high activity symptomatic men - random subsample high activity | -1.47 | 1.76E-01 | FALSE |
| 0.05 | 25 | detected only - symptomatic men | -1.14 | 2.85E-01 | FALSE |
| 0.05 | 25 | high activity symptomatic men - symptomatic men | 2.07 | 6.40E-02 | FALSE |
| 0.05 | 25 | random subsample high activity - symptomatic men | 3.54 | 9.88e-4 | TRUE |
| 0.05 | 25 | detected only - symptomatic only | 0.58 | 5.60E-01 | FALSE |
| 0.05 | 25 | high activity symptomatic men - symptomatic only | 3.79 | 5.05e-4 | TRUE |
| 0.05 | 25 | random subsample high activity - symptomatic only | 5.26 | 1e-6 | TRUE |
| 0.05 | 25 | symptomatic men - symptomatic only | 1.72 | 1.23E-01 | FALSE |
| 0.05 | 50 | detected only - high activity symptomatic men | -2.93 | 6.78E-03 | TRUE |
| 0.05 | 50 | detected only - random subsample high activity | -4.48 | 3.7e-5 | TRUE |
| 0.05 | 50 | high activity symptomatic men - random subsample high activity | -1.55 | 1.73E-01 | FALSE |
| 0.05 | 50 | detected only - symptomatic men | -0.55 | 5.81E-01 | FALSE |
| 0.05 | 50 | high activity symptomatic men - symptomatic men | 2.38 | 2.90E-02 | TRUE |
| 0.05 | 50 | random subsample high activity - symptomatic men | 3.93 | 2.87e-4 | TRUE |
| 0.05 | 50 | detected only - symptomatic only | 0.83 | 4.53E-01 | FALSE |
| 0.05 | 50 | high activity symptomatic men - symptomatic only | 3.76 | 4.28e-4 | TRUE |
| 0.05 | 50 | random subsample high activity - symptomatic only | 5.31 | 1e-6 | TRUE |
| 0.05 | 50 | symptomatic men - symptomatic only | 1.38 | 2.09E-01 | FALSE |
| 0.25 | 100 | all - detected only | -1.33 | 2.74E-01 | FALSE |
| 0.25 | 100 | all - high activity symptomatic men | -4.30 | 8.5e-5 | TRUE |
| 0.25 | 100 | detected only - high activity symptomatic men | -2.97 | 6.37E-03 | TRUE |
| 0.25 | 100 | all - random subsample high activity | -5.16 | 4e-6 | TRUE |
| 0.25 | 100 | detected only - random subsample high activity | -3.83 | 4.84e-4 | TRUE |
| 0.25 | 100 | high activity symptomatic men - random subsample high activity | -0.86 | 4.89E-01 | FALSE |
| 0.25 | 100 | all - symptomatic men | -1.59 | 1.87E-01 | FALSE |
| 0.25 | 100 | detected only - symptomatic men | -0.26 | 7.98E-01 | FALSE |
| 0.25 | 100 | high activity symptomatic men - symptomatic men | 2.71 | 1.25E-02 | TRUE |
| 0.25 | 100 | random subsample high activity - symptomatic men | 3.57 | 8.85e-4 | TRUE |
| 0.25 | 100 | all - symptomatic only | -0.68 | 5.50E-01 | FALSE |
| 0.25 | 100 | detected only - symptomatic only | 0.65 | 5.50E-01 | FALSE |
| 0.25 | 100 | high activity symptomatic men - symptomatic only | 3.62 | 8.72e-4 | TRUE |
| 0.25 | 100 | random subsample high activity - symptomatic only | 4.48 | 5.6e-5 | TRUE |
| 0.25 | 100 | symptomatic men - symptomatic only | 0.91 | 4.89E-01 | FALSE |
| 0.25 | 1 | detected only - high activity symptomatic men | -3.42 | 1.56E-03 | TRUE |
| 0.25 | 1 | detected only - random subsample high activity | -4.66 | 1.6e-5 | TRUE |
| 0.25 | 1 | high activity symptomatic men - random subsample high activity | -1.24 | 2.38E-01 | FALSE |
| 0.25 | 1 | detected only - symptomatic men | -1.70 | 1.11E-01 | FALSE |
| 0.25 | 1 | high activity symptomatic men - symptomatic men | 1.72 | 1.11E-01 | FALSE |
| 0.25 | 1 | random subsample high activity - symptomatic men | 2.96 | 6.14E-03 | TRUE |
| 0.25 | 1 | detected only - symptomatic only | 0.81 | 4.16E-01 | FALSE |
| 0.25 | 1 | high activity symptomatic men - symptomatic only | 4.23 | 7.7e-5 | TRUE |
| 0.25 | 1 | random subsample high activity - symptomatic only | 5.48 | 0.00E+00 | TRUE |
| 0.25 | 1 | symptomatic men - symptomatic only | 2.52 | 1.98E-02 | TRUE |
| 0.25 | 2 | detected only - high activity symptomatic men | -3.90 | 2.44e-4 | TRUE |
| 0.25 | 2 | detected only - random subsample high activity | -4.62 | 1.9e-5 | TRUE |
| 0.25 | 2 | high activity symptomatic men - random subsample high activity | -0.72 | 5.23E-01 | FALSE |
| 0.25 | 2 | detected only - symptomatic men | -1.83 | 8.49E-02 | FALSE |
| 0.25 | 2 | high activity symptomatic men - symptomatic men | 2.07 | 5.48E-02 | FALSE |
| 0.25 | 2 | random subsample high activity - symptomatic men | 2.79 | 1.05E-02 | TRUE |
| 0.25 | 2 | detected only - symptomatic only | 0.44 | 6.56E-01 | FALSE |
| 0.25 | 2 | high activity symptomatic men - symptomatic only | 4.34 | 4.7e-5 | TRUE |
| 0.25 | 2 | random subsample high activity - symptomatic only | 5.06 | 4e-6 | TRUE |
| 0.25 | 2 | symptomatic men - symptomatic only | 2.27 | 3.87E-02 | TRUE |
| 0.25 | 3 | detected only - high activity symptomatic men | -3.70 | 5.46e-4 | TRUE |
| 0.25 | 3 | detected only - random subsample high activity | -4.94 | 4e-6 | TRUE |
| 0.25 | 3 | high activity symptomatic men - random subsample high activity | -1.24 | 2.38E-01 | FALSE |
| 0.25 | 3 | detected only - symptomatic men | -1.86 | 8.21E-02 | FALSE |
| 0.25 | 3 | high activity symptomatic men - symptomatic men | 1.84 | 8.21E-02 | FALSE |
| 0.25 | 3 | random subsample high activity - symptomatic men | 3.08 | 4.10E-03 | TRUE |
| 0.25 | 3 | detected only - symptomatic only | 0.37 | 7.13E-01 | FALSE |
| 0.25 | 3 | high activity symptomatic men - symptomatic only | 4.06 | 1.6e-4 | TRUE |
| 0.25 | 3 | random subsample high activity - symptomatic only | 5.31 | 1e-6 | TRUE |
| 0.25 | 3 | symptomatic men - symptomatic only | 2.22 | 4.36E-02 | TRUE |
| 0.25 | 4 | detected only - high activity symptomatic men | -3.60 | 7.81e-4 | TRUE |
| 0.25 | 4 | detected only - random subsample high activity | -4.99 | 3e-6 | TRUE |
| 0.25 | 4 | high activity symptomatic men - random subsample high activity | -1.38 | 1.86E-01 | FALSE |
| 0.25 | 4 | detected only - symptomatic men | -1.81 | 9.09E-02 | FALSE |
| 0.25 | 4 | high activity symptomatic men - symptomatic men | 1.79 | 9.09E-02 | FALSE |
| 0.25 | 4 | random subsample high activity - symptomatic men | 3.18 | 2.99E-03 | TRUE |
| 0.25 | 4 | detected only - symptomatic only | 0.43 | 6.68E-01 | FALSE |
| 0.25 | 4 | high activity symptomatic men - symptomatic only | 4.03 | 1.83e-4 | TRUE |
| 0.25 | 4 | random subsample high activity - symptomatic only | 5.41 | 1e-6 | TRUE |
| 0.25 | 4 | symptomatic men - symptomatic only | 2.24 | 4.19E-02 | TRUE |
| 0.25 | 5 | detected only - high activity symptomatic men | -3.54 | 9.88e-4 | TRUE |
| 0.25 | 5 | detected only - random subsample high activity | -4.85 | 6e-6 | TRUE |
| 0.25 | 5 | high activity symptomatic men - random subsample high activity | -1.30 | 2.14E-01 | FALSE |
| 0.25 | 5 | detected only - symptomatic men | -1.70 | 1.11E-01 | FALSE |
| 0.25 | 5 | high activity symptomatic men - symptomatic men | 1.84 | 9.38E-02 | FALSE |
| 0.25 | 5 | random subsample high activity - symptomatic men | 3.14 | 3.33E-03 | TRUE |
| 0.25 | 5 | detected only - symptomatic only | 0.58 | 5.60E-01 | FALSE |
| 0.25 | 5 | high activity symptomatic men - symptomatic only | 4.13 | 1.23e-4 | TRUE |
| 0.25 | 5 | random subsample high activity - symptomatic only | 5.43 | 1e-6 | TRUE |
| 0.25 | 5 | symptomatic men - symptomatic only | 2.29 | 3.71E-02 | TRUE |
| 0.25 | 10 | detected only - high activity symptomatic men | -3.59 | 8.29e-4 | TRUE |
| 0.25 | 10 | detected only - random subsample high activity | -5.12 | 2e-6 | TRUE |
| 0.25 | 10 | high activity symptomatic men - random subsample high activity | -1.53 | 1.39E-01 | FALSE |
| 0.25 | 10 | detected only - symptomatic men | -1.86 | 9.06E-02 | FALSE |
| 0.25 | 10 | high activity symptomatic men - symptomatic men | 1.73 | 1.04E-01 | FALSE |
| 0.25 | 10 | random subsample high activity - symptomatic men | 3.27 | 2.17E-03 | TRUE |
| 0.25 | 10 | detected only - symptomatic only | 0.29 | 7.71E-01 | FALSE |
| 0.25 | 10 | high activity symptomatic men - symptomatic only | 3.88 | 3.47e-4 | TRUE |
| 0.25 | 10 | random subsample high activity - symptomatic only | 5.41 | 1e-6 | TRUE |
| 0.25 | 10 | symptomatic men - symptomatic only | 2.15 | 5.29E-02 | FALSE |
| 0.25 | 25 | detected only - high activity symptomatic men | -3.30 | 1.95E-03 | TRUE |
| 0.25 | 25 | detected only - random subsample high activity | -4.77 | 9e-6 | TRUE |
| 0.25 | 25 | high activity symptomatic men - random subsample high activity | -1.47 | 1.76E-01 | FALSE |
| 0.25 | 25 | detected only - symptomatic men | -1.23 | 2.44E-01 | FALSE |
| 0.25 | 25 | high activity symptomatic men - symptomatic men | 2.07 | 6.40E-02 | FALSE |
| 0.25 | 25 | random subsample high activity - symptomatic men | 3.54 | 9.88e-4 | TRUE |
| 0.25 | 25 | detected only - symptomatic only | 0.63 | 5.29E-01 | FALSE |
| 0.25 | 25 | high activity symptomatic men - symptomatic only | 3.93 | 2.87e-4 | TRUE |
| 0.25 | 25 | random subsample high activity - symptomatic only | 5.40 | 1e-6 | TRUE |
| 0.25 | 25 | symptomatic men - symptomatic only | 1.86 | 9.06E-02 | FALSE |
| 0.25 | 50 | detected only - high activity symptomatic men | -3.19 | 2.84E-03 | TRUE |
| 0.25 | 50 | detected only - random subsample high activity | -4.57 | 2.4e-5 | TRUE |
| 0.25 | 50 | high activity symptomatic men - random subsample high activity | -1.38 | 2.39E-01 | FALSE |
| 0.25 | 50 | detected only - symptomatic men | -0.72 | 5.23E-01 | FALSE |
| 0.25 | 50 | high activity symptomatic men - symptomatic men | 2.47 | 2.25E-02 | TRUE |
| 0.25 | 50 | random subsample high activity - symptomatic men | 3.85 | 3.93e-4 | TRUE |
| 0.25 | 50 | detected only - symptomatic only | 0.58 | 5.60E-01 | FALSE |
| 0.25 | 50 | high activity symptomatic men - symptomatic only | 3.77 | 4.02e-4 | TRUE |
| 0.25 | 50 | random subsample high activity - symptomatic only | 5.15 | 3e-6 | TRUE |
| 0.25 | 50 | symptomatic men - symptomatic only | 1.30 | 2.40E-01 | FALSE |
| 0.5 | 100 | all - detected only | -1.22 | 3.36E-01 | FALSE |
| 0.5 | 100 | all - high activity symptomatic men | -4.01 | 3.07e-4 | TRUE |
| 0.5 | 100 | detected only - high activity symptomatic men | -2.79 | 1.13E-02 | TRUE |
| 0.5 | 100 | all - random subsample high activity | -4.93 | 1.2e-5 | TRUE |
| 0.5 | 100 | detected only - random subsample high activity | -3.71 | 7.68e-4 | TRUE |
| 0.5 | 100 | high activity symptomatic men - random subsample high activity | -0.92 | 4.86E-01 | FALSE |
| 0.5 | 100 | all - symptomatic men | -1.28 | 3.34E-01 | FALSE |
| 0.5 | 100 | detected only - symptomatic men | -0.06 | 9.49E-01 | FALSE |
| 0.5 | 100 | high activity symptomatic men - symptomatic men | 2.73 | 1.20E-02 | TRUE |
| 0.5 | 100 | random subsample high activity - symptomatic men | 3.65 | 7.9e-4 | TRUE |
| 0.5 | 100 | all - symptomatic only | -0.47 | 6.81E-01 | FALSE |
| 0.5 | 100 | detected only - symptomatic only | 0.74 | 5.28E-01 | FALSE |
| 0.5 | 100 | high activity symptomatic men - symptomatic only | 3.53 | 1.02E-03 | TRUE |
| 0.5 | 100 | random subsample high activity - symptomatic only | 4.46 | 6.3e-5 | TRUE |
| 0.5 | 100 | symptomatic men - symptomatic only | 0.8066323435772448 | 5.25E-01 | FALSE |
| 0.5 | 1 | detected only - high activity symptomatic men | -3.79 | 3.78e-4 | TRUE |
| 0.5 | 1 | detected only - random subsample high activity | -4.48 | 3.7e-5 | TRUE |
| 0.5 | 1 | high activity symptomatic men - random subsample high activity | -0.69 | 5.44E-01 | FALSE |
| 0.5 | 1 | detected only - symptomatic men | -1.76 | 9.71E-02 | FALSE |
| 0.5 | 1 | high activity symptomatic men - symptomatic men | 2.02 | 6.12E-02 | FALSE |
| 0.5 | 1 | random subsample high activity - symptomatic men | 2.72 | 1.32E-02 | TRUE |
| 0.5 | 1 | detected only - symptomatic only | 0.60 | 5.50E-01 | FALSE |
| 0.5 | 1 | high activity symptomatic men - symptomatic only | 4.39 | 3.8e-5 | TRUE |
| 0.5 | 1 | random subsample high activity - symptomatic only | 5.08 | 4e-6 | TRUE |
| 0.5 | 1 | symptomatic men - symptomatic only | 2.36 | 3.03E-02 | TRUE |
| 0.5 | 2 | detected only - high activity symptomatic men | -3.64 | 6.94e-4 | TRUE |
| 0.5 | 2 | detected only - random subsample high activity | -4.92 | 4e-6 | TRUE |
| 0.5 | 2 | high activity symptomatic men - random subsample high activity | -1.29 | 2.20E-01 | FALSE |
| 0.5 | 2 | detected only - symptomatic men | -1.75 | 1.00E-01 | FALSE |
| 0.5 | 2 | high activity symptomatic men - symptomatic men | 1.89 | 8.46E-02 | FALSE |
| 0.5 | 2 | random subsample high activity - symptomatic men | 3.18 | 2.99E-03 | TRUE |
| 0.5 | 2 | detected only - symptomatic only | 0.41 | 6.79E-01 | FALSE |
| 0.5 | 2 | high activity symptomatic men - symptomatic only | 4.05 | 1.71e-4 | TRUE |
| 0.5 | 2 | random subsample high activity - symptomatic only | 5.34 | 1e-6 | TRUE |
| 0.5 | 2 | symptomatic men - symptomatic only | 2.16 | 5.09E-02 | FALSE |
| 0.5 | 3 | detected only - high activity symptomatic men | -3.44 | 1.48E-03 | TRUE |
| 0.5 | 3 | detected only - random subsample high activity | -4.88 | 5e-6 | TRUE |
| 0.5 | 3 | high activity symptomatic men - random subsample high activity | -1.44 | 1.66E-01 | FALSE |
| 0.5 | 3 | detected only - symptomatic men | -1.56 | 1.47E-01 | FALSE |
| 0.5 | 3 | high activity symptomatic men - symptomatic men | 1.87 | 8.76E-02 | FALSE |
| 0.5 | 3 | random subsample high activity - symptomatic men | 3.31 | 1.84E-03 | TRUE |
| 0.5 | 3 | detected only - symptomatic only | 0.44 | 6.56E-01 | FALSE |
| 0.5 | 3 | high activity symptomatic men - symptomatic only | 3.88 | 3.47e-4 | TRUE |
| 0.5 | 3 | random subsample high activity - symptomatic only | 5.32 | 1e-6 | TRUE |
| 0.5 | 3 | symptomatic men - symptomatic only | 2.01 | 7.41E-02 | FALSE |
| 0.5 | 4 | detected only - high activity symptomatic men | -3.53 | 1.05E-03 | TRUE |
| 0.5 | 4 | detected only - random subsample high activity | -4.71 | 1.2e-5 | TRUE |
| 0.5 | 4 | high activity symptomatic men - random subsample high activity | -1.18 | 2.64E-01 | FALSE |
| 0.5 | 4 | detected only - symptomatic men | -1.41 | 1.98E-01 | FALSE |
| 0.5 | 4 | high activity symptomatic men - symptomatic men | 2.12 | 5.71E-02 | FALSE |
| 0.5 | 4 | random subsample high activity - symptomatic men | 3.30 | 1.95E-03 | TRUE |
| 0.5 | 4 | detected only - symptomatic only | 0.44 | 6.56E-01 | FALSE |
| 0.5 | 4 | high activity symptomatic men - symptomatic only | 3.97 | 2.37e-4 | TRUE |
| 0.5 | 4 | random subsample high activity - symptomatic only | 5.15 | 3e-6 | TRUE |
| 0.5 | 4 | symptomatic men - symptomatic only | 1.86 | 9.06E-02 | FALSE |
| 0.5 | 5 | detected only - high activity symptomatic men | -3.48 | 1.24E-03 | TRUE |
| 0.5 | 5 | detected only - random subsample high activity | -4.77 | 9e-6 | TRUE |
| 0.5 | 5 | high activity symptomatic men - random subsample high activity | -1.29 | 2.20E-01 | FALSE |
| 0.5 | 5 | detected only - symptomatic men | -1.47 | 1.76E-01 | FALSE |
| 0.5 | 5 | high activity symptomatic men - symptomatic men | 2.01 | 7.41E-02 | FALSE |
| 0.5 | 5 | random subsample high activity - symptomatic men | 3.30 | 1.95E-03 | TRUE |
| 0.5 | 5 | detected only - symptomatic only | 0.44 | 6.56E-01 | FALSE |
| 0.5 | 5 | high activity symptomatic men - symptomatic only | 3.93 | 2.87e-4 | TRUE |
| 0.5 | 5 | random subsample high activity - symptomatic only | 5.22 | 2e-6 | TRUE |
| 0.5 | 5 | symptomatic men - symptomatic only | 1.92 | 7.88E-02 | FALSE |
| 0.5 | 10 | detected only - high activity symptomatic men | -3.39 | 1.40E-03 | TRUE |
| 0.5 | 10 | detected only - random subsample high activity | -4.79 | 9e-6 | TRUE |
| 0.5 | 10 | high activity symptomatic men - random subsample high activity | -1.40 | 1.91E-01 | FALSE |
| 0.5 | 10 | detected only - symptomatic men | -1.37 | 1.91E-01 | FALSE |
| 0.5 | 10 | high activity symptomatic men - symptomatic men | 2.02 | 7.15E-02 | FALSE |
| 0.5 | 10 | random subsample high activity - symptomatic men | 3.42 | 1.40E-03 | TRUE |
| 0.5 | 10 | detected only - symptomatic only | 0.49 | 6.24E-01 | FALSE |
| 0.5 | 10 | high activity symptomatic men - symptomatic only | 3.88 | 3.47e-4 | TRUE |
| 0.5 | 10 | random subsample high activity - symptomatic only | 5.28 | 1e-6 | TRUE |
| 0.5 | 10 | symptomatic men - symptomatic only | 1.86 | 9.06E-02 | FALSE |
| 0.5 | 25 | detected only - high activity symptomatic men | -3.19 | 2.84E-03 | TRUE |
| 0.5 | 25 | detected only - random subsample high activity | -4.77 | 9e-6 | TRUE |
| 0.5 | 25 | high activity symptomatic men - random subsample high activity | -1.58 | 1.63E-01 | FALSE |
| 0.5 | 25 | detected only - symptomatic men | -0.98 | 3.62E-01 | FALSE |
| 0.5 | 25 | high activity symptomatic men - symptomatic men | 2.21 | 4.53E-02 | TRUE |
| 0.5 | 25 | random subsample high activity - symptomatic men | 3.79 | 5.05e-4 | TRUE |
| 0.5 | 25 | detected only - symptomatic only | 0.43 | 6.68E-01 | FALSE |
| 0.5 | 25 | high activity symptomatic men - symptomatic only | 3.62 | 7.36e-4 | TRUE |
| 0.5 | 25 | random subsample high activity - symptomatic only | 5.20 | 2e-6 | TRUE |
| 0.5 | 25 | symptomatic men - symptomatic only | 1.41 | 1.98E-01 | FALSE |
| 0.5 | 50 | detected only - high activity symptomatic men | -3.01 | 5.29E-03 | TRUE |
| 0.5 | 50 | detected only - random subsample high activity | -4.49 | 3.5e-5 | TRUE |
| 0.5 | 50 | high activity symptomatic men - random subsample high activity | -1.49 | 1.95E-01 | FALSE |
| 0.5 | 50 | detected only - symptomatic men | -0.54 | 5.91E-01 | FALSE |
| 0.5 | 50 | high activity symptomatic men - symptomatic men | 2.47 | 2.25E-02 | TRUE |
| 0.5 | 50 | random subsample high activity - symptomatic men | 3.96 | 2.52e-4 | TRUE |
| 0.5 | 50 | detected only - symptomatic only | 0.60 | 5.91E-01 | FALSE |
| 0.5 | 50 | high activity symptomatic men - symptomatic only | 3.60 | 7.81e-4 | TRUE |
| 0.5 | 50 | random subsample high activity - symptomatic only | 5.09 | 4e-6 | TRUE |
| 0.5 | 50 | symptomatic men - symptomatic only | 1.14 | 3.20E-01 | FALSE |
| 0.75 | 100 | all - detected only | -1.28 | 2.73E-01 | FALSE |
| 0.75 | 100 | all - high activity symptomatic men | -4.31 | 8e-5 | TRUE |
| 0.75 | 100 | detected only - high activity symptomatic men | -3.03 | 5.16E-03 | TRUE |
| 0.75 | 100 | all - random subsample high activity | -5.33 | 2e-6 | TRUE |
| 0.75 | 100 | detected only - random subsample high activity | -4.05 | 1.95e-4 | TRUE |
| 0.75 | 100 | high activity symptomatic men - random subsample high activity | -1.01 | 3.90E-01 | FALSE |
| 0.75 | 100 | all - symptomatic men | -2.20 | 5.18E-02 | FALSE |
| 0.75 | 100 | detected only - symptomatic men | -0.92 | 4.11E-01 | FALSE |
| 0.75 | 100 | high activity symptomatic men - symptomatic men | 2.11 | 5.77E-02 | FALSE |
| 0.75 | 100 | random subsample high activity - symptomatic men | 3.12 | 4.46E-03 | TRUE |
| 0.75 | 100 | all - symptomatic only | -0.70 | 5.16E-01 | FALSE |
| 0.75 | 100 | detected only - symptomatic only | 0.58 | 5.65E-01 | FALSE |
| 0.75 | 100 | high activity symptomatic men - symptomatic only | 3.61 | 9.16e-4 | TRUE |
| 0.75 | 100 | random subsample high activity - symptomatic only | 4.62 | 2.8e-5 | TRUE |
| 0.75 | 100 | symptomatic men - symptomatic only | 1.50 | 2.01E-01 | FALSE |
| 0.75 | 1 | detected only - high activity symptomatic men | -3.31 | 2.31E-03 | TRUE |
| 0.75 | 1 | detected only - random subsample high activity | -4.69 | 1.3e-5 | TRUE |
| 0.75 | 1 | high activity symptomatic men - random subsample high activity | -1.38 | 1.86E-01 | FALSE |
| 0.75 | 1 | detected only - symptomatic men | -1.70 | 1.27E-01 | FALSE |
| 0.75 | 1 | high activity symptomatic men - symptomatic men | 1.61 | 1.34E-01 | FALSE |
| 0.75 | 1 | random subsample high activity - symptomatic men | 2.99 | 5.56E-03 | TRUE |
| 0.75 | 1 | detected only - symptomatic only | 1.20 | 2.32E-01 | FALSE |
| 0.75 | 1 | high activity symptomatic men - symptomatic only | 4.51 | 2.2e-5 | TRUE |
| 0.75 | 1 | random subsample high activity - symptomatic only | 5.89 | 0.00E+00 | TRUE |
| 0.75 | 1 | symptomatic men - symptomatic only | 2.90 | 6.24E-03 | TRUE |
| 0.75 | 2 | detected only - high activity symptomatic men | -3.73 | 4.84e-4 | TRUE |
| 0.75 | 2 | detected only - random subsample high activity | -4.89 | 5e-6 | TRUE |
| 0.75 | 2 | high activity symptomatic men - random subsample high activity | -1.17 | 2.71E-01 | FALSE |
| 0.75 | 2 | detected only - symptomatic men | -1.98 | 6.83E-02 | FALSE |
| 0.75 | 2 | high activity symptomatic men - symptomatic men | 1.75 | 1.00E-01 | FALSE |
| 0.75 | 2 | random subsample high activity - symptomatic men | 2.91 | 7.13E-03 | TRUE |
| 0.75 | 2 | detected only - symptomatic only | 0.55 | 5.81E-01 | FALSE |
| 0.75 | 2 | high activity symptomatic men - symptomatic only | 4.28 | 6.2e-5 | TRUE |
| 0.75 | 2 | random subsample high activity - symptomatic only | 5.45 | 1e-6 | TRUE |
| 0.75 | 2 | symptomatic men - symptomatic only | 2.53 | 1.90E-02 | TRUE |
| 0.75 | 3 | detected only - high activity symptomatic men | -3.65 | 6.54e-4 | TRUE |
| 0.75 | 3 | detected only - random subsample high activity | -4.79 | 9e-6 | TRUE |
| 0.75 | 3 | high activity symptomatic men - random subsample high activity | -1.14 | 2.85E-01 | FALSE |
| 0.75 | 3 | detected only - symptomatic men | -1.81 | 8.79E-02 | FALSE |
| 0.75 | 3 | high activity symptomatic men - symptomatic men | 1.84 | 8.79E-02 | FALSE |
| 0.75 | 3 | random subsample high activity - symptomatic men | 2.98 | 5.84E-03 | TRUE |
| 0.75 | 3 | detected only - symptomatic only | 0.66 | 5.10E-01 | FALSE |
| 0.75 | 3 | high activity symptomatic men - symptomatic only | 4.31 | 5.4e-5 | TRUE |
| 0.75 | 3 | random subsample high activity - symptomatic only | 5.45 | 1e-6 | TRUE |
| 0.75 | 3 | symptomatic men - symptomatic only | 2.47 | 2.25E-02 | TRUE |
| 0.75 | 4 | detected only - high activity symptomatic men | -3.42 | 1.56E-03 | TRUE |
| 0.75 | 4 | detected only - random subsample high activity | -4.86 | 6e-6 | TRUE |
| 0.75 | 4 | high activity symptomatic men - random subsample high activity | -1.44 | 1.66E-01 | FALSE |
| 0.75 | 4 | detected only - symptomatic men | -1.70 | 1.11E-01 | FALSE |
| 0.75 | 4 | high activity symptomatic men - symptomatic men | 1.72 | 1.11E-01 | FALSE |
| 0.75 | 4 | random subsample high activity - symptomatic men | 3.16 | 3.16E-03 | TRUE |
| 0.75 | 4 | detected only - symptomatic only | 0.78 | 4.34E-01 | FALSE |
| 0.75 | 4 | high activity symptomatic men - symptomatic only | 4.20 | 8.8e-5 | TRUE |
| 0.75 | 4 | random subsample high activity - symptomatic only | 5.64 | 0.00E+00 | TRUE |
| 0.75 | 4 | symptomatic men - symptomatic only | 2.48 | 2.16E-02 | TRUE |
| 0.75 | 5 | detected only - high activity symptomatic men | -3.62 | 7.36e-4 | TRUE |
| 0.75 | 5 | detected only - random subsample high activity | -4.94 | 4e-6 | TRUE |
| 0.75 | 5 | high activity symptomatic men - random subsample high activity | -1.32 | 2.08E-01 | FALSE |
| 0.75 | 5 | detected only - symptomatic men | -1.89 | 8.46E-02 | FALSE |
| 0.75 | 5 | high activity symptomatic men - symptomatic men | 1.73 | 1.04E-01 | FALSE |
| 0.75 | 5 | random subsample high activity - symptomatic men | 3.05 | 4.54E-03 | TRUE |
| 0.75 | 5 | detected only - symptomatic only | 0.55 | 5.81E-01 | FALSE |
| 0.75 | 5 | high activity symptomatic men - symptomatic only | 4.17 | 1.01e-4 | TRUE |
| 0.75 | 5 | random subsample high activity - symptomatic only | 5.49 | 0.00E+00 | TRUE |
| 0.75 | 5 | symptomatic men - symptomatic only | 2.44 | 2.46E-02 | TRUE |
| 0.75 | 10 | detected only - high activity symptomatic men | -3.50 | 1.18E-03 | TRUE |
| 0.75 | 10 | detected only - random subsample high activity | -4.85 | 6e-6 | TRUE |
| 0.75 | 10 | high activity symptomatic men - random subsample high activity | -1.35 | 1.97E-01 | FALSE |
| 0.75 | 10 | detected only - symptomatic men | -1.73 | 1.04E-01 | FALSE |
| 0.75 | 10 | high activity symptomatic men - symptomatic men | 1.76 | 1.04E-01 | FALSE |
| 0.75 | 10 | random subsample high activity - symptomatic men | 3.11 | 3.69E-03 | TRUE |
| 0.75 | 10 | detected only - symptomatic only | 0.72 | 4.71E-01 | FALSE |
| 0.75 | 10 | high activity symptomatic men - symptomatic only | 4.22 | 8.2e-5 | TRUE |
| 0.75 | 10 | random subsample high activity - symptomatic only | 5.57 | 0.00E+00 | TRUE |
| 0.75 | 10 | symptomatic men - symptomatic only | 2.45 | 2.35E-02 | TRUE |
| 0.75 | 25 | detected only - high activity symptomatic men | -3.42 | 1.56E-03 | TRUE |
| 0.75 | 25 | detected only - random subsample high activity | -4.68 | 1.4e-5 | TRUE |
| 0.75 | 25 | high activity symptomatic men - random subsample high activity | -1.26 | 2.32E-01 | FALSE |
| 0.75 | 25 | detected only - symptomatic men | -1.37 | 2.15E-01 | FALSE |
| 0.75 | 25 | high activity symptomatic men - symptomatic men | 2.06 | 5.69E-02 | FALSE |
| 0.75 | 25 | random subsample high activity - symptomatic men | 3.31 | 1.84E-03 | TRUE |
| 0.75 | 25 | detected only - symptomatic only | 0.72 | 4.71E-01 | FALSE |
| 0.75 | 25 | high activity symptomatic men - symptomatic only | 4.14 | 1.15e-4 | TRUE |
| 0.75 | 25 | random subsample high activity - symptomatic only | 5.40 | 1e-6 | TRUE |
| 0.75 | 25 | symptomatic men - symptomatic only | 2.09 | 5.69E-02 | FALSE |
| 0.75 | 50 | detected only - high activity symptomatic men | -3.30 | 1.95E-03 | TRUE |
| 0.75 | 50 | detected only - random subsample high activity | -4.59 | 2.3e-5 | TRUE |
| 0.75 | 50 | high activity symptomatic men - random subsample high activity | -1.29 | 2.47E-01 | FALSE |
| 0.75 | 50 | detected only - symptomatic men | -1.01 | 3.46E-01 | FALSE |
| 0.75 | 50 | high activity symptomatic men - symptomatic men | 2.29 | 3.71E-02 | TRUE |
| 0.75 | 50 | random subsample high activity - symptomatic men | 3.57 | 8.79e-4 | TRUE |
| 0.75 | 50 | detected only - symptomatic only | 0.69 | 4.90E-01 | FALSE |
| 0.75 | 50 | high activity symptomatic men - symptomatic only | 3.99 | 2.22e-4 | TRUE |
| 0.75 | 50 | random subsample high activity - symptomatic only | 5.28 | 1e-6 | TRUE |
| 0.75 | 50 | symptomatic men - symptomatic only | 1.70 | 1.27E-01 | FALSE |
| 0.95 | 100 | all - detected only | -1.25 | 2.86E-01 | FALSE |
| 0.95 | 100 | all - high activity symptomatic men | -4.39 | 5.6e-5 | TRUE |
| 0.95 | 100 | detected only - high activity symptomatic men | -3.14 | 3.66E-03 | TRUE |
| 0.95 | 100 | all - random subsample high activity | -5.26 | 2e-6 | TRUE |
| 0.95 | 100 | detected only - random subsample high activity | -4.01 | 2.3e-4 | TRUE |
| 0.95 | 100 | high activity symptomatic men - random subsample high activity | -0.87 | 4.59E-01 | FALSE |
| 0.95 | 100 | all - symptomatic men | -2.10 | 5.96E-02 | FALSE |
| 0.95 | 100 | detected only - symptomatic men | -0.85 | 4.59E-01 | FALSE |
| 0.95 | 100 | high activity symptomatic men - symptomatic men | 2.29 | 4.11E-02 | TRUE |
| 0.95 | 100 | random subsample high activity - symptomatic men | 3.16 | 3.66E-03 | TRUE |
| 0.95 | 100 | all - symptomatic only | -0.67 | 5.42E-01 | FALSE |
| 0.95 | 100 | detected only - symptomatic only | 0.59 | 5.56E-01 | FALSE |
| 0.95 | 100 | high activity symptomatic men - symptomatic only | 3.73 | 5.84e-4 | TRUE |
| 0.95 | 100 | random subsample high activity - symptomatic only | 4.60 | 3.2e-5 | TRUE |
| 0.95 | 100 | symptomatic men - symptomatic only | 1.43 | 2.27E-01 | FALSE |
| 0.95 | 1 | detected only - high activity symptomatic men | -3.13 | 3.51E-03 | TRUE |
| 0.95 | 1 | detected only - random subsample high activity | -4.46 | 4e-5 | TRUE |
| 0.95 | 1 | high activity symptomatic men - random subsample high activity | -1.33 | 2.14E-01 | FALSE |
| 0.95 | 1 | detected only - symptomatic men | -1.30 | 2.14E-01 | FALSE |
| 0.95 | 1 | high activity symptomatic men - symptomatic men | 1.83 | 1.13E-01 | FALSE |
| 0.95 | 1 | random subsample high activity - symptomatic men | 3.16 | 3.51E-03 | TRUE |
| 0.95 | 1 | detected only - symptomatic only | 0.23 | 8.18E-01 | FALSE |
| 0.95 | 1 | high activity symptomatic men - symptomatic only | 3.36 | 2.61E-03 | TRUE |
| 0.95 | 1 | random subsample high activity - symptomatic only | 4.69 | 2.7e-5 | TRUE |
| 0.95 | 1 | symptomatic men - symptomatic only | 1.53 | 1.79E-01 | FALSE |
| 0.95 | 2 | detected only - high activity symptomatic men | -3.34 | 2.07E-03 | TRUE |
| 0.95 | 2 | detected only - random subsample high activity | -4.40 | 5.4e-5 | TRUE |
| 0.95 | 2 | high activity symptomatic men - random subsample high activity | -1.06 | 3.22E-01 | FALSE |
| 0.95 | 2 | detected only - symptomatic men | -1.52 | 1.61E-01 | FALSE |
| 0.95 | 2 | high activity symptomatic men - symptomatic men | 1.83 | 1.13E-01 | FALSE |
| 0.95 | 2 | random subsample high activity - symptomatic men | 2.88 | 7.86E-03 | TRUE |
| 0.95 | 2 | detected only - symptomatic only | 0.14 | 8.90E-01 | FALSE |
| 0.95 | 2 | high activity symptomatic men - symptomatic only | 3.48 | 1.66E-03 | TRUE |
| 0.95 | 2 | random subsample high activity - symptomatic only | 4.54 | 5.4e-5 | TRUE |
| 0.95 | 2 | symptomatic men - symptomatic only | 1.66 | 1.39E-01 | FALSE |
| 0.95 | 3 | detected only - high activity symptomatic men | -3.34 | 2.07E-03 | TRUE |
| 0.95 | 3 | detected only - random subsample high activity | -4.40 | 5.4e-5 | TRUE |
| 0.95 | 3 | high activity symptomatic men - random subsample high activity | -1.06 | 3.22E-01 | FALSE |
| 0.95 | 3 | detected only - symptomatic men | -1.40 | 2.03E-01 | FALSE |
| 0.95 | 3 | high activity symptomatic men - symptomatic men | 1.95 | 8.57E-02 | FALSE |
| 0.95 | 3 | random subsample high activity - symptomatic men | 3.01 | 5.29E-03 | TRUE |
| 0.95 | 3 | detected only - symptomatic only | 0.17 | 8.66E-01 | FALSE |
| 0.95 | 3 | high activity symptomatic men - symptomatic only | 3.51 | 1.48E-03 | TRUE |
| 0.95 | 3 | random subsample high activity - symptomatic only | 4.57 | 4.9e-5 | TRUE |
| 0.95 | 3 | symptomatic men - symptomatic only | 1.56 | 1.68E-01 | FALSE |
| 0.95 | 4 | detected only - high activity symptomatic men | -3.42 | 1.56E-03 | TRUE |
| 0.95 | 4 | detected only - random subsample high activity | -4.59 | 2.3e-5 | TRUE |
| 0.95 | 4 | high activity symptomatic men - random subsample high activity | -1.17 | 2.71E-01 | FALSE |
| 0.95 | 4 | detected only - symptomatic men | -1.50 | 1.66E-01 | FALSE |
| 0.95 | 4 | high activity symptomatic men - symptomatic men | 1.92 | 9.20E-02 | FALSE |
| 0.95 | 4 | random subsample high activity - symptomatic men | 3.08 | 4.10E-03 | TRUE |
| 0.95 | 4 | detected only - symptomatic only | 0.31 | 7.59E-01 | FALSE |
| 0.95 | 4 | high activity symptomatic men - symptomatic only | 3.73 | 6.45e-4 | TRUE |
| 0.95 | 4 | random subsample high activity - symptomatic only | 4.89 | 1e-5 | TRUE |
| 0.95 | 4 | symptomatic men - symptomatic only | 1.81 | 1.00E-01 | FALSE |
| 0.95 | 5 | detected only - high activity symptomatic men | -3.60 | 7.81e-4 | TRUE |
| 0.95 | 5 | detected only - random subsample high activity | -4.49 | 3.5e-5 | TRUE |
| 0.95 | 5 | high activity symptomatic men - random subsample high activity | -0.89 | 4.15E-01 | FALSE |
| 0.95 | 5 | detected only - symptomatic men | -1.35 | 2.21E-01 | FALSE |
| 0.95 | 5 | high activity symptomatic men - symptomatic men | 2.25 | 4.02E-02 | TRUE |
| 0.95 | 5 | random subsample high activity - symptomatic men | 3.14 | 3.33E-03 | TRUE |
| 0.95 | 5 | detected only - symptomatic only | 0.32 | 7.47E-01 | FALSE |
| 0.95 | 5 | high activity symptomatic men - symptomatic only | 3.93 | 2.87e-4 | TRUE |
| 0.95 | 5 | random subsample high activity - symptomatic only | 4.82 | 1.5e-5 | TRUE |
| 0.95 | 5 | symptomatic men - symptomatic only | 1.67 | 1.35E-01 | FALSE |
| 0.95 | 10 | detected only - high activity symptomatic men | -3.57 | 8.79e-4 | TRUE |
| 0.95 | 10 | detected only - random subsample high activity | -4.63 | 1.8e-5 | TRUE |
| 0.95 | 10 | high activity symptomatic men - random subsample high activity | -1.06 | 3.22E-01 | FALSE |
| 0.95 | 10 | detected only - symptomatic men | -1.26 | 2.61E-01 | FALSE |
| 0.95 | 10 | high activity symptomatic men - symptomatic men | 2.32 | 3.42E-02 | TRUE |
| 0.95 | 10 | random subsample high activity - symptomatic men | 3.37 | 1.48E-03 | TRUE |
| 0.95 | 10 | detected only - symptomatic only | 0.34 | 7.36E-01 | FALSE |
| 0.95 | 10 | high activity symptomatic men - symptomatic only | 3.91 | 3.06e-4 | TRUE |
| 0.95 | 10 | random subsample high activity - symptomatic only | 4.97 | 7e-6 | TRUE |
| 0.95 | 10 | symptomatic men - symptomatic only | 1.60 | 1.58E-01 | FALSE |
| 0.95 | 25 | detected only - high activity symptomatic men | -3.44 | 1.18E-03 | TRUE |
| 0.95 | 25 | detected only - random subsample high activity | -4.71 | 1.2e-5 | TRUE |
| 0.95 | 25 | high activity symptomatic men - random subsample high activity | -1.27 | 2.32E-01 | FALSE |
| 0.95 | 25 | detected only - symptomatic men | -1.26 | 2.32E-01 | FALSE |
| 0.95 | 25 | high activity symptomatic men - symptomatic men | 2.18 | 4.90E-02 | TRUE |
| 0.95 | 25 | random subsample high activity - symptomatic men | 3.45 | 1.18E-03 | TRUE |
| 0.95 | 25 | detected only - symptomatic only | 0.43 | 6.68E-01 | FALSE |
| 0.95 | 25 | high activity symptomatic men - symptomatic only | 3.87 | 3.7e-4 | TRUE |
| 0.95 | 25 | random subsample high activity - symptomatic only | 5.14 | 3e-6 | TRUE |
| 0.95 | 25 | symptomatic men - symptomatic only | 1.69 | 1.31E-01 | FALSE |
| 0.95 | 50 | detected only - high activity symptomatic men | -3.45 | 1.25E-03 | TRUE |
| 0.95 | 50 | detected only - random subsample high activity | -4.53 | 3e-5 | TRUE |
| 0.95 | 50 | high activity symptomatic men - random subsample high activity | -1.07 | 3.14E-01 | FALSE |
| 0.95 | 50 | detected only - symptomatic men | -1.10 | 3.14E-01 | FALSE |
| 0.95 | 50 | high activity symptomatic men - symptomatic men | 2.35 | 3.15E-02 | TRUE |
| 0.95 | 50 | random subsample high activity - symptomatic men | 3.42 | 1.25E-03 | TRUE |
| 0.95 | 50 | detected only - symptomatic only | 0.57 | 5.70E-01 | FALSE |
| 0.95 | 50 | high activity symptomatic men - symptomatic only | 4.02 | 1.95e-4 | TRUE |
| 0.95 | 50 | random subsample high activity - symptomatic only | 5.09 | 4e-6 | TRUE |
| 0.95 | 50 | symptomatic men - symptomatic only | 1.67 | 1.35E-01 | FALSE |

**Supplementary Table 7: Number of sequences and proportion of all infections captured**

| Dataset | Coverage Level | Median # Sequences | Range (# Sequences) | Median proportion of infections | Range (proportion of infections) |
| --- | --- | --- | --- | --- | --- |
| Detected Only | 100 | 27565 | 23506-32807 | 0.64 | 0.63-0.65 |
| Detected Only | 50 | 13779.5 | 11751-16400 | 0.32 | 0.31-0.32 |
| Detected Only | 25 | 6887 | 5873-8197 | 0.16 | 0.16-0.16 |
| Detected Only | 10 | 2752 | 2346-3276 | 0.064 | 0.063-0.065 |
| Detected Only | 5 | 1374 | 1171-1635 | 0.032 | 0.031-0.032 |
| Detected Only | 4 | 1097.5 | 936-1308 | 0.025 | 0.025-0.026 |
| Detected Only | 3 | 822 | 701-980 | 0.019 | 0.019-0.019 |
| Detected Only | 2 | 546.5 | 467-652 | 0.013 | 0.012-0.013 |
| Detected Only | 1 | 270.5 | 230-324 | 0.0063 | 0.0061-0.0064 |
| Symptomatic Only | 100 | 26237.5 | 22461-31183 | 0.61 | 0.6-0.61 |
| Symptomatic Only | 50 | 13115.5 | 11228-15590 | 0.3 | 0.3-0.31 |
| Symptomatic Only | 25 | 6555.5 | 5612-7792 | 0.15 | 0.15-0.15 |
| Symptomatic Only | 10 | 2618.5 | 2241-3113 | 0.061 | 0.06-0.061 |
| Symptomatic Only | 5 | 1307 | 1119-1554 | 0.03 | 0.03-0.03 |
| Symptomatic Only | 4 | 1044 | 893-1242 | 0.024 | 0.024-0.024 |
| Symptomatic Only | 3 | 782.5 | 669-931 | 0.018 | 0.018-0.018 |
| Symptomatic Only | 2 | 519 | 445-618 | 0.012 | 0.012-0.012 |
| Symptomatic Only | 1 | 257 | 219-307 | 0.006 | 0.0059-0.006 |
| Symptomatic Men | 100 | 17064.5 | 14181-20615 | 0.4 | 0.38-0.42 |
| Symptomatic Men | 50 | 8529.5 | 7087-10304 | 0.2 | 0.19-0.21 |
| Symptomatic Men | 25 | 4261.5 | 3541-5148 | 0.099 | 0.094-0.1 |
| Symptomatic Men | 10 | 1701.5 | 1414-2057 | 0.039 | 0.038-0.042 |
| Symptomatic Men | 5 | 848 | 705-1026 | 0.02 | 0.019-0.021 |
| Symptomatic Men | 4 | 677 | 562-819 | 0.016 | 0.015-0.017 |
| Symptomatic Men | 3 | 506 | 422-614 | 0.012 | 0.011-0.012 |
| Symptomatic Men | 2 | 336.5 | 278-407 | 0.0078 | 0.0074-0.0082 |
| Symptomatic Men | 1 | 166.5 | 137-202 | 0.0038 | 0.0037-0.0041 |
| High Activity Symptomatic Men | 100 | 7853 | 6647-9803 | 0.18 | 0.17-0.21 |
| High Activity Symptomatic Men | 50 | 3924 | 3320-4899 | 0.091 | 0.084-0.11 |
| High Activity Symptomatic Men | 25 | 1959.5 | 1657-2446 | 0.046 | 0.042-0.053 |
| High Activity Symptomatic Men | 10 | 780.5 | 660-976 | 0.018 | 0.017-0.021 |
| High Activity Symptomatic Men | 5 | 387.5 | 326-486 | 0.009 | 0.0082-0.01 |
| High Activity Symptomatic Men | 4 | 310 | 262-388 | 0.0072 | 0.0066-0.0084 |
| High Activity Symptomatic Men | 3 | 231 | 195-289 | 0.0054 | 0.0049-0.0062 |
| High Activity Symptomatic Men | 2 | 152 | 127-190 | 0.0035 | 0.0032-0.0041 |
| High Activity Symptomatic Men | 1 | 73 | 61-93 | 0.0017 | 0.0016-0.002 |
| Random Subsample High Activity | 100 | 2201 | 1673-2902 | 0.051 | 0.044-0.062 |
| Random Subsample High Activity | 50 | 1098.5 | 834-1448 | 0.025 | 0.022-0.031 |
| Random Subsample High Activity | 25 | 546.5 | 414-721 | 0.013 | 0.011-0.015 |
| Random Subsample High Activity | 10 | 215.5 | 163-284 | 0.005 | 0.0042-0.0061 |
| Random Subsample High Activity | 5 | 105 | 78-140 | 0.0024 | 0.0021-0.003 |
| Random Subsample High Activity | 4 | 82.5 | 61-111 | 0.0019 | 0.0016-0.0024 |
| Random Subsample High Activity | 3 | 61.5 | 47-83 | 0.0014 | 0.0012-0.0018 |
| Random Subsample High Activity | 2 | 39 | 28-53 | 0.0009 | 0.00073-0.0011 |
| Random Subsample High Activity | 1 | 19 | 10-24 | 0.00043 | 0.00027-0.00051 |

**Supplementary Table 8: Pair Formation Network Parameters**

| Parameter | Description | Group | | | Baseline Value | Rationale |
| --- | --- | --- | --- | --- | --- | --- |
| Total population | Number of people in the population |  | | | 10000 |  |
| Behavior proportions | Proportion of population in each behavior group | WSM | | | 0.475 | In Natsal-3 (UK, 2010-2012), 7.3% of men reported any sexual experience or contact with a same-sex partner, and 2.9% had at least one sexual partner of the same sex in the past 5 years.^1^ The NSFG (U.S. 2017-2019) found that 7.3% of men had ever had sex with another man and 3.2% had done so in the past 12 months.^2^ |
|  |  | MSW | | | 0.475 |  |
|  |  | MSM | | | 0.03 |  |
|  |  | MSMW | | | 0.02 |  |
| Proportion high activity | The proportion of each behavior group that is classified as high activity | WSM | | | 0.03 | NSFG 2015-2019 reported 2.0% of women and 2.9% of men had >= 5 opposite sex partners in the past year, 0.6% of women and 1.6% of men had sex in exchange for money or drugs.^2^  Nastal-3 found that 23.2% of MSM had >=5 male sexual partners in the past year, and 13.4% had unprotected anal intercourse with 2+ partners in the past year.^3^  The NSFG 2006-2010 found that in women 40-44, 2.3% had 40+ lifetime sexual partners.^4^ |
|  |  | MSW | | | 0.03 |  |
|  |  | MSM | | | 0.15 |  |
|  |  | MSMW | | | 0.15 |  |
| Allowed partner types | Who forms partners with whom | WSM | | | MSW, MSMW | Definition |
|  |  | MSW | | | WSM |  |
|  |  | MSM | | | MSM, MSMW |  |
|  |  | MSMW | | | WSM, MSM, MSMW |  |
| New partner request frequency distribution  (Shifted gamma distribution with shape, scale, and min) | The rate of seeking a new partner each year | All behavior groups, low activity | | | Shape = 1.5  Scale = 0.13  Min = 0.05  Median = 0.20 new partners per year; new partner every ~5 years | NSFG 2015-2019 found that 12.9% of WSM and 28.3% MSW reported >=15 lifetime partners.^8^ Natsal-3 found that 19.9% of women had >= 10 lifetime male sexual partners, and 33.9% of men had >= 10 lifetime female sexual partners.^1^  The NSFG 2006-2010 found that sexually active MSM had a mean of 2.3 partners in the last year.^9^  Natsal-3 found that 19.7% of men had at least one new female sexual partner in the past year, and 14.5% of women had at least one new male partner in the past year.^1^  One study looking at data from the Seatle sex Survey, the Urban Men’s Health Study, and the Seattle MSM RDD found a median of 1 sexual partner in the last year for MSW and WSM, and between 2-4 for MSM. It also found median lifetime numbers of sexual partners of 8 for MSW, 6 for WSM, and 45 for MSM.^10^ |
|  |  | All behavior groups, high activity | | | Shape = 5  Scale = 3  Min = 0.1  Median = 14.11 new partners per year |  |
| Maximum # of concurrent steady partners | The proportion of the population with a maximum steady partnership capacity of each value | High activity | | 0 | 0.5 | One analysis of the NHBS 2014 in the U.S. found that ~15% of MSM had 1 main partner only, and ~40% had main and casual partners in the past year.^11^ |
|  |  |  |  | 1 | 0.3 |  |
|  |  |  |  | 2 | 0.2 |  |
|  |  | Low Activity | 0 | | 0.0 |  |
|  |  |  | 1 | | 0.8 |  |
|  |  |  | 2 | | 0.2 |  |
| ρ | The probability that, when seeking a new partner, an MSMW individual will seek a female partner |  | | | 0.5 | Relevant to substantive question – tested values of 0.05, 0.25, 0.5, 0.75, and 0.95 |
| Steady partnership duration distribution (Gamma shape, scale, and median) | Each new steady partnership has a duration drawn from this distribution | All | | | Shape = 4  Scale = 638.5  Median = 2345.53 or about 6.4 years | Based on the Kretzschmar 1996 modeling study parameterization of the average duration for a steady partnership is 6.9 years^5^ |
| Casual partnership duration distribution (exponential rate parameter) | Each new casual partnership has a duration drawn from this distribution | All | | | Rate = 0.1 | Based on the Kretzschmar 1996 modeling study parameterization where the average duration of casual partnership was 10 days.^5^ |

**Supplementary Table 9: Gonorrhea Transmission Parameters**

| Parameter | Description | Group | | | Baseline Value | Sources | Notes / Rationale |
| --- | --- | --- | --- | --- | --- | --- | --- |
| Initial gonorrhea prevalence | Proportion of each behavior stratum infected with gonorrhea at the start of the simulation | WSM | | All | 0.015 | The U.S. CDC’s STD Surveillance Network found that test positivity for rectal gonorrhea was 17.1% in MSM in 2018.^12^ The NHBS in 2017 found rectal and pharyngeal gonorrhea prevalences in community venue attending MSM to be 4.5% and 4.6% respectively.^13^ A U.S. study in 2019 found rectal gonorrhea prevalences of 2.3% in cisgender MSM, 4.3% in TGW, and 1.4% in MSW. ^14^ Pollock et al. estimated gonorrhea prevalences between 0.1-0.4% for men and women.^15^ The EMERALD study of female sex workers in Baltimore 2017-2019 found a gonorrhea prevalence of 18%.^16^  The overall prevalence of gonorrhea in MSM in modeling studies Reichert et al and Kline et al. was 3%.^17,18^ Oliveira Roster et al’s modeling study assumed a high activity MSM prevalence of 8%.^19^ | |
|  |  |  |  | High | 0.08 |  |  |
|  |  |  |  | Low | (from model) |  |  |
|  |  | MSW | | All | 0.015 |  |  |
|  |  |  |  | High | 0.08 |  |  |
|  |  |  |  | Low | (from model) |  |  |
|  |  | MSM | | All | 0.045 |  |  |
|  |  |  |  | High | 0.11 |  |  |
|  |  |  |  | Low | (from model) |  |  |
|  |  | MSMW | | All | 0.04 |  |  |
|  |  |  |  | High | 0.10 |  |  |
|  |  |  |  | Low | (from model) |  |  |
| Initial proportion symptomatic | Proportion of initially seeded infections that are symptomatic | Men (MSM, MSMW, MSW) | | | 0.089 | From Reichert et al^17^ | |
|  |  | Women (WSM) | | | 0.05 |  |  |
| Symptomatic infection duration distribution  (discretized gamma shape and scale, median) | How long it takes for a symptomatic infected individual to seek care, get treatment, and recover (time from infection to recovery) | Men (MSM, MSMW, MSW) | | | Shape: 4  Scale: 2.5  Median: 8.26 days | Based on other modeling studies^5,17–19^ | |
|  |  | Women (WSM) | | | Shape: 5  Scale: 2.4  Median: 11.21 days |  |  |
| Asymptomatic infection duration distribution  (discretized gamma shape and scale, median) | Average time to next asymptomatic gonorrhea screening | MSM, MSMW – high activity | | | Shape: 3  Scale: 24.3  Median: 64.98 days | The U.S. CDC recommends annual gonorrhea screening for sexually active women <25 years old or high-risk women >= 25. MSM should be screened at least annually, and high risk individuals should be screened every 3-6 months.^20^ CDC data from SSuN has shown that about 60% of MSM get rectal gonorrhea screening at least once a year.^12^  Other modeling studies have adapted these numbers similarly.^18,19^ | |
|  |  | WSM/MSW – high activity | | | Shape: 2.5  Scale: 110  Median: 239.33 days |  |  |
|  |  | All groups – low activity | | | Shape: 5  Scale: 73  Median: 340.98 days |  |  |
| Natural clearance duration distribution (discretized gamma shape and scale, median) | How long it takes for someone to naturally clear an infection without being tested or treated | Men (MSM, MSMW, MSW) | | | Shape: 9  Scale: 7  Median: 60.68 days | Barbee et al. 2021 and 2022 estimated the natural duration of pharyngeal and rectal gonorrhea infections to be a median of 16.3 weeks and 9 weeks respectively.  Other modeling studies have adapted these numbers similarly. ^18,19,21,22^ | |
|  |  | Women (WSM) | | | Shape: 10  Scale: 8  Median: 77.35 days |  |  |
| Transmission probability | Probability of transmission per partnership per day | MSW, MSMW -> WSM | Steady | | 0.15 | As used in Kretzschmar 1996 et al.^5^ | |
|  |  | WSM -> MSW, MSMW | Steady | | 0.0625 |  |  |
|  |  | MSW, MSMW -> WSM | Casual | | 0.6 |  |  |
|  |  | WSM -> MSW, MSMW | Casual | | 0.25 |  |  |
|  |  | MSM,MSMW -> MSM, MSMW | Steady | | 0.25 | Assumption | |
|  |  | MSM, MSMW -> MSM, MSMW | Casual | | 0.75 |  |  |
| Probability of symptoms | Probability that a new infection is symptomatic | Men (MSW, MSMW, MSM) | | | 0.65 | From Oliveira Roster et al.^19^ | |
|  |  | Women (WSM) | | | 0.54 |  |  |

**Supplementary Table 10: Sequence simulation parameters**

| Parameter | Description | Value | Source | Notes |
| --- | --- | --- | --- | --- |
| Sequence length | How many nucleotides to simulate | 10000 | -- | Should be long enough for no reversions and short enough to be manageable |
| Scale | Scales branch lengths so that time is in terms of substitutions rather than days | 2.254247e-06 | ^7^ | 3.74e-6 subs per site per year * 2.2e6 = 8.2 subs per year /365 = 0.023 subs per genome per day / 10000 = 2.254247e-06 subs per site per day |
| Evolutionary model | GTR | Rate parameters: A-C: 1.00578 A-G: 4.59475 A-T: 0.74535 C-G: 0.46171 C-T: 4.55284 G-T: 1.00000 | Assumed to be reasonable estimates | GTR = specify different rates for each transition / transversion |
| Base frequencies |  | Base frequencies: A: 0.154 C: 0.346 G: 0.343 T: 0.157 | Assumed to be reasonable estimates |  |
| Gamma rate heterogeneity | Shape (alpha) parameter | 0.450 | Assumed to be reasonable estimates |  |

**Supplementary Table 11: Simulation run parameters**

| Parameter | Baseline Value |
| --- | --- |
| Partnership burn-in duration (days) | 2000 |
| Transmission burn-in duration (days) | 10000 |
| Simulation period (days) | 3650 |

### **Supplementary References**

1.	Mercer CH, Tanton C, Prah P, et al. Changes in sexual attitudes and lifestyles in Britain through the life course and over time: findings from the National Surveys of Sexual Attitudes and Lifestyles (Natsal). *The Lancet*. 2013;382(9907):1781-1794. doi:10.1016/S0140-6736(13)62035-8

2.	NSFG - Listing S - Key Statistics from the National Survey of Family Growth. August 19, 2025. Accessed November 13, 2025. https://web.archive.org/web/20250819093310/https://www.cdc.gov/nchs/nsfg/key_statistics/s-keystat.htm#sexactbetwmen

3.	Prah P, Hickson F, Bonell C, et al. Men who have sex with men in Great Britain: comparing methods and estimates from probability and convenience sample surveys. *Sex Transm Infect*. 2016;92(6):455-463. doi:10.1136/sextrans-2015-052389

4.	Haderxhanaj LT, Leichliter JS, Aral SO, Chesson HW. Sex in a Lifetime: Sexual Behaviors in the United States by Lifetime Number of Sex Partners, 2006–2010. *Sex Transm Dis*. 2014;41(6):345-352. doi:10.1097/OLQ.0000000000000132

5.	Kretzschmar M, Van Duynhoven YTHP, Severijnen AJ. Modeling Prevention Strategies for Gonorrhea and Chlamydia Using Stochastic Network Simulations. *American Journal of Epidemiology*. 1996;144(3):306-317. doi:10.1093/oxfordjournals.aje.a008926

6.	Rambaut A, Grass NC. Seq-Gen: an application for the Monte Carlo simulation of DNA sequence evolution along phylogenetic trees. *Bioinformatics*. 1997;13(3):235-238. doi:10.1093/bioinformatics/13.3.235

7.	Sánchez-Busó L, Golparian D, Corander J, et al. The impact of antimicrobials on gonococcal evolution. *Nat Microbiol*. 2019;4(11):1941-1950. doi:10.1038/s41564-019-0501-y

8.	NSFG - Listing N - Key Statistics from the National Survey of Family Growth. August 17, 2025. Accessed November 13, 2025. https://web.archive.org/web/20250817021129/https://www.cdc.gov/nchs/nsfg/key_statistics/n-keystat.htm#numberlifetime

9.	Leichliter JS, Haderxhanaj LT, Chesson HW, Aral SO. Temporal Trends in Sexual Behavior Among Men Who Have Sex With Men in the United States, 2002 to 2006–2010. *JAIDS Journal of Acquired Immune Deficiency Syndromes*. 2013;63(2):254. doi:10.1097/QAI.0b013e31828e0cfc

10.	Glick SN, Morris M, Foxman B, et al. A comparison of sexual behavior patterns among men who have sex with men and heterosexual men and women. *J Acquir Immune Defic Syndr*. 2012;60(1):83-90. doi:10.1097/QAI.0b013e318247925e

11.	Chapin-Bardales J, Rosenberg ES, Sullivan PS, Jenness SM, Paz-Bailey G. Trends in Number and Composition of Sex Partners Among Men Who Have Sex With Men in the United States, National HIV Behavioral Surveillance, 2008–2014. *J Acquir Immune Defic Syndr*. 2019;81(3):257-265. doi:10.1097/QAI.0000000000002025

12.	Llata E, Cuffe K, Picchetti V, Braxton J, Torrone E. Demographic, Behavioral, and Clinical Characteristics of Persons Seeking Care at Sexually Transmitted Disease Clinics — 14 Sites, STD Surveillance Network, United States, 2010–2018 | MMWR. November 5, 2021. Accessed September 29, 2025. https://www.cdc.gov/mmwr/volumes/70/ss/ss7007a1.htm

13.	Johnson Jones ML, Chapin-Bardales J, Bizune D, et al. Extragenital Chlamydia and Gonorrhea Among Community Venue-Attending Men Who Have Sex with Men - Five Cities, United States, 2017. *MMWR Morb Mortal Wkly Rep*. 2019;68(14):321-325. doi:10.15585/mmwr.mm6814a1

14.	Mimiaga MJ, Tian J, Chiu I, et al. Prevalence of chlamydial, gonococcal and syphilis positivity by anatomical site and sex via self-collected biospecimens using at-home testing kits among a large and diverse cohort of men and women enrolled in the MACS/WIHS Combined Cohort Study (MWCCS) in the USA. *Sex Transm Infect*. Published online July 25, 2025. doi:10.1136/sextrans-2025-056508

15.	Pollock ED, Clay PA, Kreisel KM, Spicknall IH. Estimated Incidence and Prevalence of Gonorrhea in the United States, 2006–2019. *Sexually Transmitted Diseases*. 2023;50(4):188. doi:10.1097/OLQ.0000000000001763

16.	Sherman SG, Tomko C, White RH, et al. Structural and Environmental Influences Increase the Risk of Sexually Transmitted Infection in a Sample of Female Sex Workers. *Sex Transm Dis*. 2021;48(9):648-653. doi:10.1097/OLQ.0000000000001400

17.	Reichert E, Yaesoubi R, Rönn MM, Gift TL, Salomon JA, Grad YH. Resistance-minimising strategies for introducing a novel antibiotic for gonorrhoea treatment: a mathematical modelling study. *The Lancet Microbe*. 2023;4(10):e781-e789. doi:10.1016/S2666-5247(23)00145-3

18.	Kline MC, Roster KO, Helekal D, Rumpler E, Grad YH. Comparing Strategies to Introduce Two New Antibiotics for Gonorrhea: A Modeling Study. *medRxiv*. Preprint posted online July 3, 2025:2025.07.01.25330638. doi:10.1101/2025.07.01.25330638

19.	Oliveira Roster KI, Rönn MM, Elder H, et al. Estimating the undetected burden and the likelihood of strain persistence of drug-resistant Neisseria gonorrhoeae. *American Journal of Epidemiology*. Published online December 13, 2024:kwae455. doi:10.1093/aje/kwae455

20.	Gonococcal Infections Among Adolescents and Adults - STI Treatment Guidelines. September 2, 2025. Accessed November 13, 2025. https://web.archive.org/web/20250902050308/https://www.cdc.gov/std/treatment-guidelines/gonorrhea-adults.htm

21.	Barbee LA, Soge OO, Khosropour CM, et al. The Duration of Pharyngeal Gonorrhea: A Natural History Study. *Clin Infect Dis*. 2021;73(4):575-582. doi:10.1093/cid/ciab071

22.	Barbee LA, Khosropour CM, Soge OO, et al. The Natural History of Rectal Gonococcal and Chlamydial Infections: The ExGen Study. *Clinical Infectious Diseases*. 2022;74(9):1549-1556. doi:10.1093/cid/ciab680
